# Post-malnutrition growth and its associations with child survival and non-communicable disease risk: A secondary analysis of the Malawi ‘ChroSAM’ cohort

**DOI:** 10.1101/2022.02.25.22271497

**Authors:** Natasha Lelijveld, Sioned Cox, Kenneth Anujuo, Abena S Amoah, Charles Opondo, Tim J. Cole, Jonathan C. Wells, Debbie Thompson, Kimberley McKenzie, Mubarek Abera, Melkamu Berhane, Marko Kerac, CHANGE study collaborators group

## Abstract

**Background:** Rapid catch-up growth after prenatal undernutrition is associated with increased risk of non-communicable diseases (NCDs) in high-income countries. Severe malnutrition treatment programmes in low- and middle-income countries promote rapid post-malnutrition growth (PMGr) as desirable. Our aim was to explore patterns of PMGr during and in the year following treatment, and describe associations with survival and NCD risk seven years post-treatment.

**Methods:** Secondary data analysis from a cohort of children treated for severe malnutrition in Malawi in 2006/7. Six definitions of PMGr were derived based on a variety of timepoints, weight, weight-for-age z-score (WAZ) and height-for-age z-score (HAZ). Three categorisation methods included: no categorisation, quintiles, and latent class analysis (LCA). Associations with mortality risk, and with eight NCD indicators were analysed visually using scatter plots and boxplots, and statistically using simple and multivariable linear regression.

**Findings:** Faster weight gain was associated with lower risk of death (g/day during treatment aOR 0.99, 95%CI 0.99 to 1.00, p=0.04; after treatment g/kg/month aOR 0.91, 95% CI 0.87 to 0.94, p<0.001). In survivors, it was associated with greater hand grip strength in some instances (g/day during treatment 0.02, 95%CI 0.00 to 0.03, p=0.007) and larger HAZ 7-years post-discharge (adjusted Δ WAZ per day during treatment 6.62, 95%CI 1.31 to 11.9, p=0.02), both indicators of better health. However, faster weight gain in treatment was also associated with increased waist:hip ratio (adjusted g/day during treatment 0.02, 95%CI 0.01 to 0.03, p=0.003), a key indicator of later life NCD risk. The clearest patterns of association were seen when defining PMGr based on weight gain in g/day during treatment, and using the LCA method to describe growth patterns. Weight deficit at admission was a major confounder.

**Conclusion:** We found a complex pattern of benefits and risks associated with faster PMGr with a possible trade-off between short- and long-term benefits/risks. Peripheral versus visceral weight distribution in particular requires further exploration. Both initial weight deficit and rate of weight gain have important implications for future health. Because conclusions from observational studies can go only so far, future randomised intervention trials are needed.

## Introduction

Undernutrition in childhood and adult non-communicable diseases (NCDs) are important, global public health problems. In all its forms, malnutrition accounts for some 45% of all mortality in children under five years (1). Severe malnutrition, particularly wasting, threatens the survival of an estimated 47 million children under five in low and middle-income countries (LMICs) (2). In parallel, obesity-related NCDs are emerging as a leading cause of death among adults in these settings, with nearly three quarters of all NCD deaths occurring in LMICs (3). The need to tackle this ‘double burden’ of malnutrition is increasingly urgent(4).

There is convincing evidence that the two major types of malnutrition, undernutrition and overnutrition, are linked via the DoHAD (Developmental Origins of Health and Disease) hypothesis. This posits that prenatal undernutrition results in increased NCD risk later in life, and also that severe malnutrition during postnatal periods of developmental plasticity is linked to increased risk of NCDs (5-8). Mechanisms include the “capacity-load” model whereby the metabolic capacity for homeostasis is developed in the first 1000 days of life and any disturbance or limitation during this period results in reduced capacity to deal with metabolic stressors later in life(9). This reduced homeostatic capacity manifests as increased later-life NCD risk.

Evidence from high-income countries (HICs) on low birth weight (LBW), a marker of prenatal undernutrition, links the rate of catch-up growth in undernourished infants with childhood risk-markers for NCDs (10-13). For example, infants born small for gestational age who were given a nutrient-enriched formula to promote faster weight gain had higher blood pressure 6-8 years later than those given control formula that resulted in slower weight gain (11). In HIC contexts, where the short-term risk of death from other morbidities is low and environmental stressors such as infectious disease load are relatively few, promoting slower post-malnutrition growth (PMGr) can thus be seen as preferable.

In contrast, in LMIC contexts with high risks of short-term mortality, the balance of short- and long-term risks associated with PMGr is unknown. Given the strong associations between severe undernutrition (as assessed by low weight and other anthropometric deficits) and high child mortality (14), many treatment programmes prioritise fast recovery to ‘normal’ weight. Children suffering from severe wasting and/or nutritional oedema are currently treated with high-calorie, high-fat ready-to-use therapeutic food (RUTF) or therapeutic milks (f100) that promote rapid weight gain (15). The rationale is that this will reduce the immediate mortality risks associated with being small. However, this priority has been questioned in wider public health literature (16-18). The long-terms risk associated with rapid PMGr for survivors of severe malnutrition is currently unknown. More data is needed to find the right balance between a rate of post-malnutrition weight and height gain which optimises survival whilst minimizing long-term NCD risk.

To address this research gap, we must consider what constitutes adequate vs too rapid vs too slow PMGr and how to define this. There is currently no standard definition. Data are also needed on what effect PMGr may have on later life NCD risks. Our objectives in this analysis were therefore: (1) to explore rates of PMGr based on a variety of weight- and height-related definitions; (2) to develop different ways of categorising these PMGr definitions into faster and slower growth; and (3) to examine associations and patterns between the different definitions and categorizations of PMGr with NCD risk markers.

## Methods

This was a secondary analysis of data collected from children treated for severe wasting and/or nutritional oedema in 2006/7 in Malawi, and followed up seven years later to assess risk of NCD.

The data come from a prospective cohort of severely malnourished children recruited into a randomised controlled trial of pre- and probiotics (PRONUT study) (19). The intervention in this trial showed no overall effect on nutritional recovery nor weight gain. The same children were subsequently followed up at 1-year post-discharge (FuSAM study) (20) and again at 7-years post-discharge (ChroSAM study), where the focus was on longer term growth and on indicators of NCD risk (7).

All children originally admitted for treatment during the recruitment period were eligible for inclusion into this current analysis. Hence our sample size was constrained by this limit: a-priori formal sample size calculations for our analyses was not done. Admission criteria at the time of the original PRONUT study were weight-for-length z-score <70% median (using the NCHS reference) and/or MUAC<110 mm and/or bilateral oedema. All patients received initial inpatient care for there was no stand-alone outpatient-based CMAM (Community Management of Acute Malnutrition) programme in the area at that time. Children were initially stabilized with F75 therapeutic milk. As clinical status and appetite improved, they moved to a ‘transition phase’ diet where RUTF (Ready to Use Therapeutic Food) was introduced alongside F75 milk. After continued improvement, they moved to a ‘rehabilitation phase’ in which they were transferred home with a 2-week ration of RUTF to continue their recovery as outpatients, returning to the ward for monitoring and to collect another ration every 2 weeks until recovery. Children were discharged from the programme after 2 consecutive visits at or above the target weight of 80% NCHS median weight-for-height. Minimum time in outpatient care was therefore 4 weeks, with a maximum time of 10 weeks before classification as a ‘non-responder’. Anthropometric data were thus available at: admission to treatment; in-treatment minimum weight; at transfer from hospital to home-based care; at programme discharge; at 1- and 7-years post-discharge.

### Definitions and classifications of PMGr (exposure variables)

At present, there is no one standard definition of PMGr and we thus explored a range of possible definitions. Six were created, based on the available data and assessment methods commonly used by programmers and policy makers working in this area (**Table 1**).

**Table 1:**
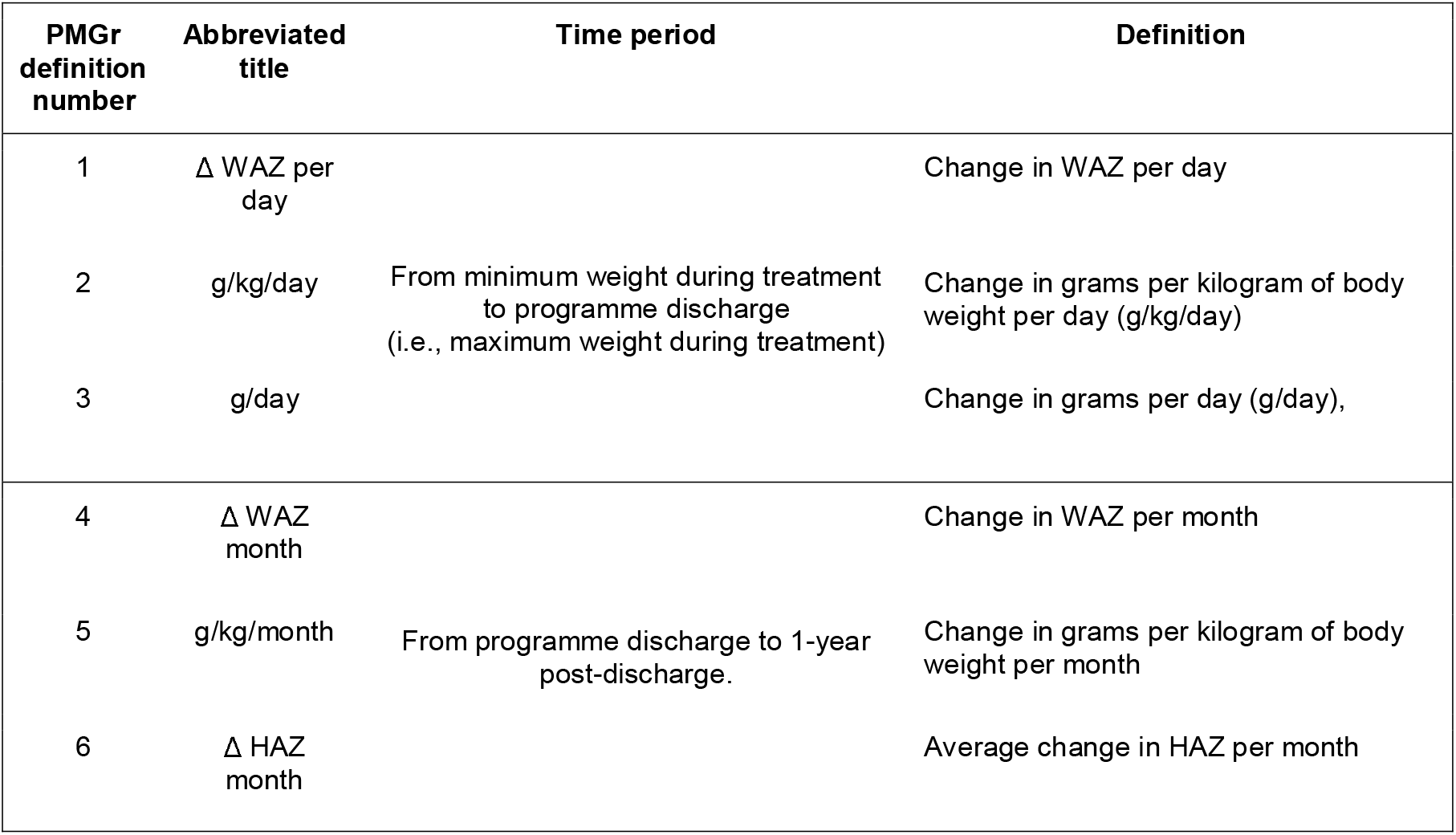
Definitions of PMGr.

Three definitions focus on the in-treatment period, from the point when children were at their minimum weight during treatment, which was often at admission unless oedema first needed to subside, until discharge from care, which was also their point of maximum weight during treatment. These definitions are based on weight for age z-score (WAZ) (using WHO 2006 growth standards), grams per kilogram (g per kg) of body weight and absolute weight in grams. Height-for-age z-score (HAZ) or other measures of height change was not included since there was minimal if any gain in height during the few weeks of in-programme treatment. Since the length of stay until discharge criteria were met varied by individual, gains in weight indicators were calculated ‘per day’ to capture rates of gains.

Three other PMGr definitions were based on the post-treatment period, between discharge and 1-year post-discharge. We expected a more “natural” level of catch-up growth during this time since high-energy additional foods were no longer being provided. Again, this included WAZ and g per kg, and an additional metric: HAZ (WHO 2006). Growth increment was calculated per month to account for any differences in follow up period.

Each definition of PMGr was subsequently categorised using each of three different methods:

1. No categorisation (continuous data)
2. Quintiles (Q1 slowest and Q5 fastest)
3. Latent class analysis (LCA) machine learning (6 classes)

### NCD risk indicators (outcome variables)

Seven NCD indicators were selected as outcomes of interest, and assessed 7 years post-discharge from treatment. These were: blood pressure (systolic and diastolic), handgrip strength, waist circumference, waist/hip ratio, lean mass index (LMI), fat mass index (FMI) and HAZ. These indicators are all commonly associated with NCD risk in adulthood and were previously analysed as part of the ChroSAM study (21-25). Blood pressure was assessed using an Omron digital device with a paediatric cuff. Muscle strength was measured with a Takei Grip-D device (Takei, Niigata, Japan). LMI and FMI were assessed by bioelectrical impedance analysis (BIA) using a Quadscan 4000 device (Bodystat, Douglas, Isle of Man). Usually, the BIA output impedance (Z) is adjusted for height (HT) to give the impedance index (HT^2^/Z), which is then used to predict to total body water or lean mass via population-specific empirical equations. Lean mass can then be divided by height-squared, to give LMI in the same kg/m^2^ units as BMI. Obtained via impedance, however, LMI is essentially HT^2^/Z/HT^2^, which simplifies to 1/Z. In the absence of a population-specific equation, 1/Z can therefore be used as a valid marker of LMI, as it differs only by two constants (26). A proxy for FMI can then be calculated as the regression residual of BMI on LMI, on the assumption that greater BMI for a given LMI value indicates increased adiposity (26). Lastly, while HAZ change during early post-treatment period was an exposure variable in these analyses, its assessment at 7-years post-discharge was also and outcome indicator of NCD risk (27).

### Statistical Analyses

Data were analysed using Stata SE 17 (StataCorp LLC, College Station, Texas, USA). Implausible z-scores exceeding +/- 10 were set as missing. For the ‘no-categorisation’ method, each PMGr definition was assessed for its association with death using data from the full prospective cohort, using logistic regression. Associations between PMGr definitions and NCD outcomes were analysed using only data from survivors, using linear regression. Age, sex, and HIV status were included as *a priori* confounding factors in adjusted regression analyses, based on the literature and previous analyses (7). HAZ at follow-up was also included as a potential confounder when examining blood pressure as the outcome, as a known confounder (associated with both rate of growth and blood pressure) in the literature. Adjusting for end-point BMI was explored but not included since it was considered to be on the causal pathway between rapid weight gain and NCD risk factors.

For the quintiles method, individuals were assigned to quintiles based on each of the PMGr definitions; quintile 1 being the smallest values and quintile 5 the highest. Associations between PMGr definitions and NCD outcomes were explored visually using boxplots, and using linear regression where the NCD risk factor was a continuous outcome and the quintile number (1-5) was the predictor.

For the LCA method, latent classes were identified using a generalised structural equation model with Gaussian-distributed WAZ, weight, and HAZ measurements across five timepoints (admission, minimum weight during treatment, discharge from inpatient care, discharge from care, and one year post-discharge). Since LCA uses anthropometric values at multiple fixed timepoints to identify patterns, it does not account for different lengths of treatment. A balance of sample size and model fit according to Bayesian information criteria (BIC) were used to determine the number of classes to extract; models with lower BIC and classes with at least eight individuals each were preferred. We also conducted additional LCAs with WAZ, weight and HAZ at each timepoint standardised by admission WAZ/weight/HAZ respectively, in order to adjust for the confounding influence of admission anthropometry.

The LCA-predicted PMGr classes were plotted against the relevant anthropometric measures to facilitate comparison. As with the quintiles-based method, boxplots for each of the LCA-derived classes and NCD indictors were created to explore associations, and simple and multivariable linear regression analyses were conducted.

### Ethical approval

Ethical approval for the 7-year follow up study was granted by the Malawi College of Medicine Research and Ethics Committee (reference P.02/13/1342), and the University College London Research Ethics Committee (reference 4683/001). Approval for this secondary analysis was granted by London School of Hygiene and Tropical Medicine (LSHTM) MSc Research Ethics Committee (reference 26076).

## Results

Anthropometric data at the point of discharge from treatment was available for 786 children. Follow-up data was available for 477 children up until 1-year post discharge. Data on NCD risk factors at 7-years post-discharge was available for 320 children. These data, which were used for defining PMGr, are described in Figure 1.

**Figure 1:**
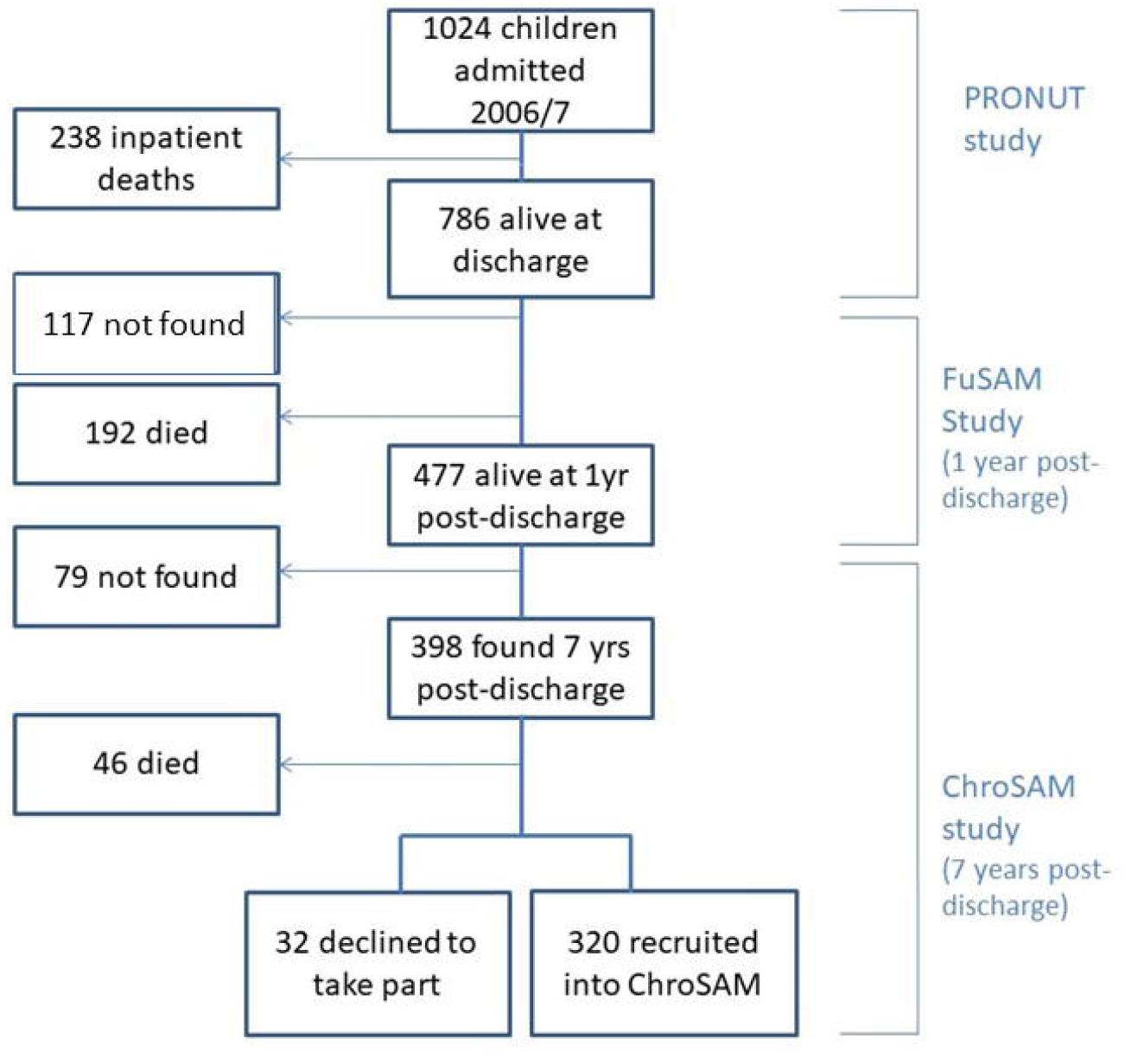
Participant flow diagram.

### Survival

Using the ‘no categorisation’ method, faster growth during treatment (change in WAZ per day, g/kg/day, and g/day) was associated with significantly reduced odds of death (crude odds ratio 0.001, 0.98, and 0.99 respectively) (**Table 2**). However, after adjustment for age and HIV, there was no association between treatment growth rate and odds of death for change in WAZ per day (aOR 0.19, 95%CI 0.01 to 6.34, p=0.35) and g/kg/pay (aOR 0.99, 95%CI 0.97 to 1.01, p=0.55). Younger age and HIV were positively associated with increased odds of death, and HIV was also associated with slower weight gain during treatment. Between treatment and 1-year post-discharge, only faster growth in g/kg/month was associated with reduced odds of death, both before and after adjustment (adjusted odds ratio 0.91, 95%CI 0.87 to 0.94, p<0.001).

**Table 2:**
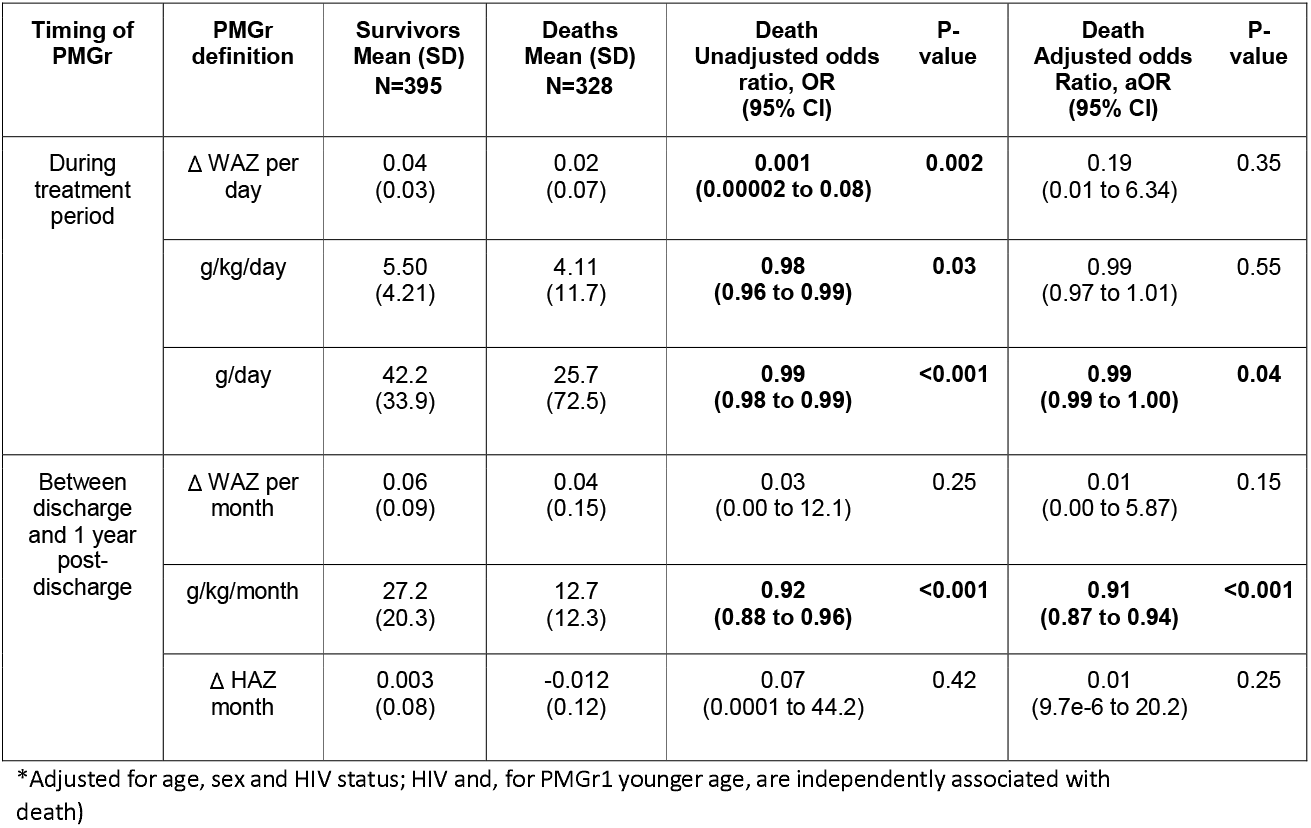
Logistic Regression of mortality on growth rate (PMGr as continuous predictor) (n=320)

### Descriptive analyses of survivors

A total of 320 survivors seen at 7 years post-discharge were included in this analysis **(Figure 1)**. At admission, the sample had a high prevalence of HIV (approximately 30%) and 83% had oedematous malnutrition (**Table 3**). Three-quarters were stunted (HAZ<-2) at admission (78%) and a large proportion (41%) remained stunted 7 years post-discharge. Across all six definitions of PMGr, children had wide ranging changes in growth status, including both positive and negative (**Table 4, Annex Figure 1**). The method of defining PMGr influenced the pattern of WAZ change over the course of the cohort timepoints (**Annex Figure 2**). Both the quintile categories and the LCA-defined categories (described in **Figure 2**) highlight the link between greater anthropometric deficits at admission to treatment and rapid PMGr; this is especially true for post-discharge growth **(Table 4)**. Those with larger WAZ at admission have slower PMGr after treatment (Table 4). The sample size for each of the LCA classes is presented in **Annex Table 1**.

**Table 3:** Descriptive demographics of survivors at different timepoints (n=320)

| Variables | Admission | Discharge | 1 year post-discharge | 7 year post-discharge |
| --- | --- | --- | --- | --- |
| Age (months)<br>Median IQR | 24.0 (16-34) | 25.7 (18 – 35) | 41.5 (33 – 50) | 111.4<br>(104 – 124) |
| % Males | 54.6% |  |  |  |
| % HIV positive | 29.5% |  |  |  |
| Anthropometry |  |  |  |  |
| % Oedema | 83.1% | 0.0% | 0.32% | 0.0% |
| Weight (kg) (mean, SD) | 8.68 (2.74) | 9.58 (2.76) | 13.04 (3.16) | 23.99 (4.91) |
| Height (cm) (mean, SD) | 78.23 (10.2) | 78.71 (9.88) | 88.38 (11.3) | 124.99 (9.03) |
| MUAC (cm) (mean, SD) | 12.33 (1.89) | 13.52 (1.76) | 15.22 (1.49) | 17.24 (1.96) |
| Anthropometric Indicators |  |  |  |  |
| % Stunted (HAZ<-2) | 78.2% | 81.7% | 81.9% | 41.1% |
| % Wasted by MUAC (<12.5cm) | 51.9% | 26.0% | 2.9% | 0% |
| % Wasted by WHZ (<-2) | 49.8% | 19.7% | 3.0% | 0% |
| % Underweight (WAZ<-2) | 75.4% | 62.1% | 40.0% | 33.0% |
| Days since admission<br>(median, IQR) | 0 | 35 (30-51) | 531<br>(493 – 558) | 2682<br>(2611-2755) |

Z-scores based on WHO2006 and WHO2007 references

**Table 4:** Summary statistics by each of the PMGr definition, and their association with admission WAZ (n=320 survivors)

| PMGr definition | Mean change (SD) | Minimum | Maximum | Change in growth rate per unit increase in admission WAZ* |
| --- | --- | --- | --- | --- |
| 1<br>Δ WAZ per day | 0.03 (0.03) | -0.04 | 0.21 | -0.001<br>(0.93, -0.002) |
| 2<br>g/kg/day | 5.31 (3.66) | -3.66 | 27.39 | **-0.36**<br>**(-0.63, -0.09)** |
| 3<br>g/day | 40.6 (27.1) | -38.6 | 155.9 | 0.83<br>(-1.05, 2.72) |
| 4<br>Δ WAZ per month | 0.06 (0.08) | -0.14 | 0.39 | **-0.03**<br>**(-0.03, -0.02)** |
| 5<br>g/kg/month | 26.3 (18.1) | -3.8 | 117.0 | **-5.17**<br>**(-6.48, -3.87)** |
| 6<br>Δ HAZ per month | -0.00 (0.08) | -0.24 | 0.29 | **-0.02**<br>**(-0.02, -0.01)** |

\*Results of linear regression analysis, presented as coefficient (95% CI). WAZ= weight for age z-score. Bold= p<0.05.

**Figure 2:**
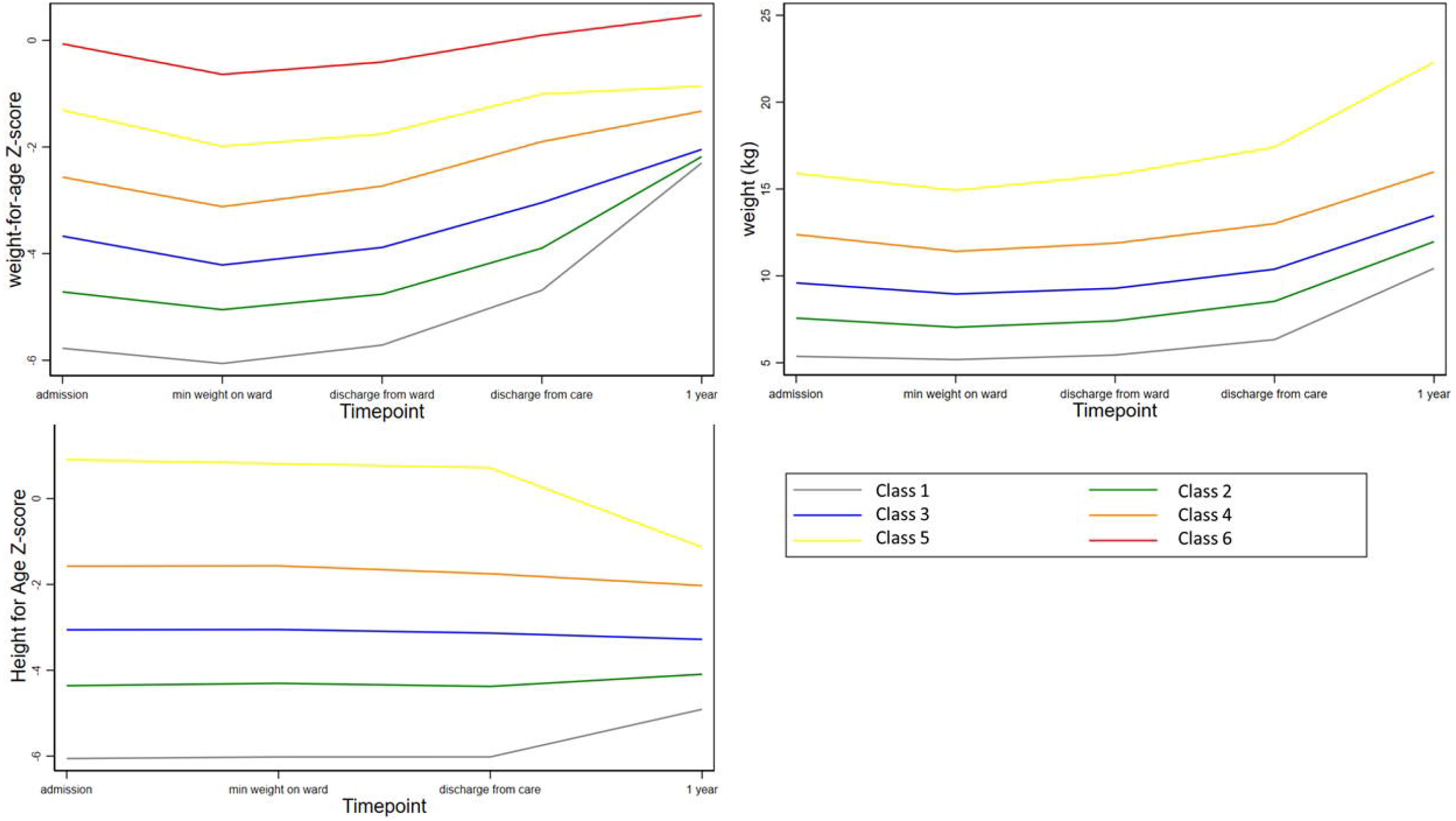
Growth patterns for WAZ, weight, and HAZ described based on LCA classes.

### NCD outcomes

#### No categorisation method

There were mixed patterns of association between rapid PMGr and NCD outcomes in this young cohort; only a few indicators of PMGr were significantly associated with NCD indicators after adjusting for age, sex and HIV status (**Table 5**). In the unadjusted analysis, the general trend was that faster weight gain was associated with reduced blood pressure, increased waist:hip ratio, and increased HAZ, with mixed associations across the other NCD outcomes. In the adjusted analysis, faster WAZ and g/kg/day gains during treatment were associated with a larger waist circumference, and faster gains in g/day during treatment was associated with larger waist:hip ratio. Faster gains in WAZ and g/day during treatment were associated with significantly larger HAZ at 7-year follow-up. Post-discharge, faster gains in HAZ were associated with a smaller waist circumference. See full panel of scatter plots for all NCD outcomes in **Annex Figure 3a-h**.

Class 1 = lowest initial admission weight and the steepest change.

Classes 5&6 = least / shallowest change

**Table 5:** Association between PMGr definitions and NCD risk outcomes (linear regression)

| NCD indicator | PMGr definition | Unadjusted difference | P-value | 95% CI | Adjusted difference | P-value | 95% CI |
| --- | --- | --- | --- | --- | --- | --- | --- |
| Systolic BP (mm Hg) | 1 | 9.24 | 0.69 | (-33.2, 51.7) | -14.7 | 0.50 | (-57.5, 28.1) |
|  | 2 | 0.00 | 0.98 | (-0.29, 0.30) | -0.04 | 0.79 | (-0.32, 0.24) |
|  | 3 | 0.03 | 0.07 | (-0.00, 0.06) | -0.01 | 0.76 | (-0.04, 0.03) |
|  | 4 | **-19.3** | **0.008** | **(-33.5, -5.04)** | -12.3 | 0.12 | (-27.9, 3.31) |
|  | 5 | **-0.09** | **0.01** | **(-0.15, -0.02)** | -0.04 | 0.28 | (-0.11, 0.03) |
|  | 6 | -5.95 | 0.46 | (-22.0, 10.1) | -9.49 | 0.24 | (-25.4, 6.39) |
| Diastolic BP (mm Hg) | 1 | -30.93 | 0.12 | (-70.85, 9.00) | -33.3 | 0.12 | (-75.5, 8.87) |
|  | 2 | -0.19 | 0.17 | (-0.46, 0.08) | -0.18 | 0.20 | (-0.45, 0.09) |
|  | 3 | -0.01 | 0.51 | (-0.04, 0.02) | -0.17 | 0.29 | (-0.05, 0.01) |
|  | 4 | **-14.1** | **0.04** | **(-27.8, -0.38)** | -13.4 | 0.09 | (-29.2, 2.34) |
|  | 5 | -0.05 | 0.11 | (-0.11, 0.01) | -0.04 | 0.23 | (-0.11, 0.03) |
|  | 6 | 2.94 | 0.71 | (-12.5, 18.4) | -2.44 | 0.77 | (-18.5, 13.7) |
| Hand grip (kg) | 1 | 6.25 | 0.46 | (-10.3, 22.8) | 5.16 | 0.52 | (-10.4, 20.7) |
|  | 2 | -0.04 | 0.55 | (-0.16, 0.09) | -0.00 | 0.98 | (-0.11, 0.11) |
|  | 3 | **0.02** | **0.007** | **(0.00, 0.03)** | 0.01 | 0.26 | (-0.01, 0.02) |
|  | 4 | -3.62 | 0.19 | (-9.05, 1.81) | 1.34 | 0.65 | (-4.33, 7.01) |
|  | 5 | **-0.04** | **0.008** | **(-0.07, 0.00)** | 0.00 | 0.82 | (-0.02, 0.03) |
|  | 6 | 4.57 | 0.13 | (-1.37, 10.5) | 4.55 | 0.12 | (-1.25, 10.4) |
| Waist circumference (cm) | 1 | **21.3** | **0.02** | **(3.78, 38.8)** | **22.8** | **0.009** | **(5.73, 39.9)** |
|  | 2 | 0.07 | 0.32 | (-0.06, 0.19) | **0.12** | **0.04** | (0.01, 0.24) |
|  | 3 | 0.00 | 0.79 | (-0.00, 0.00) | 0.00 | 0.69 | (-0.00, 0.00) |
|  | 4 | **-6.62** | **0.01** | **(-11.9, -1.32)** | -4.60 | 0.17 | (-9.89, 1.76) |
|  | 5 | **-0.04** | **0.004** | **(-0.07, -0.01)** | -0.00 | 0.90 | (-0.03, 0.03) |
|  | 6 | -4.47 | 0.14 | (-10.5, 1.54) | **-6.15** | **0.04** | **(-12.1, -0.22)** |
| Waist:Hip ratio | 1 | 0.09 | 0.54 | (-0.19, 0.37) | 0.11 | 0.46 | (-0.18, 0.39) |
|  | 2 | 0.00 | 0.17 | (-0.00, 0.003) | 0.00 | 0.19 | (-0.00, 0.003) |
|  | 3 | **0.03** | **<0.001** | **(0.02, 0.04)** | **0.02** | **0.003** | **(0.01, 0.03)** |
|  | 4 | 0.09 | 0.07 | (-0.01, 0.19) | 0.01 | 0.92 | (-0.10, 0.11) |
|  | 5 | **0.01** | **0.02** | **(0.00, 0.01)** | 0.00 | 0.49 | (-0.00, 0.001) |
|  | 6 | 0.02 | 0.76 | (-0.13, 0.09) | -0.04 | 0.51 | (-0.15, 0.06) |
| Lean mass index (BIA) | 1 | 2.66 | 0.50 | (-5.15, 19.5) | 0.88 | 0.83 | (-7.22, 8.97) |
|  | 2 | -0.10 | 0.71 | (-0.06, 0.04) | -0.01 | 0.64 | (-0.07, 0.04) |
|  | 3 | 0.00 | 0.56 | (-0.00, 0.01) | -0.00 | 0.66 | (-0.01, 0.00) |
|  | 4 | -2.36 | 0.08 | (-5.03, 0.32) | -1.58 | 0.31 | (-4.66, 1.49) |
|  | 5 | -0.01 | 0.12 | (-0.02, 0.003) | 0.00 | 0.95 | (-0.01, 0.01) |
|  | 6 | -1.18 | 0.43 | (-4.15, 1.79) | -0.69 | 0.66 | (-3.82, 2.43) |
| Fat mass index (BIA) | 1 | 0.53 | 0.84 | (-4.76, 5.82) | 2.02 | 0.47 | (-3.46, 7.49) |
|  | 2 | -0.00 | 0.89 | (-0.04, 0.03) | 0.01 | 0.54 | (-0.03, 0.05) |
|  | 3 | **0.01** | **0.04** | **(0.00, 0.01)** | 0.00 | 0.09 | (-0.00, 0.01) |
|  | 4 | 0.08 | 0.93 | (-1.69, 1.85) | -0.03 | 0.97 | (-2.06, 1.99) |
|  | 5 | -0.01 | 0.21 | (-0.13, 0.003) | -0.00 | 0.65 | (-0.01, 0.01) |
|  | 6 | 0.08 | 0.94 | (-1.89, 2.04) | -0.94 | 0.37 | (-2.99, 1.11) |
| HAZ | 1 | **8.11** | **0.002** | **(2.89, 13.3)** | **6.62** | **0.02** | **(1.31, 119)** |
|  | 2 | **0.04** | **0.03** | **(0.004, 0.08)** | 0.03 | 0.13 | (-0.01, 0.06) |
|  | 3 | 0.001 | 0.64 | (-0.00, 0.01) | **0.01** | **0.003** | **(0.00, 0.01)** |
|  | 4 | -0.32 | 0.72 | (-2.08, 1.44) | -0.12 | 0.90 | (-2.04, 1.81) |
|  | 5 | 0.01 | 0.06 | (-0.00, 0.02) | 0.00 | 0.34 | (-0.01, 0.01) |
|  | 6 | -0.64 | 0.51 | (-2.54, 1.26) | 0.32 | 0.75 | (-1.63, 2.28) |

Linear regression, both simple and multivariable, adjusting for age, sex and HIV status. Blood pressure outcomes are also adjusted for height. Predictor variable is continuous values for PMGr definitions.

#### Quintiles method

When the rate of PMGr was split into quintiles, there were few instances of association with NCD indicators, especially after adjustment for age, sex and HIV status (**Annex Table 2; Figures 5 and 6**). In general, faster PMGr after discharge was associated with lower blood pressure (PMGr4 crude difference −1.08, 95%CI −1.92 to −0.24, p=0.01; PMGr5 crude difference −0.92, 95% CI −1.75 to −0.09, p=0.03) and smaller waist circumference (PMGr4 crude difference −0.38, 95%CI −0.69 to −0.06, p=0.02; PMGr5 crude difference −0.47, 95%CI −0.82 to −0.12, p=0.01). After adjustment, only waist circumference was associted with PMGr defined by change in WAZ per day during treatment (adjusted difference 0.32, 95%CI 0.01 to 0.63, p-value 0.04) and change in g/day during treatment (adjusted difference 0.37, 95%CI 0.06 to 0.68, p-value 0.02) (**Figure 6**). This was similar to findings from the method with no categorisation of PMGr. More boxplots are presented in **Annex Figures 4a-h**)

**Figure 5:**
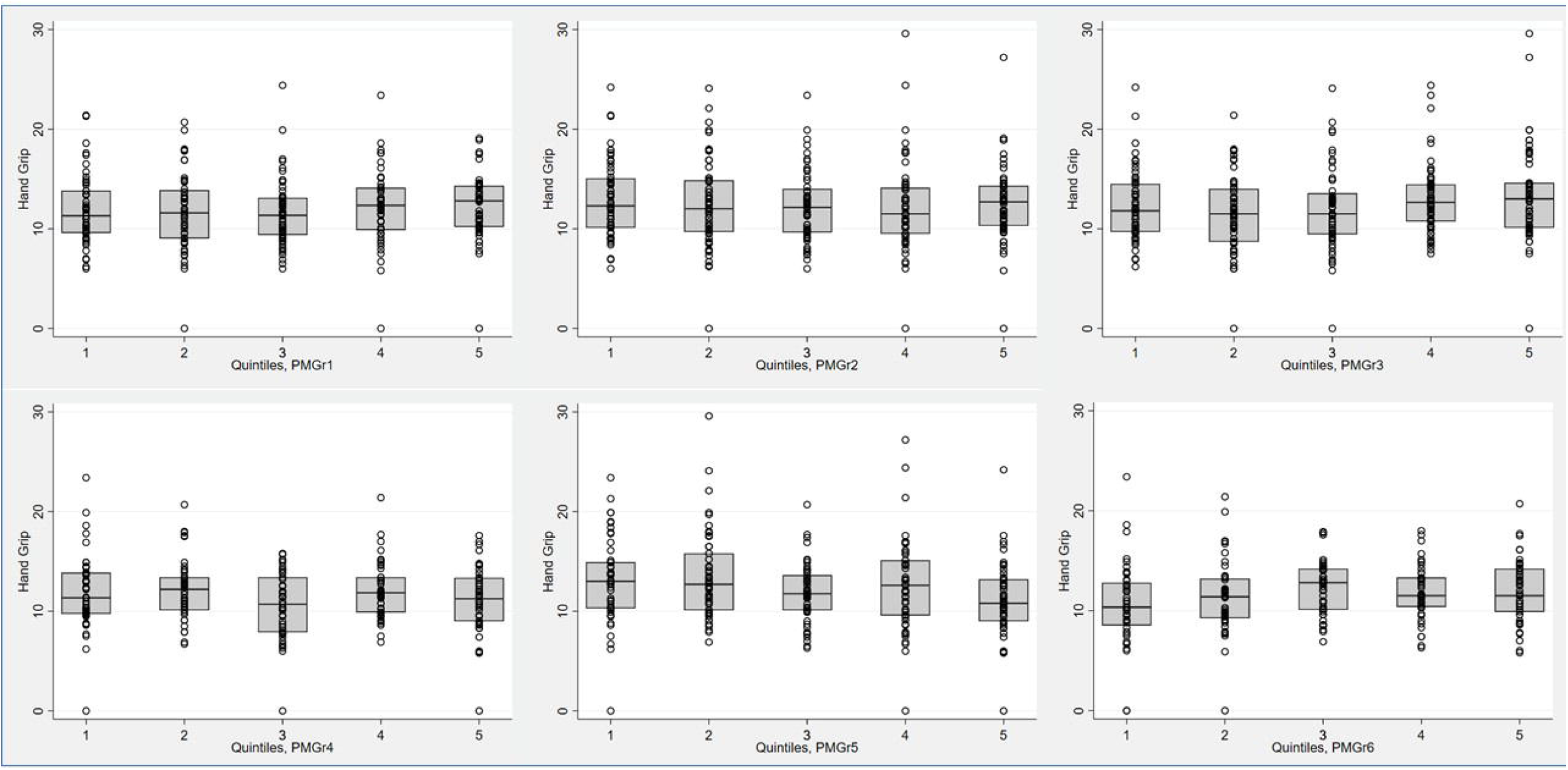
Boxplots for the association between each quintile of PMGr and handgrip strength, for the 6 PMGr definitions. Note quintile 1 is slowest and quintile 5 is fastest growth. PMGr1, 2 and 3 = during treatment, PMGr4, 5 and 6= from discharge to 1 year post-discharge. PMGr1= Δ WAZ per day, PMGr2= g/kg per day, PMGr3= g/day, PMGr4= Δ WAZ month, PMGr5= g/kg/month, PMGr6= Δ HAZ month

**Figure 6:**
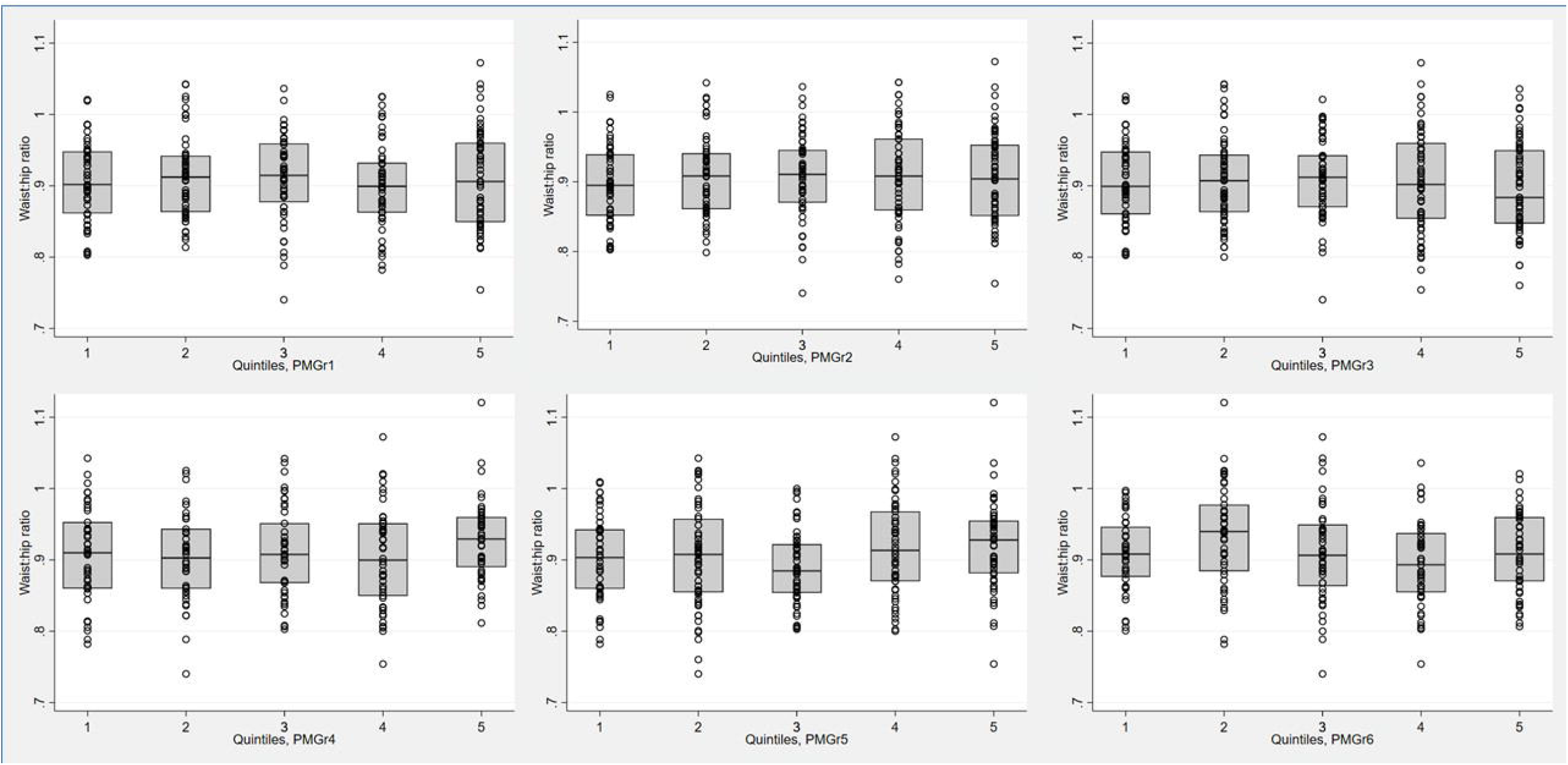
Boxplots for the association between each quintile of PMGr and waist circumference, for the 6 PMGr definitions. Note quintile 1 is slowest and quintile 5 is fastest growth. PMGr1, 2 and 3 = during treatment, PMGr4, 5 and 6= from discharge to 1 year post-discharge. PMGr1= Δ WAZ per day, PMGr2= g/kg per day, PMGr3= g/day, PMGr4= Δ WAZ month, PMGr5= g/kg/month, PMGr6= Δ HAZ month

#### Latent Class Analysis (LCA) method

The LCA method showed clear associations between growth patterns and NCD risk factors. Results from regression analysis of NCD indicators according to the latent classes are summarised in **Annex Table 3**. Latent class 1 had the lowest initial admission weight and the steepest change; class 5 had the lowest change in PMGr indicators. Faster PMGr was associated with lower systolic blood pressure (weight crude difference 2.37, 95%CI 1.38 to 3.36, p-value <0.001; HAZ crude difference 1.18, 95%CI 0.01 to 2.36, p=0.04), weaker hand grip strength (adjusted difference for WAZ: 0.58, weight: 0.99, HAZ: 0.96, all p<0.001) (**Figure 7**), smaller waist circumference (adjusted difference WAZ: 0.95, weight 1.16, HAZ: 1.22, all p<0.001), larger waist-hip ratio (adjusted difference WAZ: − 0.01, p<0.001, weight: −0.01, p=0.004, HAZ: −0.02, p<0.001) (**Figure 8**), more lean mass (adjusted difference WAZ: 0.29, p<0.001, weight: 0.42, p=0.002) and more fat mass (adjusted difference WAZ 0.17, p=0.002, weight: 0.19, p=0.03). No pattern of association with diastolic blood pressure was discernible (**all boxplots are in Annex Figures 5a-h**). The LCA classes highlight the large influence of admission anthropometry on the subsequent growth pattern, and on NCD risk factors (association between admission WAZ, weight (kg) and HAZ, and the NCD risk factors is presented in **Annex Table 4**). To try to adjust for the influence of initial admission WAZ, weight and HAZ on subsequent growth, we conducted an additional LCA after standardising initial admission WAZ/weight/ HAZ respectively, and found similar albeit slightly attenuated magnitudes of association (see **Annex Figure 6 and 7a-g**).

**Figure 7:**
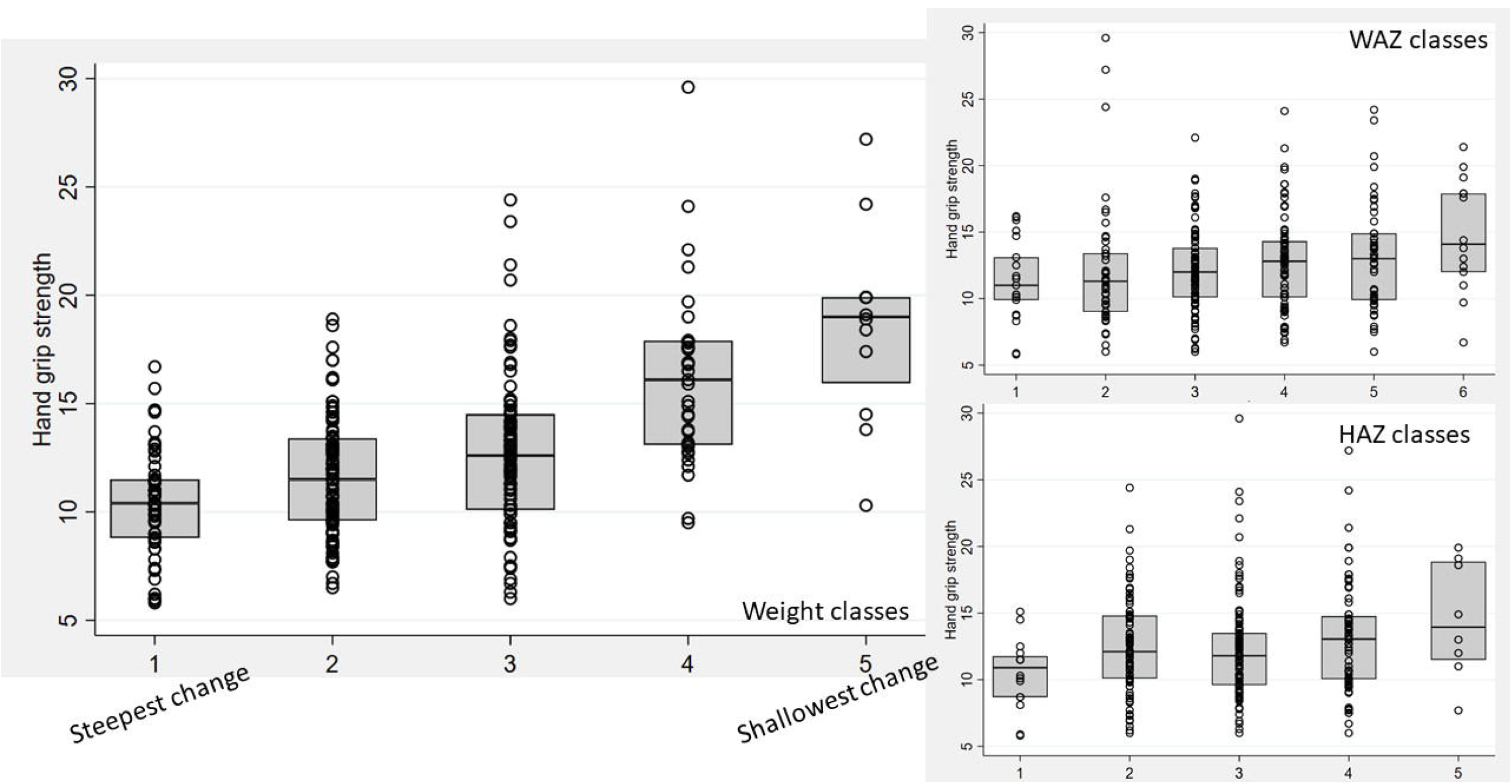
Boxplots for the association between LCA classes and hand grip strength, for weight, WAZ and HAZ. Note, class 1 have the lowest initial admission weight and the steepest change; class 5 have the lowest change in PMGr indicators

**Figure 8:**
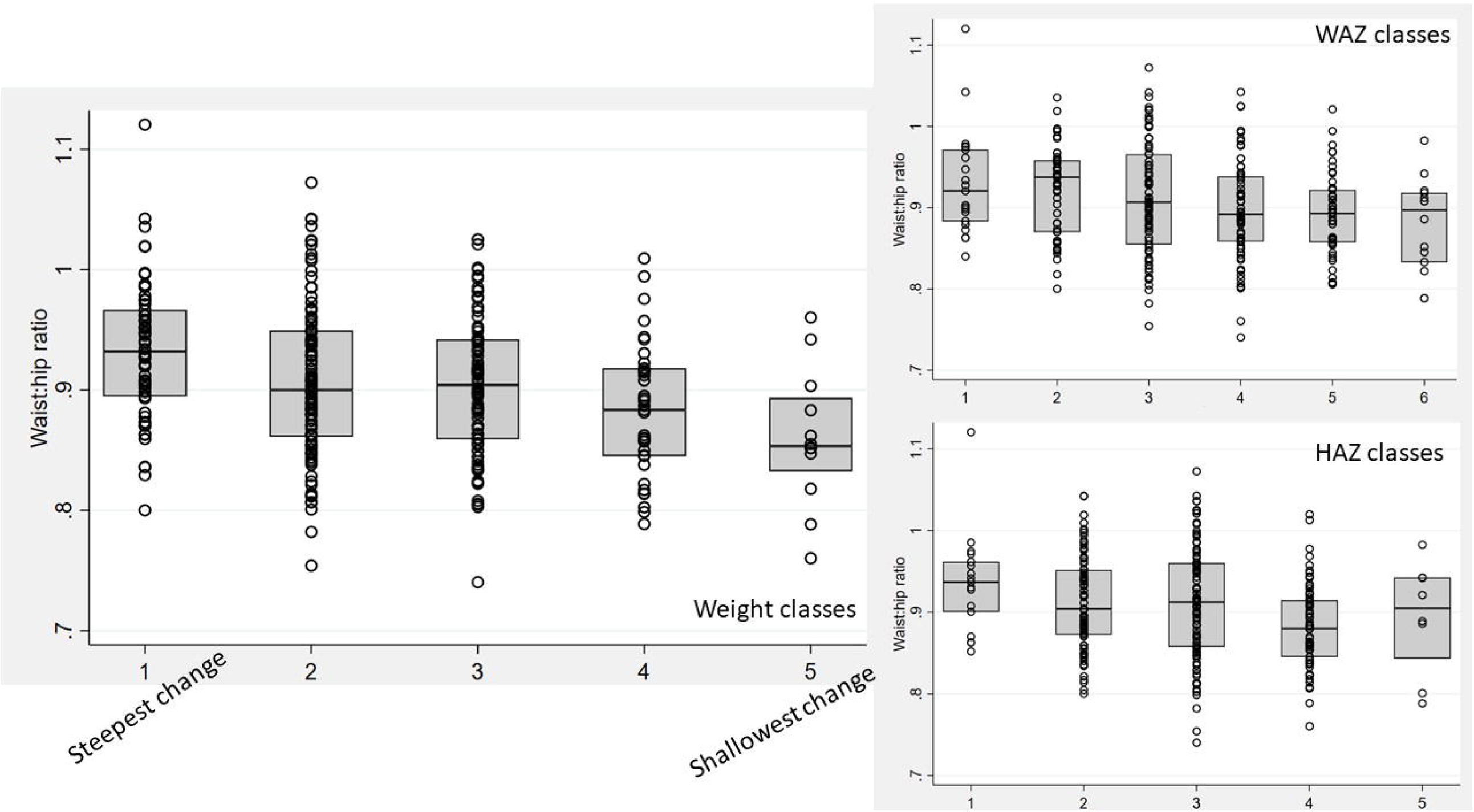
Boxplots for the association between LCA classes and waist:hip ratio, for weight, WAZ and HAZ. Note, class 1 have the lowest initial admission weight and the steepest change; class 5 have the lowest change in PMGr indicators

## Discussion

This exploratory analysis of methods for describing PMGr and its impact on later NCD risk factors is the first of its kind for survivors of severe malnutrition. We found that faster weight gain was associated with better odds of survival. Faster/steeper weight gain was also associated with stronger hand grip strength and larger HAZ 7 years post-discharge, both indicators of better health. However, there was also an association between faster PMGr and increased waist:hip ratio in later childhood, which can be an indicator of risk for later life heart disease and diabetes (28).

We found that the definition of PMGr had a large impact on how children are categorised and their associations with later health indicators. The clearest patterns were seen in the simplest definition based on the measure of g/day during treatment. Also, the analysis method had clear impact, the LCA method of categorising growth patterns showed the clearest effect. While the smooth regression (no categorisation) method and the quintiles method described the children’s growth velocity, the LCA method was more focused on the pattern of growth. The pattern seemed to have more of an influence on later health, and was highly influenced by the severity of anthropometric deficits at admission. It is very difficult to separate the risk of faster weight gain and greater weight deficits at admission to treatment, since the two are closely related (29). Those who start the smallest have the furthest to catch-up, plus there is also regression to the mean. Oedematous malnutrition is also a possible important confounder in our cohort since oedematous admission are less severely wasted at admission and tend to have lower weight gain. It is also less frequently associated with low birth weight and fewer long-term consequences (30). In an observational study such as ours it is not possible to disentangle the independent impact of these factors.

There may be some difference in NCD associations based on the timing of PMGr. Faster in-treatment growth (definitions 1-3) is associated with larger waist circumference whereas faster post-treatment growth (definitions 4-6) is associated with smaller waist circumference. The LCA patterns of growth show that the greatest changes in weight and height happen during the post-treatment period. It is important to note that larger waist:hip ratio in these survivors, and in children generally, may not have the same meaning as when applied to adults in HICs (31). Most of the evidence that associates larger waist:hip ratio with NCDs relates to the greater deposition of fat on the core body, close to vital organs (i.e., waist) rather than on the peripheral body (i.e., hips) (32-34). Previous analyses of this data found that survivors of severe malnutrition tended to have smaller waists than controls, but even smaller hips. This results in larger waist:hip ratio, but rather than having extra fat on their waists they appear to lack gluteofemoral fat (23). This may still have adverse health effects, as both peripheral fat and peripheral lean mass (i.e. larger hip circumference) are thought to be protective against NCDs, both of which are lacking in SAM survivors generally and especially in those with the steepest PMGr patterns (35). Fat distribution and proportion of visceral fat was also one of the key outcomes associated with rapid PMGr in a related analysis of adult SAM survivors in Jamaica (36).

Other studies that have described PMGr and its associations with NCD risk factors have largely focused on prenatal or very early postnatal nutrition. Our definitions of PMGr were influenced by this literature as well as indicators currently used in severe malnutrition programming. For example, LCA of WAZ has been used to identify growth profiles in low birth weight and small-for-gestational age (SGA) infants (37, 38). We include a measure of linear growth since “catch up growth” is commonly defined in SGA infants as height velocity (39). Other commonly used definitions of rapid weight gain or rapid catch-up growth in the literature include a change of WAZ>0.67 or crossing of one centile line on a growth chart, both of which have been associated with later life obesity when it occurs during infancy (40). However, neither of these could be used in relation to those treated for severe malnutrition since all children exceed these definitions.

Another definition we included was based on weight gain in g/kg/day, as this is often used in severe malnutrition programming. Standards for humanitarian programming recommend that “good” weight gain should be at least 10g/kg/day (41, 42). This is difficult to achieve for many programmes – the average in this dataset was 5.31g/kg/day. Such targets are not based on any hard evidence regarding healthiest rate of weight gain(43). Since the advent of outpatient-focused Community Management of Acute Malnutrition (CMAM) treatment programmes, weight gain per day has been overall slower than in old-style inpatient-only programmes (44). This is observed in spite of CMAM programme using high energy, (high-fat, high sugar) Ready-to-Use Therapeutic Foods (RUTF). Reasons for this include longer in-programme stays. There is currently no evidence that the ‘type of weight gain’ (i.e. fat vs lean) is excessively fatty (45, 46). Our results suggest that both severity at admission and rate of weight gain needs to be considered in future studies. Rate of in-programme gain can be easily controlled by food rations given at different energy doses (WHO recommends a range, although most programmes give doses at the upper end of this range(47)). Admission weight deficit is less easy to control – though this finding does reiterate the need for prevention and proactive case finding to catch children early in their deterioration. Programmes for moderate malnutrition are important in this prevention role, limiting and monitoring further weight loss (48).

Experimental studies on diet in early infancy, such as breastfed vs formula-fed infants, or use of formula milks with a range of calorie or protein contents, suggest a causal link between early growth acceleration and childhood risk-markers for NCDs (49). However, as in these results, they have also found some benefits of rapid infant weight gain, such as better neurodevelopment; and the risk-benefit balance is tipped in favour of the promotion of rapid growth in infants born preterm (50, 51). This level of experimental evidence to deduce where the balance of risks and benefits lie needs to be urgently replicated for children being treated for severe malnutrition. Recent studies exploring simplified protocols with reduced dosages of RUTF have observed slower weight gain but without any obvious ‘penalty’ in terms of increased mortality or reduced recovery rate from malnutrition (52-54). In most settings globally, child mortality is falling and it may be that even in this most vulnerable group of severely malnourished children, the optimal risk/benefit balance of rapid PMGr is different today compared to even 10 years ago.

### Limitations

Among the key limitations of our secondary analysis is the impact of survivor bias. More than half of the original cohort are known to have died either during or since treatment. These were generally children who had slow initial in-programme growth. Had they lived they may have changed our results in either direction: they may have had more or less NCD risk than the ‘healthier’ survivors. Our results are also limited by the relatively young age at which NCD risk was assessed (median age 11 years). It could be that metabolic loads are not yet sufficient to manifest as risk factors for NCD. Both risk factors and NCDs themselves will only become apparent as the cohort ages. Also important to acknowledge is that a risk factor for NCD does not necessarily mean that the NCD itself will develop (55). Related to this, risk factors established in and applicable to high-income western populations may not apply equally to populations such as ours in Malawi(56).

Since this is an observational analysis in which all children received similar diets, we cannot say from our data what would have happened had growth rate been controlled by different caloric regimes. There are many other factors affecting weight gain that are not controlled for, such as breastfeeding status, diarrhoea and vomiting during treatment, and other concurrent illnesses and infections which would affect weight gain and survival (57). Weight gain differences may also reflect individual variations in general health status and resilience to illness episodes. Lastly, our results may not be generalisable to other setting, especially to those at a different stage of the nutrition transition, since the nutrition environment post-discharge is likely to have a large influence on later NCD risk factors (58, 59).

Addressing these limitations, we call for future intervention trials where different doses and types of therapeutic food for severe malnutrition are compared and where long term as well as short term benefits as well as risks are assessed. Valid measures, markers and biomarkers of long term NCD risk are particularly important to develop since it is unlikely that any study will be able to follow-up children to the point where NCDs are directly measurable.

## Conclusion

Our study paints a complex picture of benefits and risks associated with faster PMGr. Different definitions of PMGr show different relationships with the outcomes, and our results indicate that simple measures of weight gain (g/day), initial growth deficit, and the pattern of change have important implications for future health and NCDs. To research this topic in the future, better understanding of NCD risk markers and biomarkers are needed; peripheral vs visceral weight distribution in particular requires further exploration. Observational studies are limited in what they can deduce; intervention trials are needed.

## Supporting information

Annexes see accompanying PowerPoint slides

## Data Availability

All data produced in the present study are available upon reasonable request to the authors via the LSHTM Data Repository

https://datacompass.lshtm.ac.uk/id/eprint/1248/

**Annexes – see accompanying PowerPoint slides**

