## Supplementary material for "Post-malnutrition growth and its associations with child survival and non-communicable disease risk: A secondary analysis of the Malawi ‘ChroSAM’ cohort": Annexes see accompanying PowerPoint slides

### Annex Tables and Figures

Chrosam CHANGE study analysis

### Annex Table 1: sample size for LCA-derived PMGr classes

| LCA classes | WAZ LCA<br>N (%) | Weight LCA<br>N (%) | HAZ LCA<br>N (%) |
|---|---|---|---|
| Class 1 | 21 (6.7) | 60 (18.8) | 17 (5.4) |
| Class 2 | 54 (17.1) | 111 (34.8) | 96 (30.2) |
| Class 3 | 78 (24.7) | 97 (30.4) | 121 (38.1) |
| Class 4 | 53 (16.8) | 38 (11.9) | 75 (23.6) |
| Class 5 | 16 (5.1) | 13 (4.1) | 8 (2.5) |
| Class 6 | 94 (29.8) | N/A | N/A |

### Annex Figure 1: mean WAZ, weight, and HAZ over time

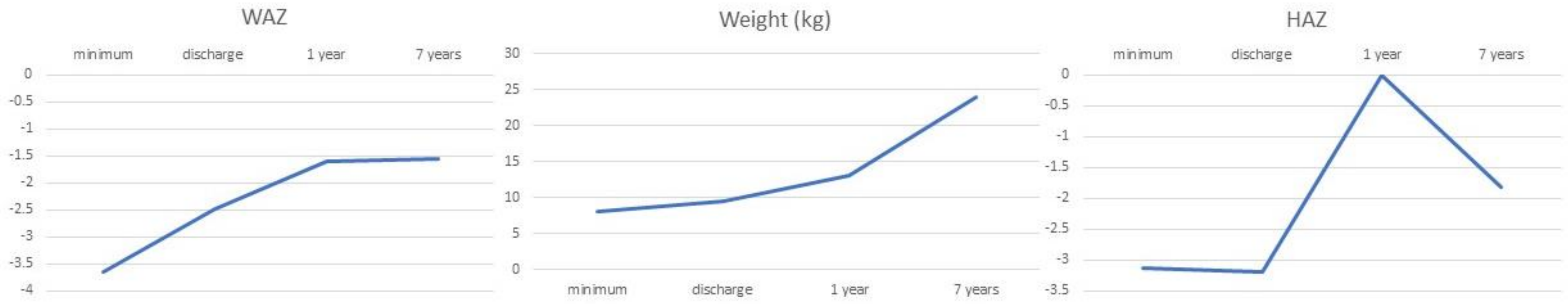

### Annex Figure 2: Mean quintile weight-for-age z-scores (WAZ) according to five post-malnutrition growth rate definition (PMGr) at four (non-uniformly spaced) time point

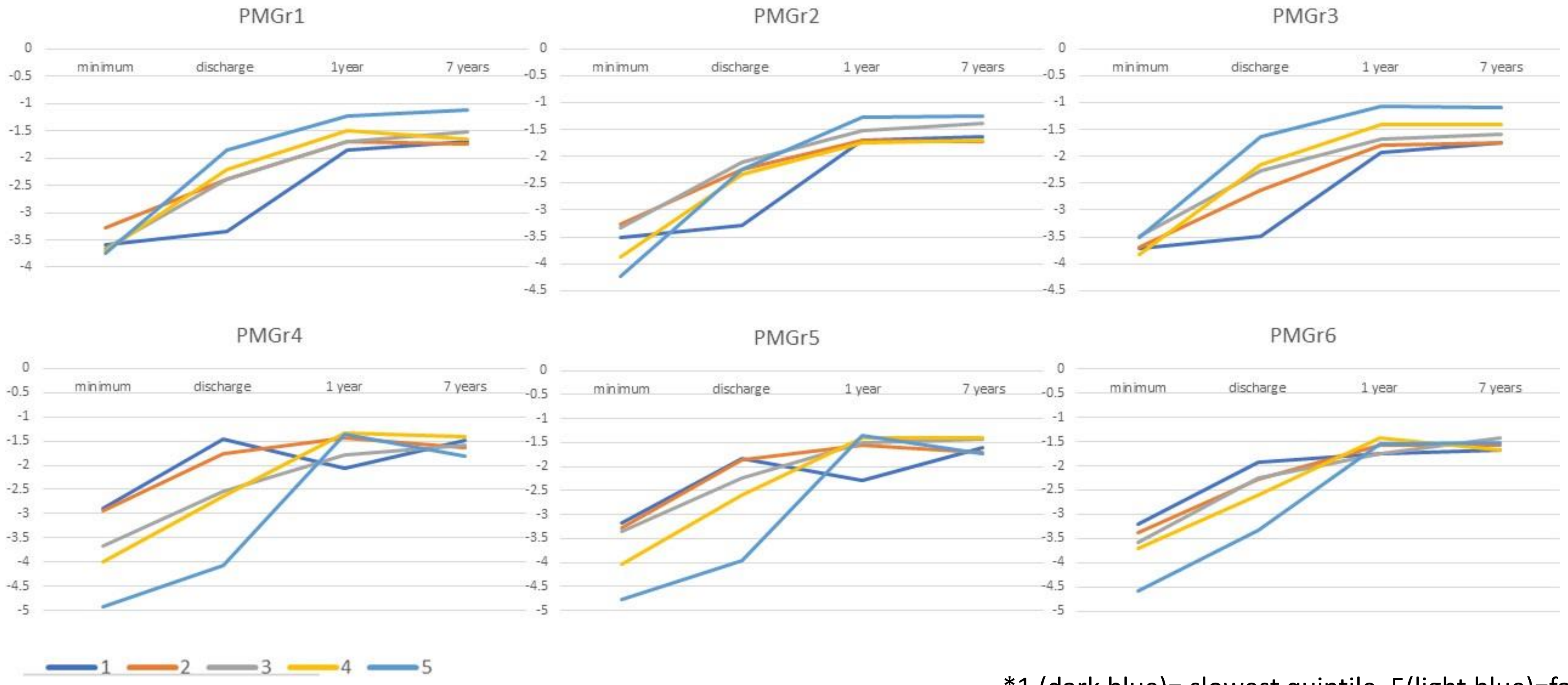


*1 (dark blue)= slowest quintile, 5(light blue)=fastest quintile

### Annex Table 2: Association between PMGr definition quintiles and NCD outcomes

| NCD indicator | PMGr definition | Unadjusted coefficient | P-value | 95% CI | Adjusted coefficient | P-value | 95% CI |
|---|---|---|---|---|---|---|---|
| Systolic BP | 1 | 0.368 | 0.349 | (-0.40, 1.14) | -0.061 | 0.877 | (-0.84, 0.72) |
| | 2 | 0.010 | 0.979 | (-0.77, 0.79) | -0.072 | 0.847 | (-0.80, 0.66) |
| | 3 | 0.629 | 0.115 | (-0.15, 1.41) | -0.158 | 0.682 | (-0.92, 0.60) |
| | 4 | -1.081 | 0.012 | (-1.92, -0.24) | -0.703 | 0.113 | (-1.57, 0.17) |
| | 5 | -0.919 | 0.030 | (-1.75, -0.09) | -0.373 | 0.400 | (-1.25, 0.49) |
| | 6 | -0.164 | 0.703 | (-1.01, 0.68) | -0.360 | 0.390 | (-1.18, 0.46) |
| Diastolic BP | 1 | -0.247 | 0.506 | (-098, 0.48) | -0.349 | 0.373 | (-1.12, 0.42) |
| | 2 | -0.272 | 0.446 | (-0.97, 0.43) | -0.281 | 0.440 | (-0.99, 0.43) |
| | 3 | 0.026 | 0.943 | (-0.67, 0.73) | -0.234 | 0.535 | (-0.97, 0.51) |
| | 4 | -0.936 | 0.023 | (-1.74, -0.13) | -0.864 | 0.053 | (-1.74, 0.01) |
| | 5 | -0.685 | 0.073 | (-1.44, 0.07) | -0.689 | 0.117 | (-1.55, 0.17) |
| | 6 | 0.242 | 0.558 | (-0.57, 1.05) | -0.024 | 0.954 | (-0.86, 0.81) |
| Hand grip | 1 | 0.106 | 0.488 | (-0.19, 0.41) | 0.037 | 0.797 | (-0.25, 0.32) |
| | 2 | -0.100 | 0.543 | (-0.43, 0.22) | 0.000 | 0.999 | (-0.28, 0.28) |
| | 3 | 0.365 | 0.027 | (0.04, 0.69) | 0.098 | 0.508 | (-0.19, 0.39) |
| | 4 | -0.155 | 0.329 | (-0.47, 0.16) | 0.093 | 0.562 | (-0.22, 0.41) |
| | 5 | -0.459 | 0.010 | (-0.81, -0.11) | 0.079 | 0.643 | (-0.26, 0.42) |
| | 6 | 0.295 | 0.064 | (-0.12, 0.61) | 0.278 | 0.069 | (-0.02, 0.58) |
| Waist circumference | 1 | 0.332 | 0.042 | (0.01, 0.65) | 0.316 | 0.047 | (0.00, 0.63) |
| | 2 | 0.109 | 0.534 | (-0.24, 0.45) | 0.253 | 0.099 | (-0.05, 0.55) |
| | 3 | 0.604 | 0.001 | (0.27, 0.94) | 0.369 | 0.018 | (0.06, 0.68) |
| | 4 | -0.375 | 0.022 | (-0.69, -0.06) | -0.249 | 0.137 | (-0.58, 0.08) |
| | 5 | -0.474 | 0.008 | (-0.82, -0.12) | 0.059 | 0.733 | (-0.28, 0.39) |
| | 6 | -0.214 | 0.189 | (-0.54, 0.11) | -0.299 | 0.059 | (-0.61, 0.01) |

| NCD indicator | PMGr definition | Unadjusted coefficient | P-value | 95% CI | Adjusted coefficient | P-value | 95% CI |
|---|---|---|---|---|---|---|---|
| Waist:Hip ratio | 1 | -0.000 | 0.923 | (-0.01, 0.01) | 0.001 | 0.796 | (-0.00, 0.01) |
| | 2 | 0.002 | 0.441 | (-0.00, 0.01) | 0.002 | 0.382 | (-0.00, 0.01) |
| | 3 | -0.002 | 0.409 | (-0.01, 0.00) | 0.000 | 0.993 | (-0.01, 0.01) |
| | 4 | 0.004 | 0.155 | (-0.00, 0.01) | -0.000 | 0.989 | (-0.01, 0.01) |
| | 5 | 0.005 | 0.061 | (-0.00, 0.01) | 0.001 | 0.672 | (-0.00, 0.01) |
| | 6 | -0.004 | 0.182 | (-0.01, 0.00) | -0.004 | 0.147 | (-0.01, 0.00) |
| Lean mass index (BIA) | 1 | 0.138 | 0.056 | (-0.00, 0.28) | 0.109 | 0.143 | (-0.04, 0.26) |
| | 2 | 0.039 | 0.577 | (-0.10, 0.18) | 0.041 | 0.559 | (-0.09, 0.18) |
| | 3 | 0.136 | 0.057 | (-0.00, 0.28) | 0.067 | 0.356 | (-0.08, 0.21) |
| | 4 | -0.111 | 0.168 | (-0.27, 0.05) | -0.071 | 0.423 | (-0.24, 0.10) |
| | 5 | -0.141 | 0.067 | (-0.29, 0.01) | -0.015 | 0.860 | (-0.18, 0.15) |
| | 6 | -0.049 | 0.543 | (-0.21, 0.11) | -0.026 | 0.755 | (-0.19, 0.14) |
| Fat mass index (BIA) | 1 | -0.012 | 0.800 | (-0.11, 0.08) | 0.005 | 0.923 | (-0.09, 0.11) |
| | 2 | -0.024 | 0.623 | (-0.12, 0.07) | 0.006 | 0.901 | (-0.09, 0.09) |
| | 3 | 0.082 | 0.094 | (-0.01, 0.18) | 0.065 | 0.186 | (-0.03, 0.16) |
| | 4 | -0.019 | 0.711 | (-0.12, 0.08) | -0.033 | 0.568 | (-0.15, 0.08) |
| | 5 | -0.061 | 0.227 | (-0.16, 0.04) | -0.018 | 0.746 | (-0.13, 0.09) |
| | 6 | 0.051 | 0.337 | (-0.05, 0.16) | 0.006 | 0.910 | (-0.10, 0.11) |
| HAZ | 1 | 0.115 | 0.019 | (0.02, 0.21) | 0.091 | 0.066 | (-0.01, 0.19) |
| | 2 | 0.083 | 0.090 | (-0.01, 0.18) | 0.051 | 0.281 | (-0.04, 0.14) |
| | 3 | 0.049 | 0.310 | (-0.05, 0.15) | 0.097 | 0.05 | (0.00, 0.19) |
| | 4 | -0.023 | 0.663 | (-0.12, 0.08) | -0.013 | 0.819 | (-0.12, 0.09) |
| | 5 | 0.141 | 0.007 | (0.04, 0.24) | 0.089 | 0.114 | (-0.02, 0.20) |
| | 6 | -0.027 | 0.599 | (-0.13, 0.07) | 0.016 | 0.757 | (-0.09, 0.12) |

### Annex Figures 3a-1h

Scatterplots showing associations between ‘no categorised’ PMGr indicators and NCD risk outcomes

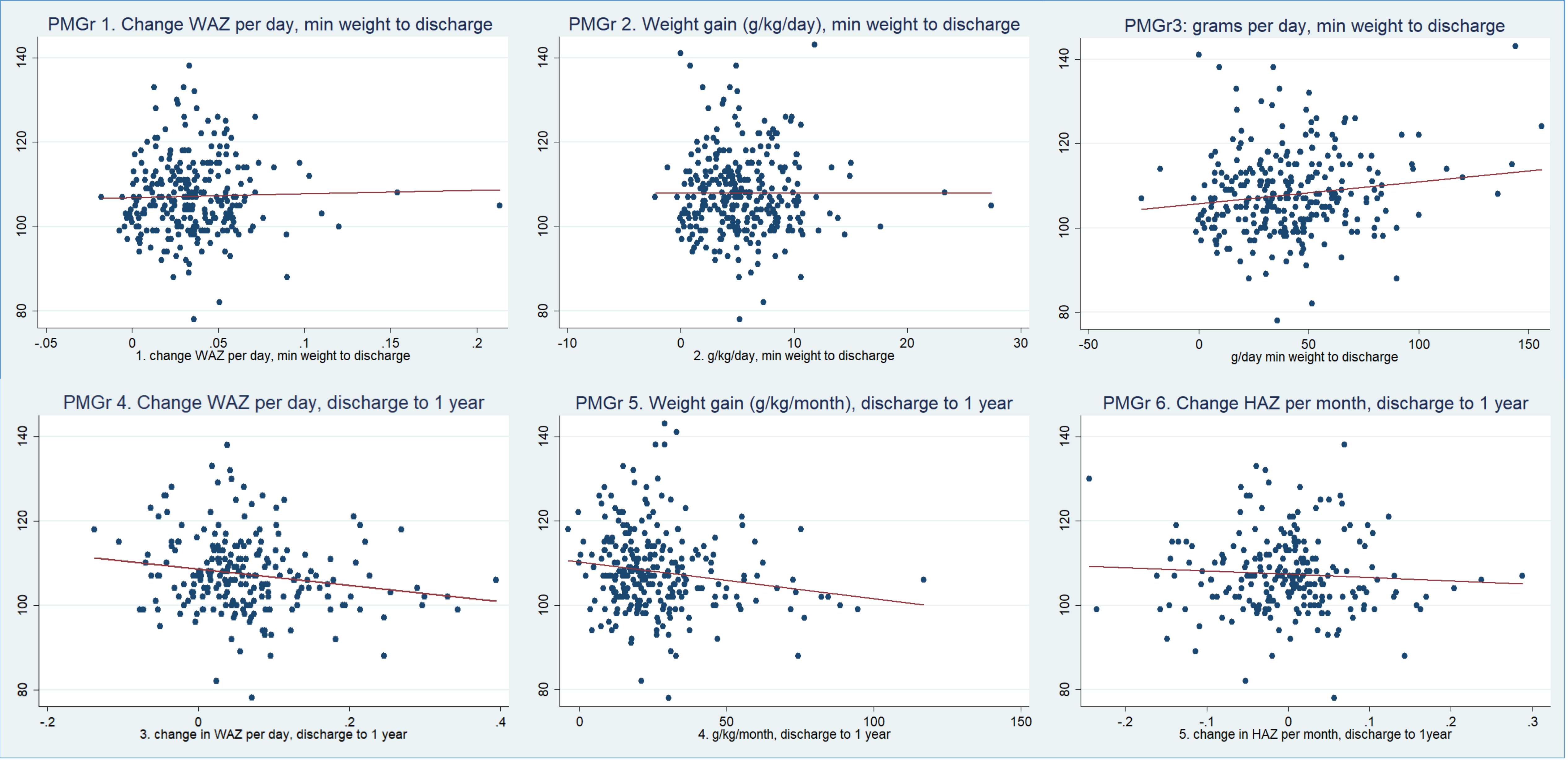


Systolic Blood Pressure

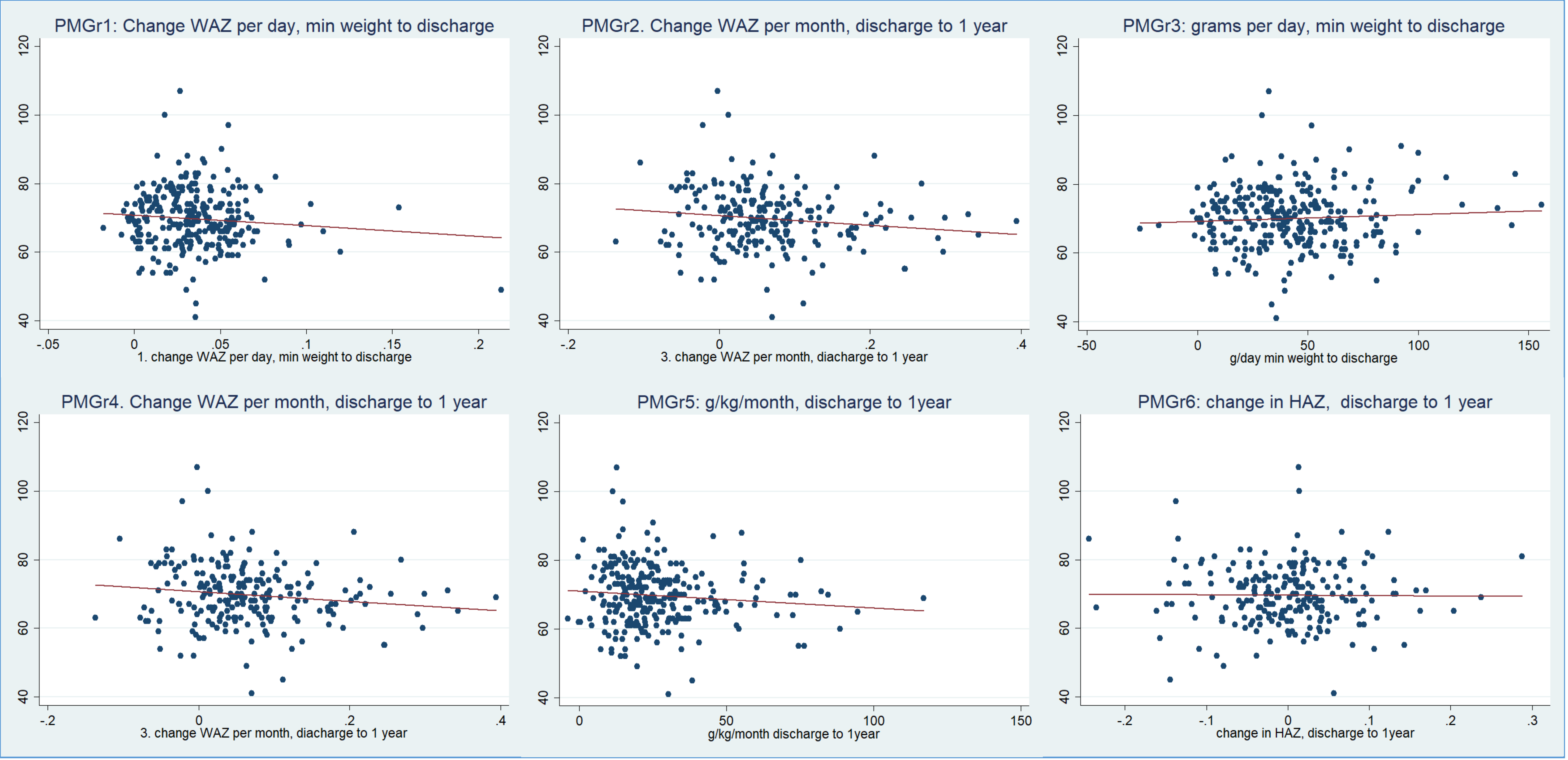


Diastolic Blood Pressure

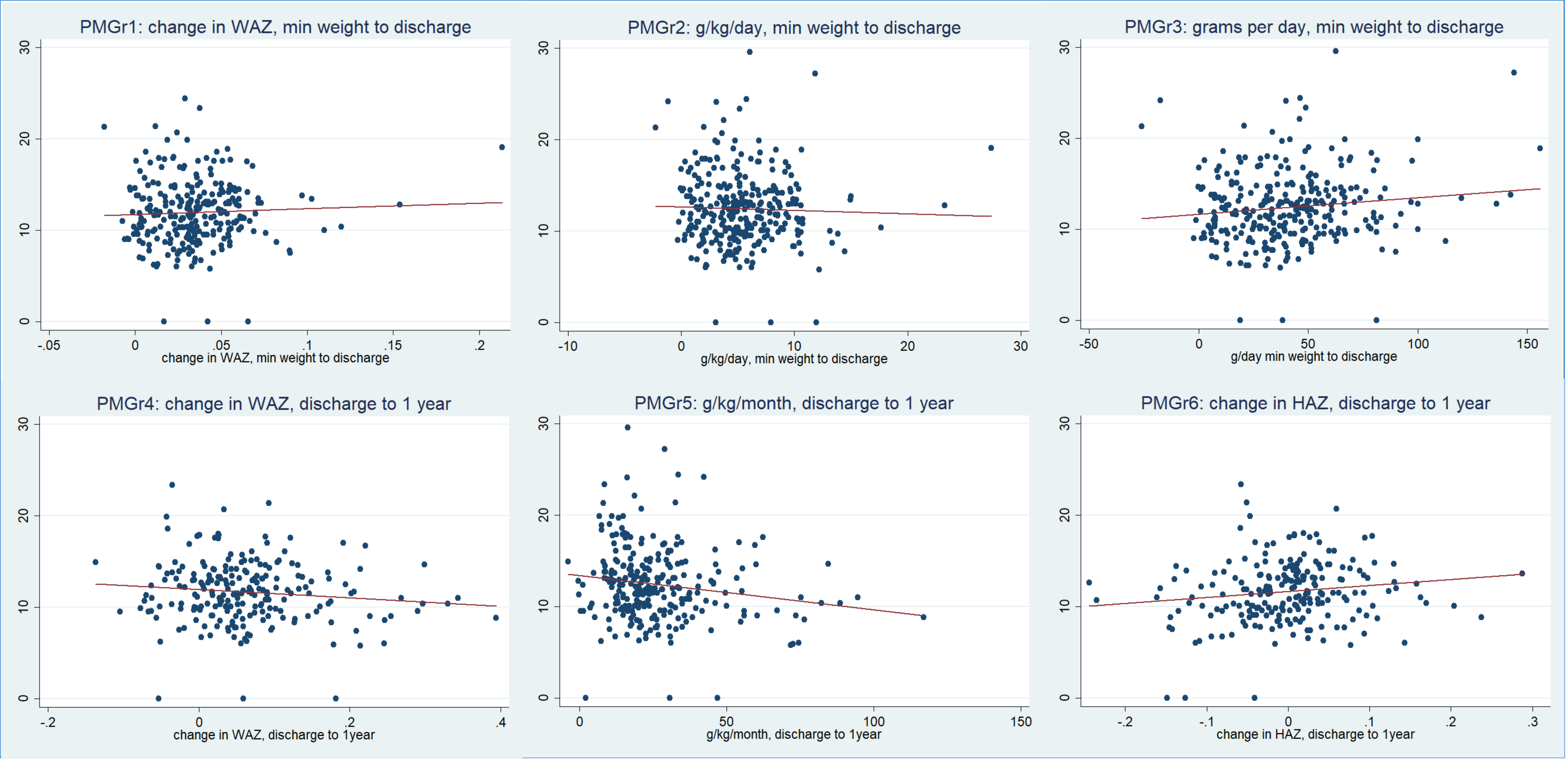


Hand grip strength

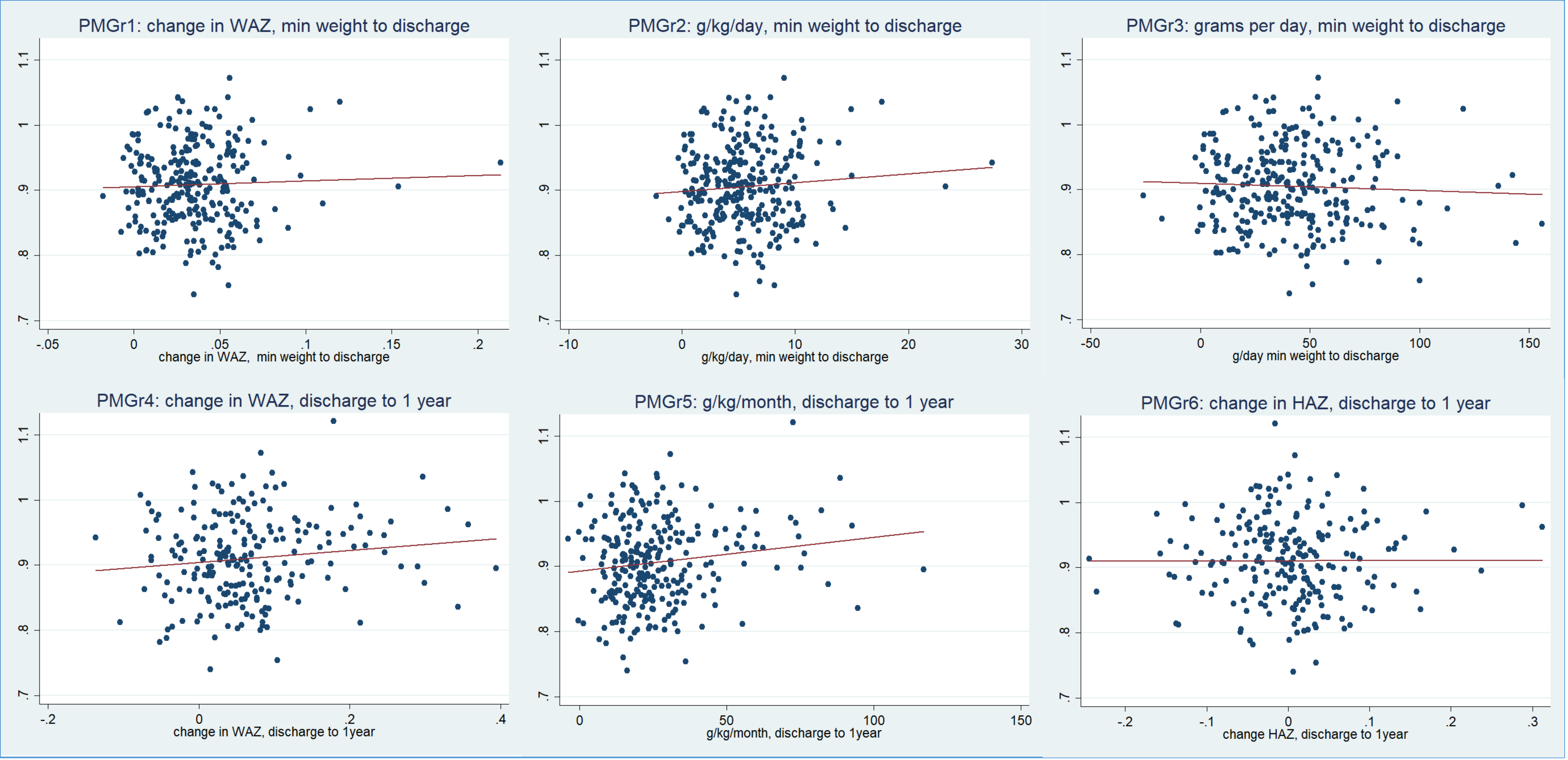


Waist/Hip ratio

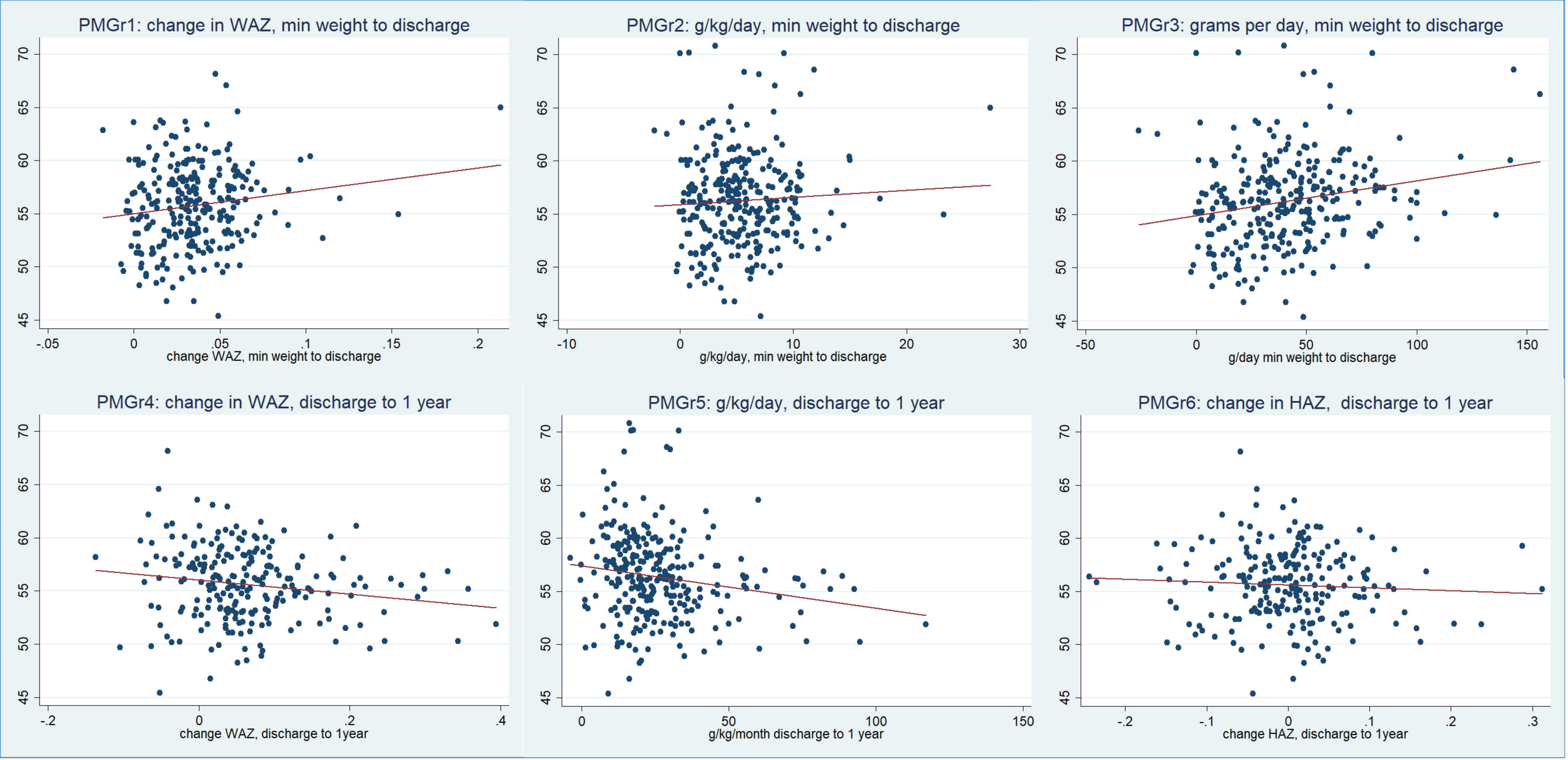


Waist circumference

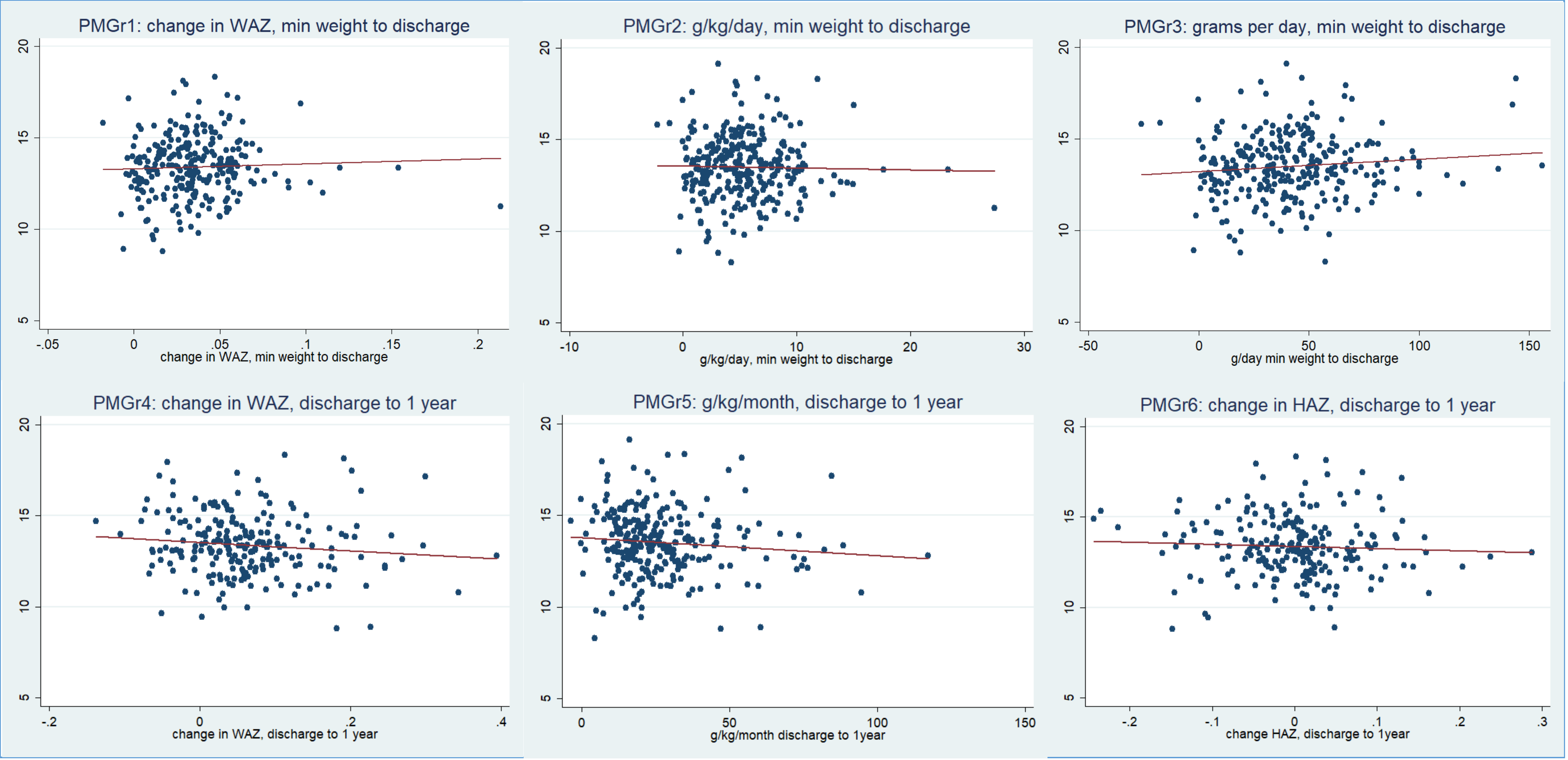


Lean Mass Index

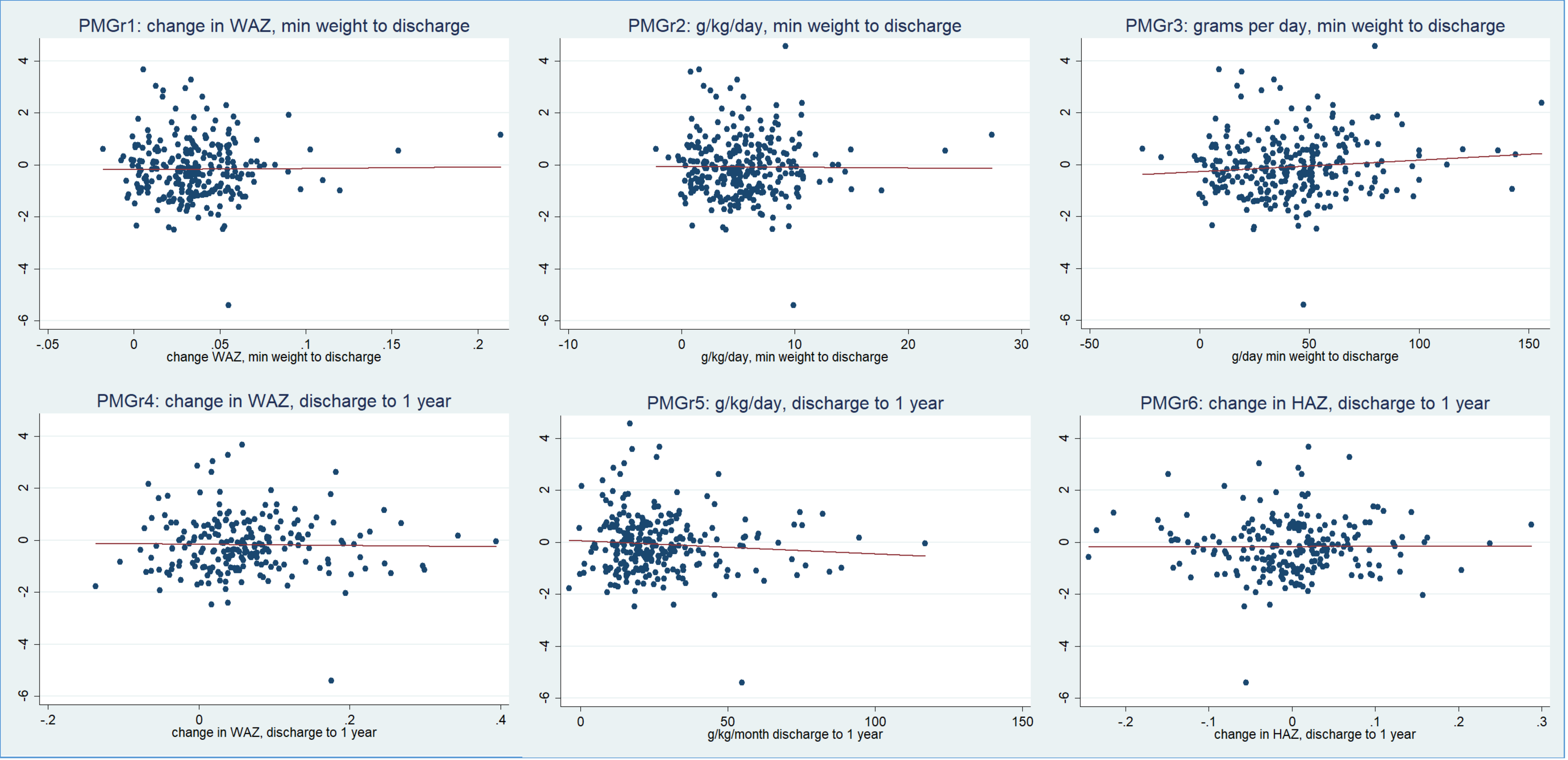


Fat Mass Index

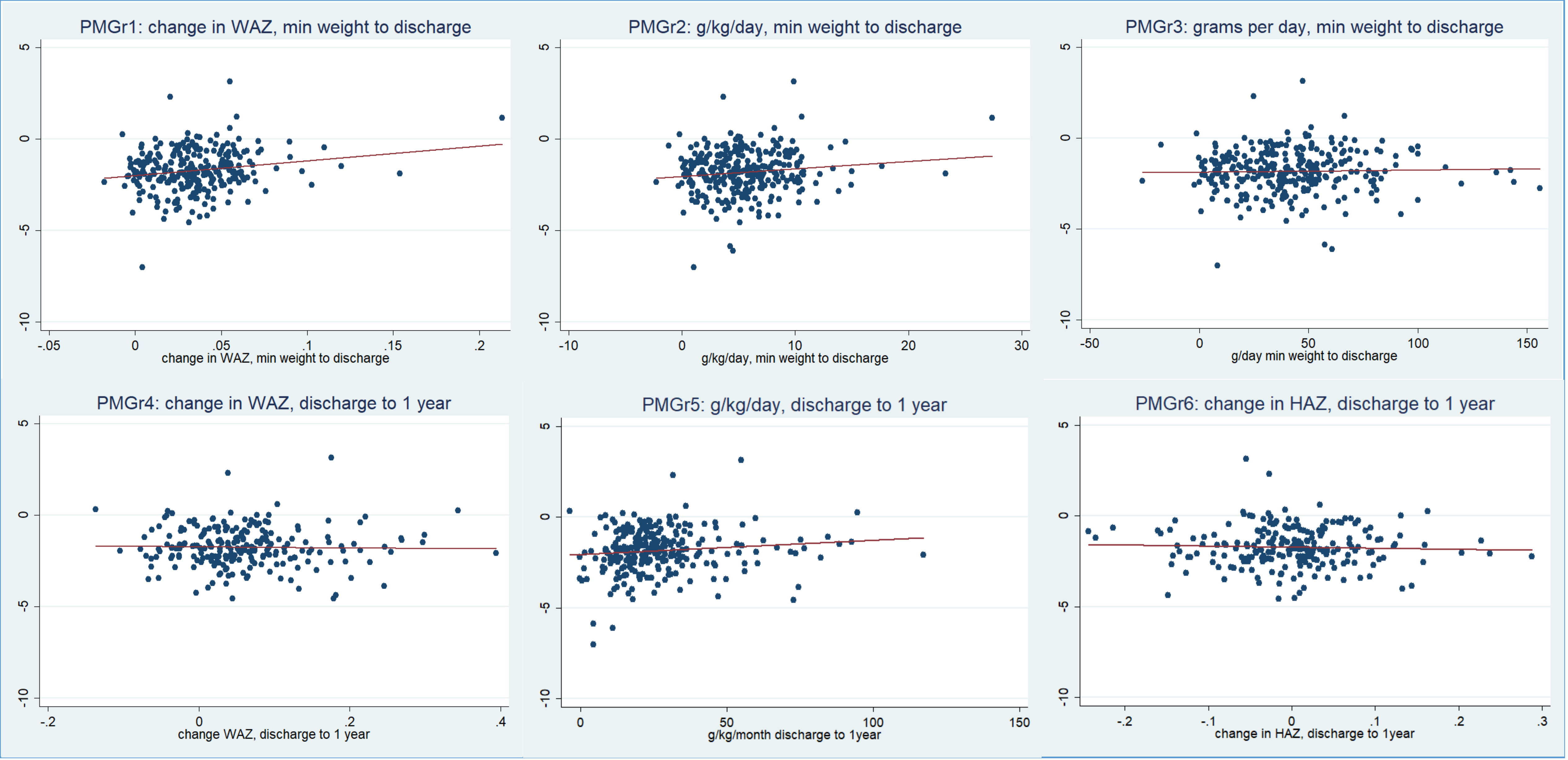
PMGr1: change in WAZ, min weight to discharge
5
0
-5
-10
-.05
0
.05
.1
.15
.2
change in WAZ, min weight to discharge
PMGr2: g/kg/day, min weight to discharge
5
0
-5
-10
-10
0
10
20
30
g/kg/day, min weight to discharge
PMGr3: grams per day, min weight to discharge
5
0
-5
-10
-50
0
50
100
150
g/day min weight to discharge
PMGr4: change in WAZ, discharge to 1 year
5
0
-5
-10
-.2
0
.2
.4
change WAZ, discharge to 1 year
PMGr5: g/kg/day, discharge to 1 year
5
0
-5
-10
0
50
100
150
g/kg/month discharge to 1year
PMGr6: change in HAZ, discharge to 1 year
5
0
-5
-10
-.2
-.1
0
.1
.2
.3
change in HAZ, discharge to 1year

HAZ

### Annex Figures 4a-2h

Boxplots showing associations between PMGr quintiles and NCD risk outcomes

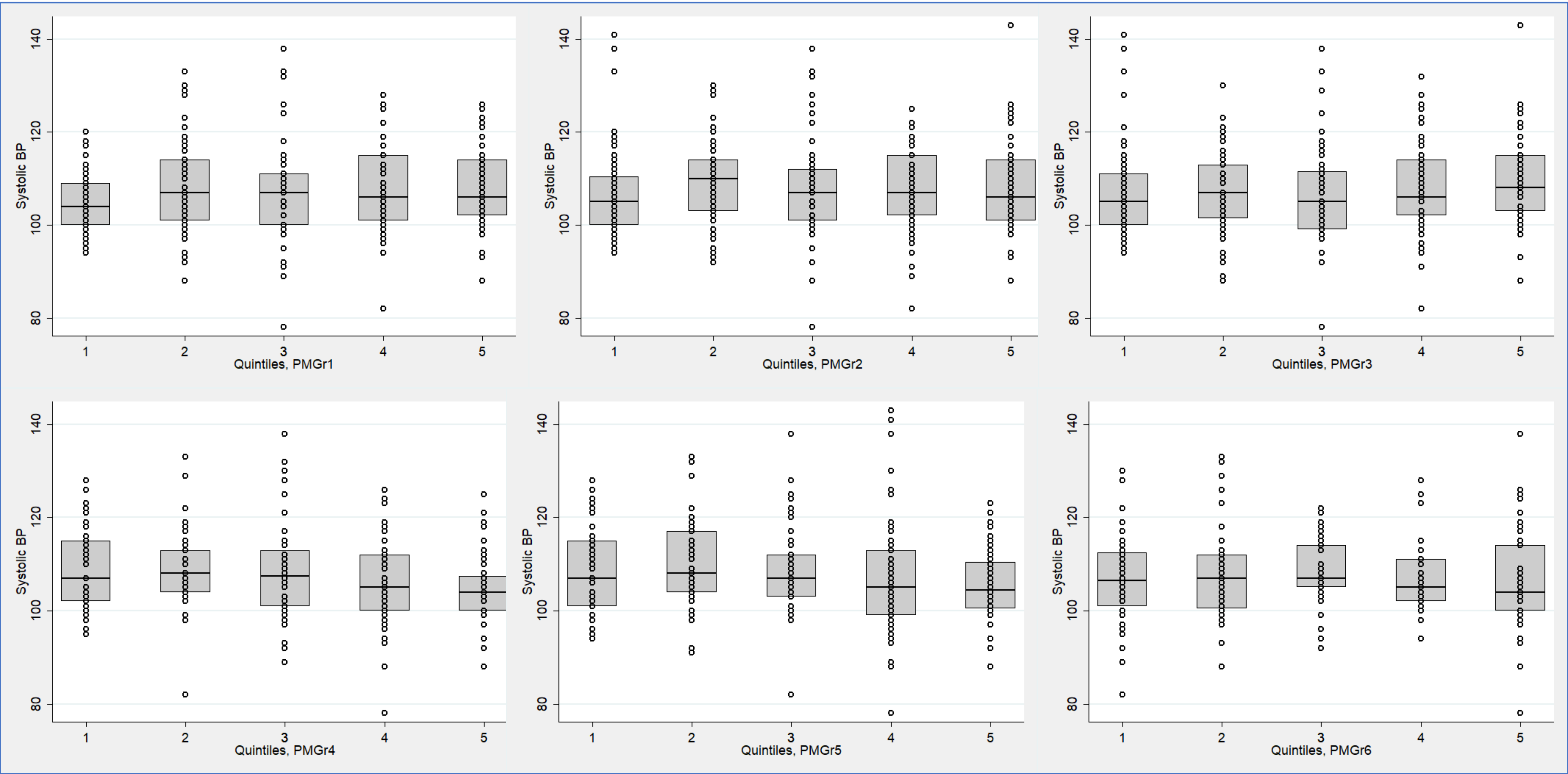
Systolic BP
140
120
100
80
1
2
3
4
5
Quintiles, PMGr1
Quintiles, PMGr2
Quintiles, PMGr3
Quintiles, PMGr4
Quintiles, PMGr5
Quintiles, PMGr6

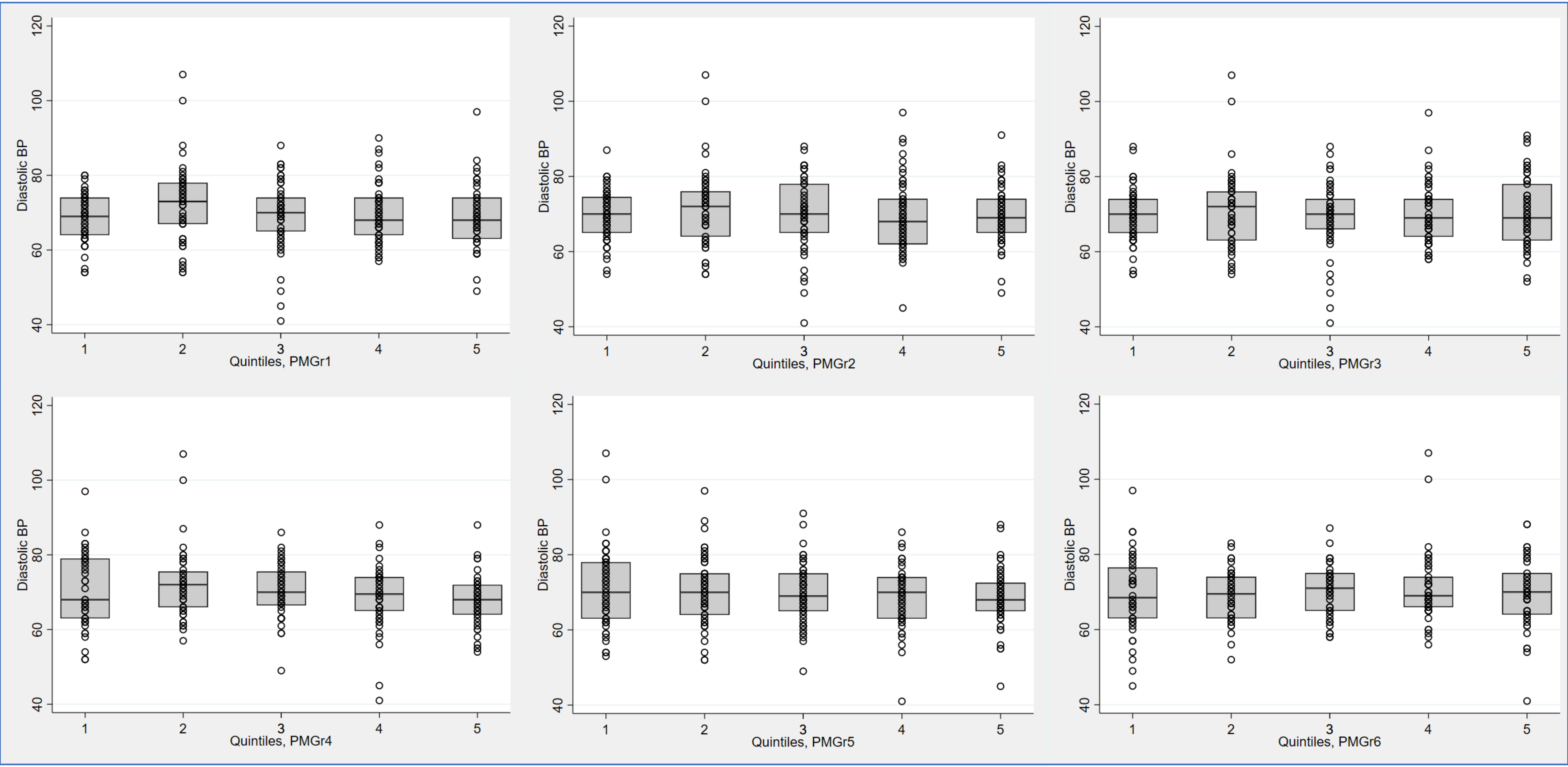
Diastolic BP
Quintiles, PMGr1
Quintiles, PMGr2
Quintiles, PMGr3
Quintiles, PMGr4
Quintiles, PMGr5
Quintiles, PMGr6

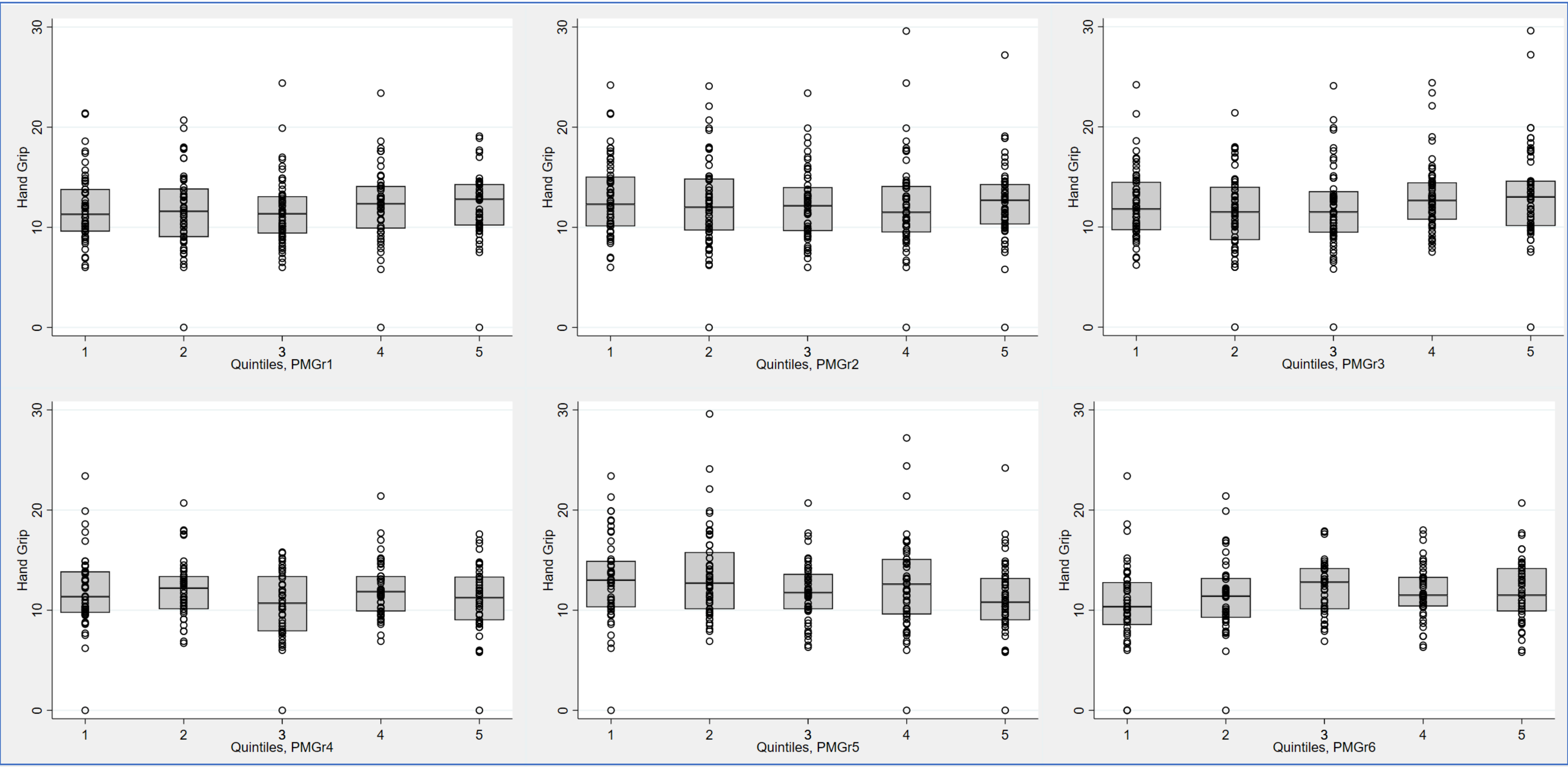
Hand Grip
Quintiles, PMGr1
Quintiles, PMGr2
Quintiles, PMGr3
Quintiles, PMGr4
Quintiles, PMGr5
Quintiles, PMGr6

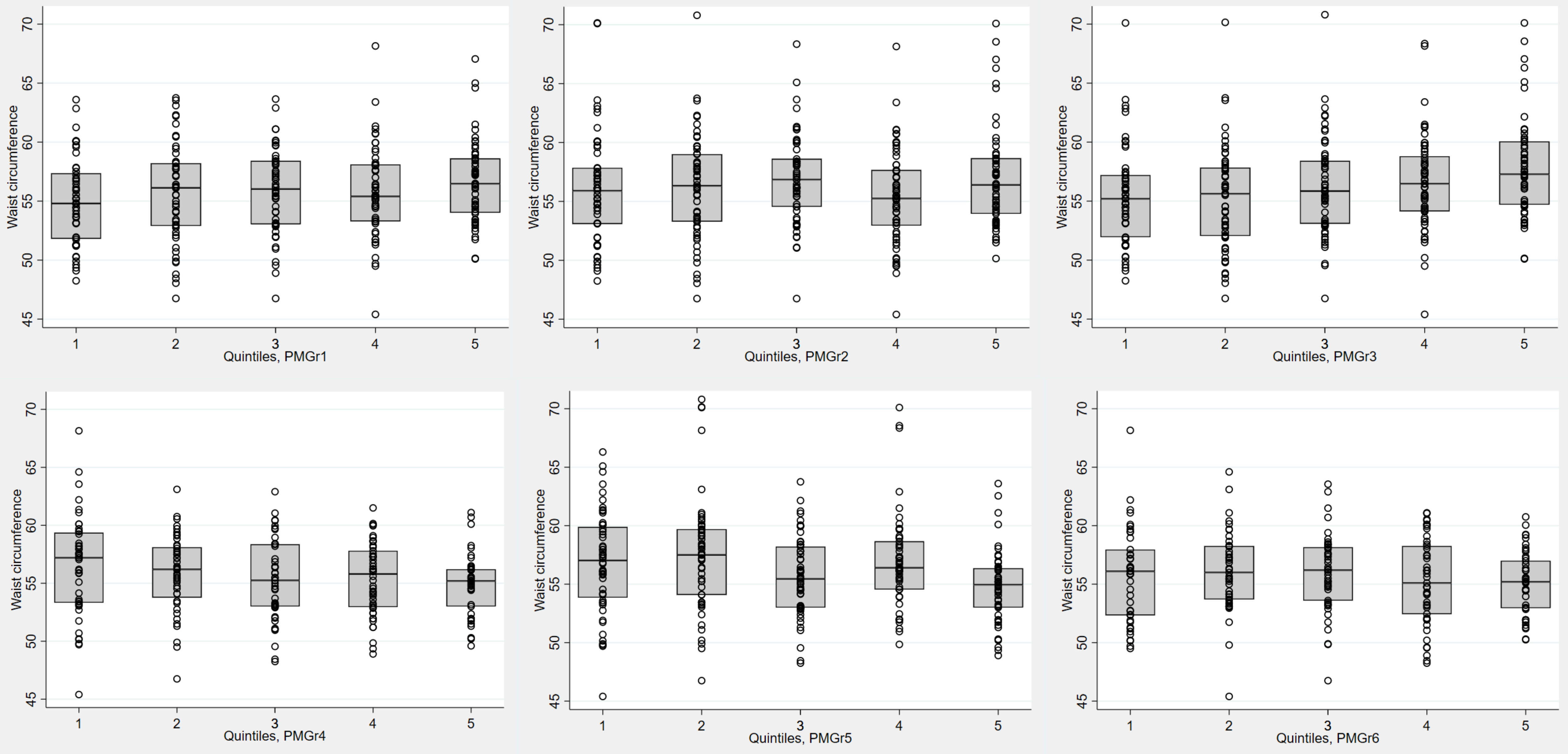
Waist circumference
Quintiles, PMGr1
Quintiles, PMGr2
Quintiles, PMGr3
Quintiles, PMGr4
Quintiles, PMGr5
Quintiles, PMGr6

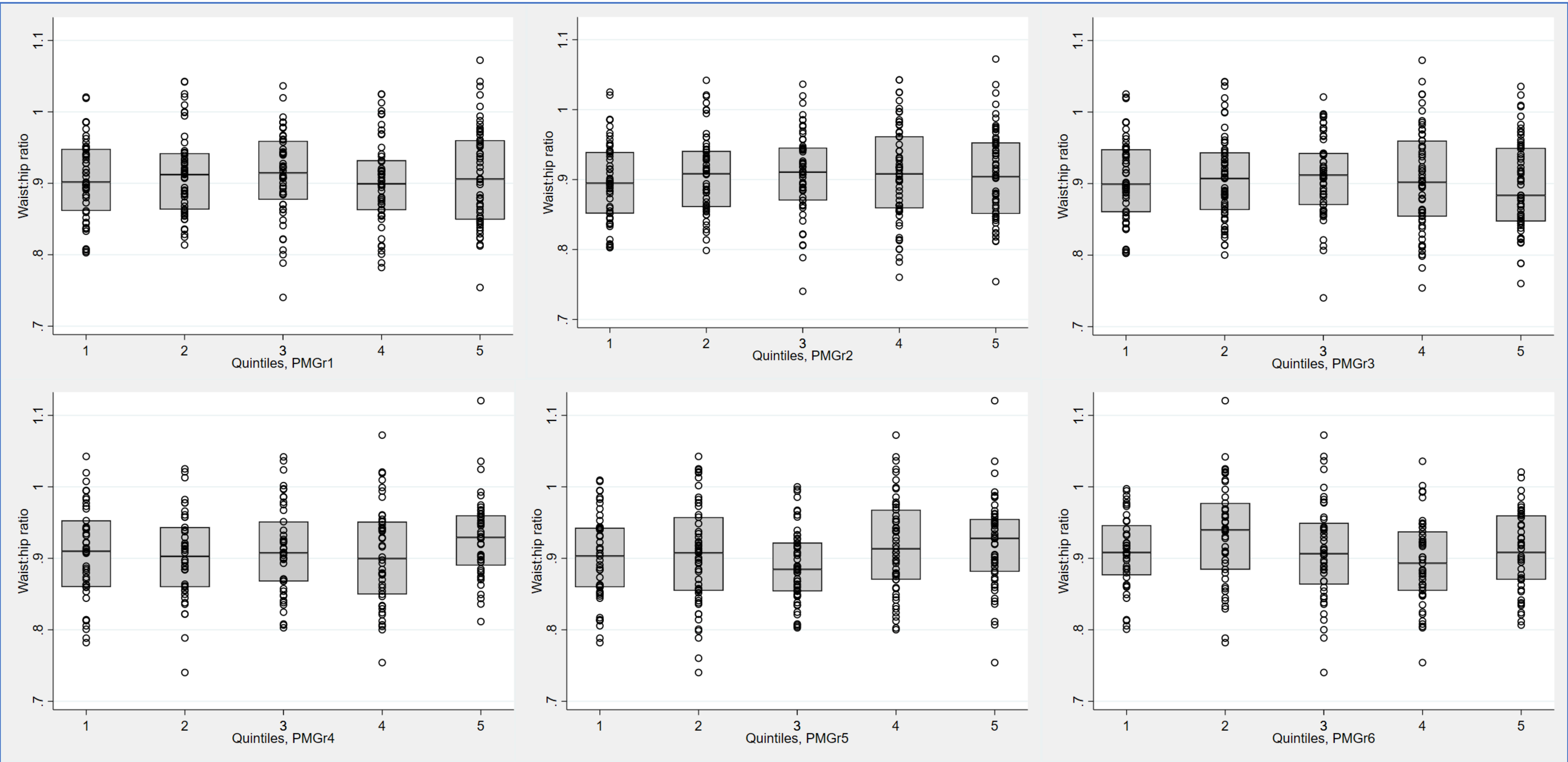
Waist:hip ratio
.7
.8
.9
1
1.1
1
2
3
4
5
Quintiles, PMGr1
Quintiles, PMGr2
Quintiles, PMGr3
Quintiles, PMGr4
Quintiles, PMGr5
Quintiles, PMGr6

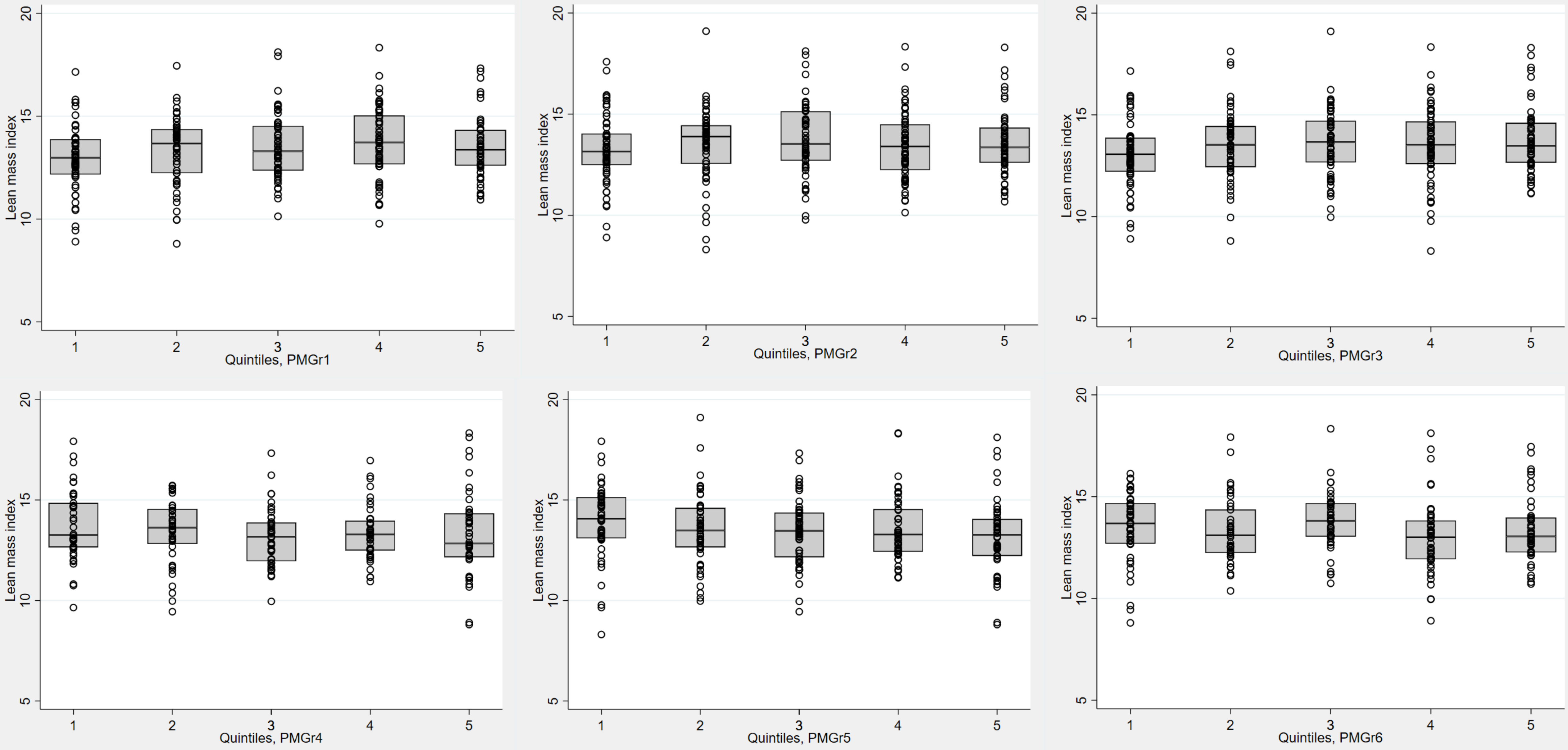
Lean mass index
Quintiles, PMGr1
Quintiles, PMGr2
Quintiles, PMGr3
Quintiles, PMGr4
Quintiles, PMGr5
Quintiles, PMGr6

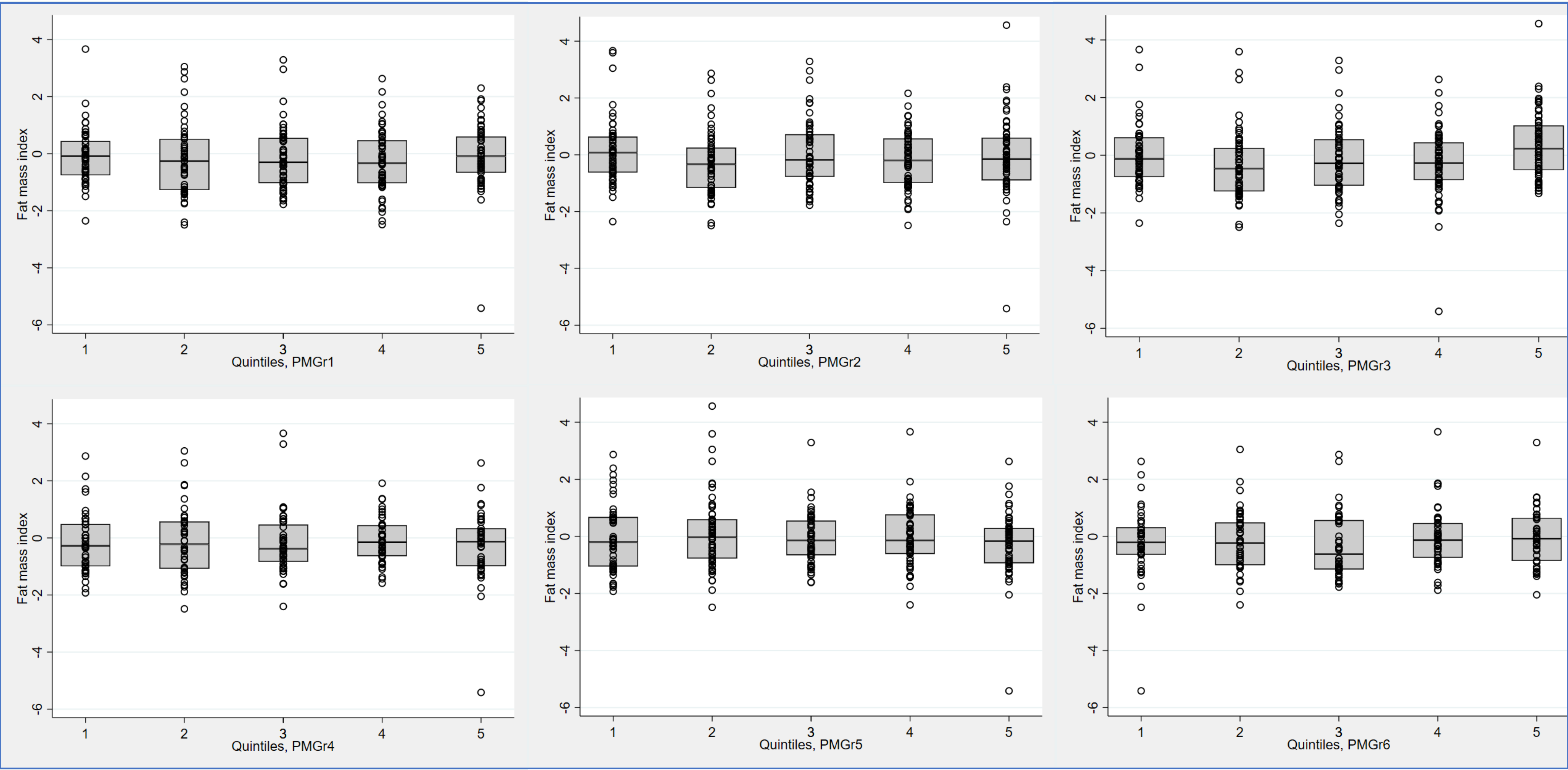
Fat mass index
Quintiles, PMGr1
Quintiles, PMGr2
Quintiles, PMGr3
Quintiles, PMGr4
Quintiles, PMGr5
Quintiles, PMGr6

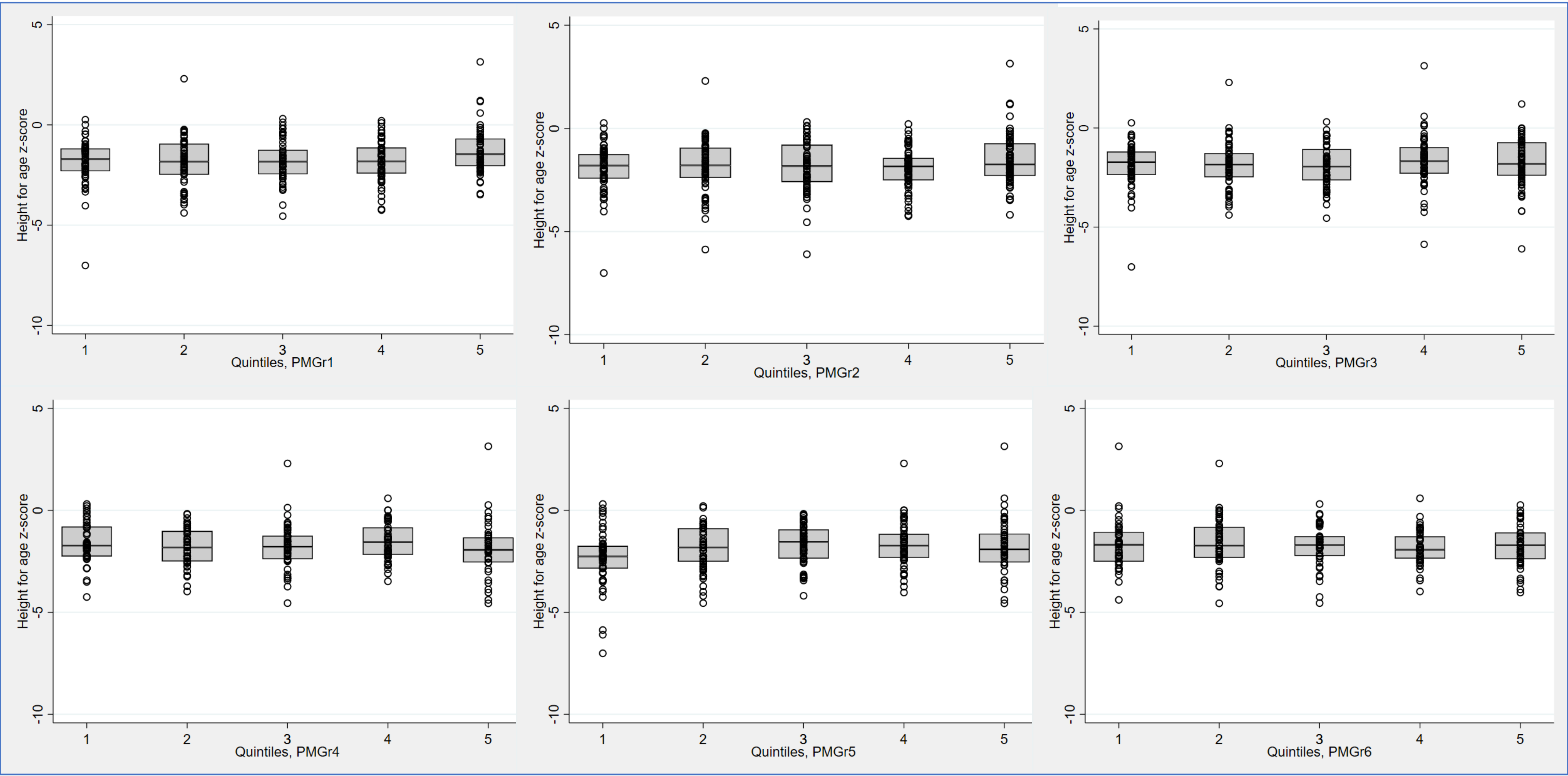
Height for age z-score
Quintiles, PMGr1
Quintiles, PMGr2
Quintiles, PMGr3
Quintiles, PMGr4
Quintiles, PMGr5
Quintiles, PMGr6

### Annex Table 3: Association between LCA classes and NCD outcomes

| NCD indicator | PMGr definition | Unadjusted difference | P-value | 95% CI | Adjusted difference | P-value | 95% CI |
|---|---|---|---|---|---|---|---|
| Systolic BP | WAZ | 0.6414 | 0.140 | (-0.21, 1.49) | -0.2037 | 0.645 | (-1.07, 0.67) |
| | Weight (kg) | 2.3708 | <0.001 | (1.38, 3.36) | -0.3867 | 0.593 | (-1.81, 1.04) |
| | HAZ | 1.1814 | 0.049 | (0.01, 2.36) | 0.0577 | 0.930 | (-1.24, 1.36) |
| Diastolic BP | WAZ | 0.2809 | 0.478 | (-0.49, 1.06) | 0.3101 | 0.478 | (-0.55, 1.17) |
| | Weight (kg) | 0.9327 | 0.049 | (0.00, 1.86) | 0.5185 | 0.469 | (-0.89, 1.92) |
| | HAZ | -0.3271 | 0.548 | (-1.39, 0.74) | -0.4877 | 0.453 | (-1.77, 0.79) |
| Hand grip | WAZ | 0.5679 | 0.002 | (0.21, 0.82) | 0.5780 | <0.001 | (0.27, 0.89) |
| | Weight (kg) | 1.9069 | <0.001 | (1.54, 2.28) | 0.9918 | <0.001 | (0.17, 1.51) |
| | HAZ | 0.6217 | 0.014 | (0.13, 1.12) | 0.9560 | <0.001 | (0.53, 1.38) |
| Waist circumference | WAZ | 0.8542 | <0.001 | (0.49, 1.21) | 0.9525 | <0.001 | (0.63, 1.27) |
| | Weight (kg) | 2.0531 | 0.000 | (1.66, 2.44) | 1.1631 | <0.001 | (0.62, 1.71) |
| | HAZ | 0.7906 | 0.003 | (0.28, 1.31) | 1.2171 | <0.001 | (0.78, 1.66) |

| NCD indicator | PMGr definition | Unadjusted difference | P-value | 95% CI | Adjusted difference | P-value | 95% CI |
|---|---|---|---|---|---|---|---|
| Waist:Hip ratio | WAZ | -0.0105 | <0.001 | (-0.02, -0.01) | -0.0094 | 0.001 | (-0.01, -0.00) |
| | Weight (kg) | -0.0152 | <0.001 | (-0.02, -0.01) | -0.0135 | 0.004 | (-0.02, -0.00) |
| | HAZ | -0.0136 | <0.001 | (-0.02, -0.01) | -0.0150 | <0.001 | (-0.02, -0.01) |
| Lean mass index (BIA) | WAZ | 0.2927 | <0.001 | (0.14, 0.44) | 0.2889 | <0.001 | (0.13, 0.44) |
| | Weight (kg) | 0.4648 | <0.001 | (0.29, 0.64) | 0.4182 | 0.002 | (0.16, 0.68) |
| | HAZ | 0.1614 | 0.139 | (-0.05, 0.38) | 0.1949 | 0.076 | (-0.02, 0.41) |
| Fat mass index (BIA) | WAZ | 0.1300 | 0.014 | (0.03, 0.23) | 0.1667 | 0.002 | (0.06, 0.27) |
| | Weight (kg) | 0.2919 | <0.001 | (0.17, 0.41) | 0.1933 | 0.032 | (0.02, 0.37) |
| | HAZ | 0.0369 | 0.621 | (-0.11, 0.18) | 0.1105 | 0.135 | (-0.03, 0.26) |
| HAZ | WAZ | 0.3202 | <0.001 | (0.22, 0.42) | 0.3144 | <0.001 | (0.21, 0.41) |
| | Weight (kg) | -0.0746 | 0.263 | (-0.21, 0.06) | 0.3719 | <0.001 | (0.19, 0.54) |
| | HAZ | 0.6768 | <0.001 | (0.55, 0.81) | 0.6358 | <0.001 | (0.51, 0.76) |

### Annex Figures 5a-2h

Boxplots showing associations between LCA classes for weight, WAZ and HAZ, and NCD risk outcomes

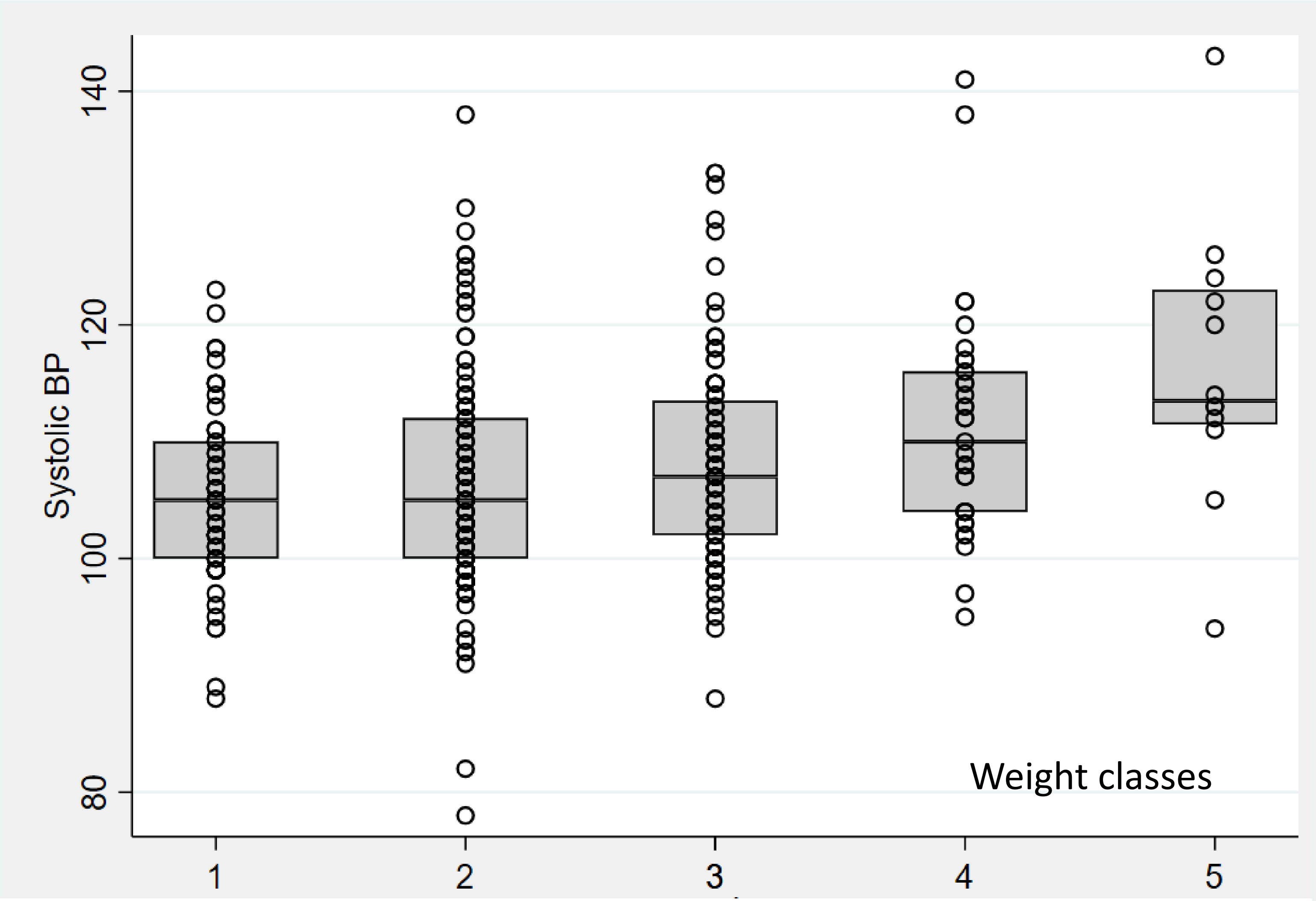

Systolic BP
140
120
100
80
1
2
3
4
5
Weight classes


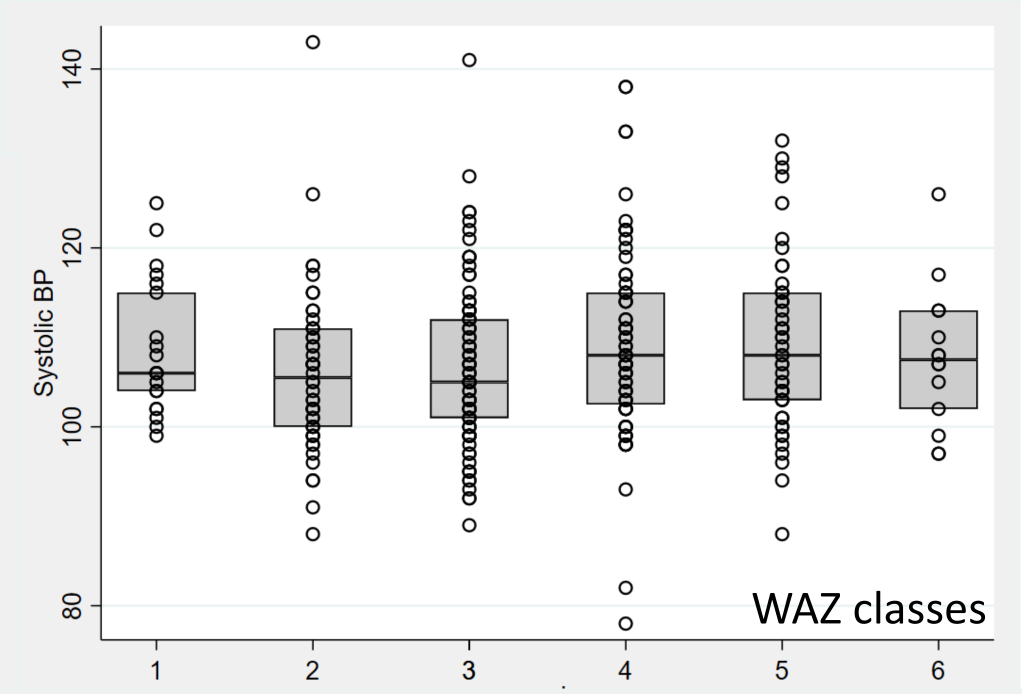

Systolic BP
140
120
100
80
1
2
3
4
5
6
WAZ classes


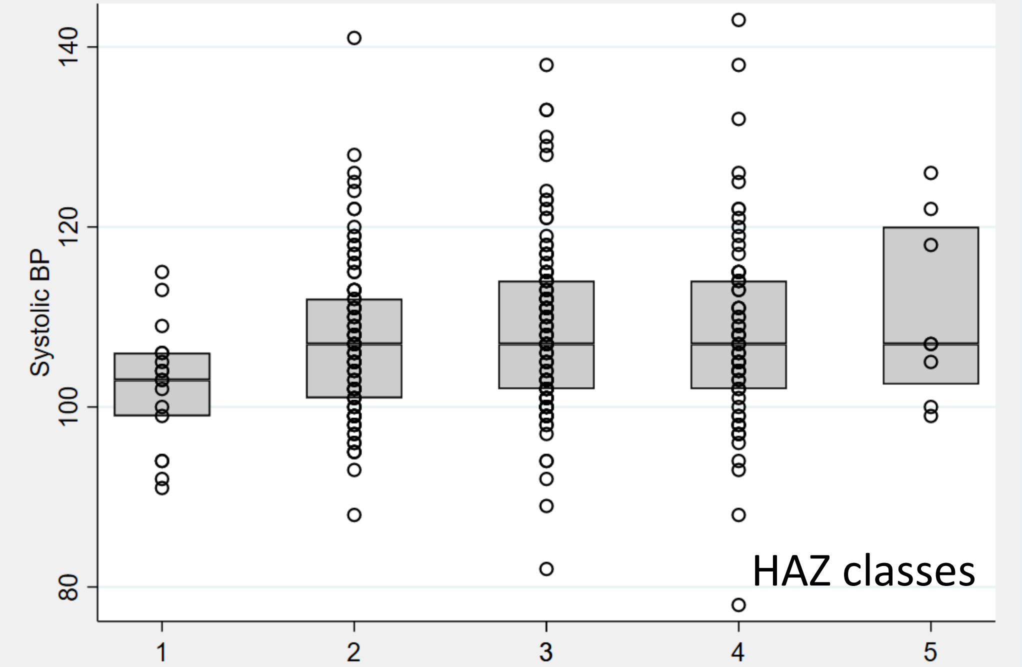

Systolic BP
140
120
100
80
1
2
3
4
5
HAZ classes

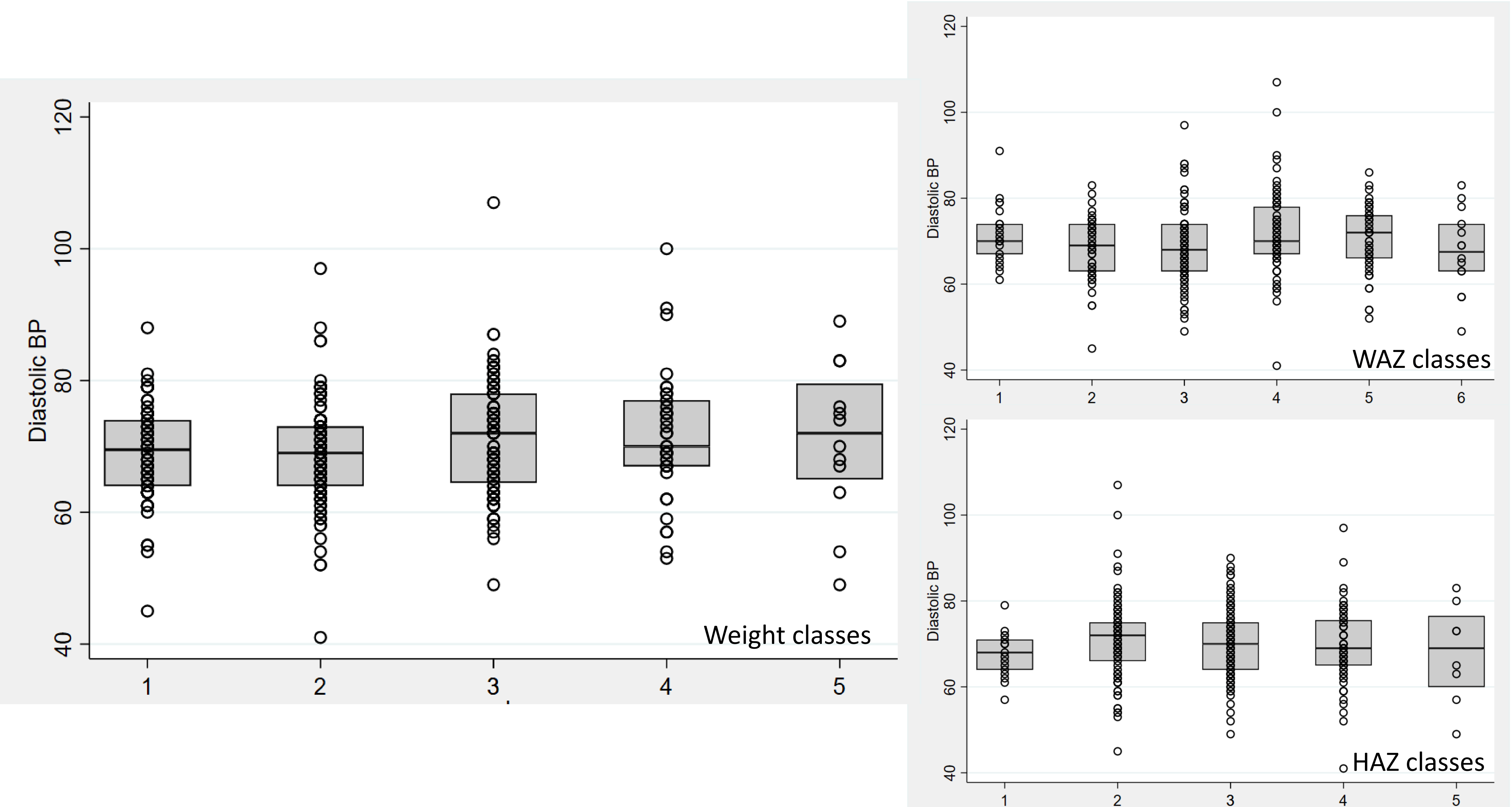

Diastolic BP
Weight classes
WAZ classes
HAZ classes

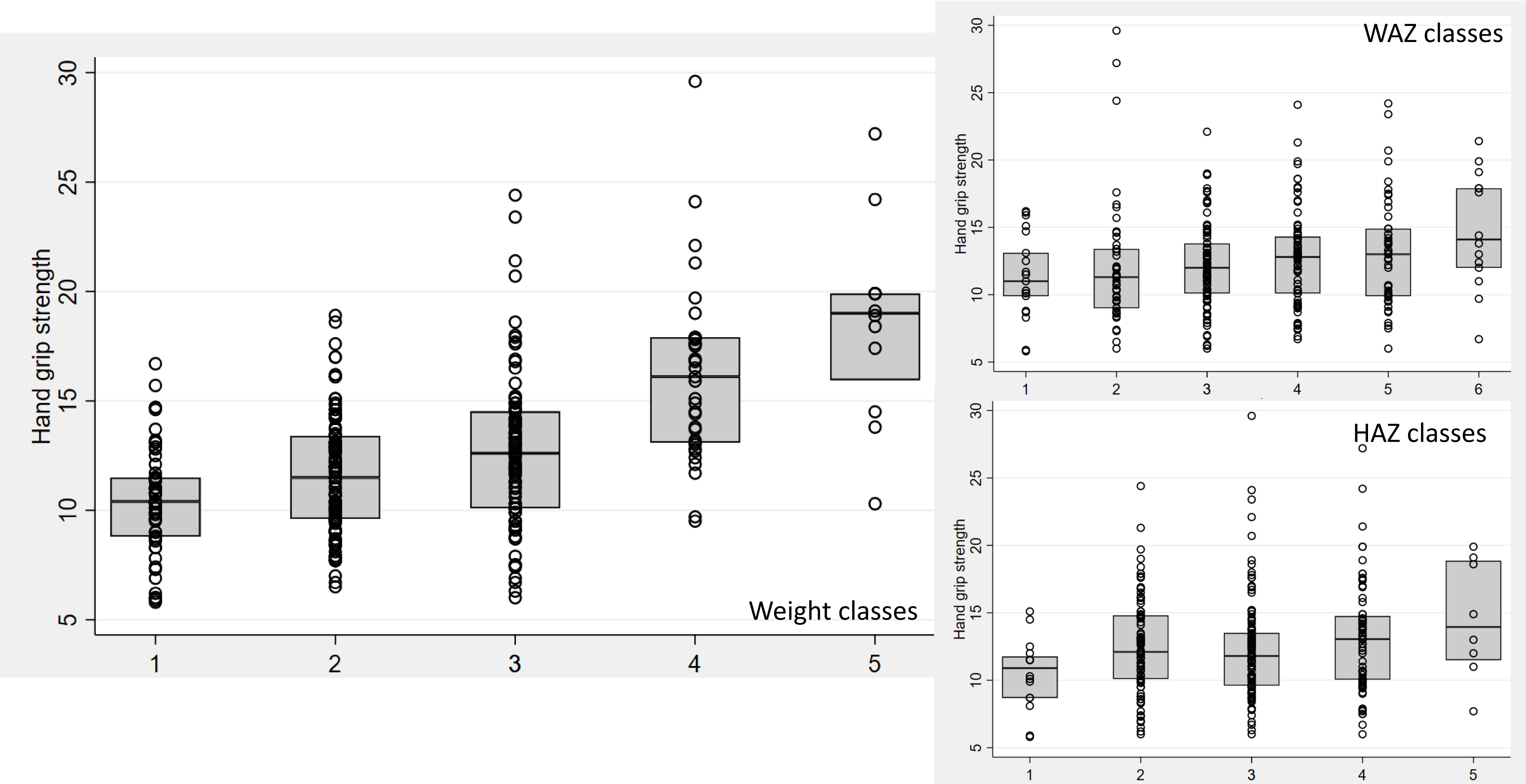
Hand grip strength
30
25
20
15
10
5
1
2
3
4
5
Weight classes
WAZ classes
6
HAZ classes

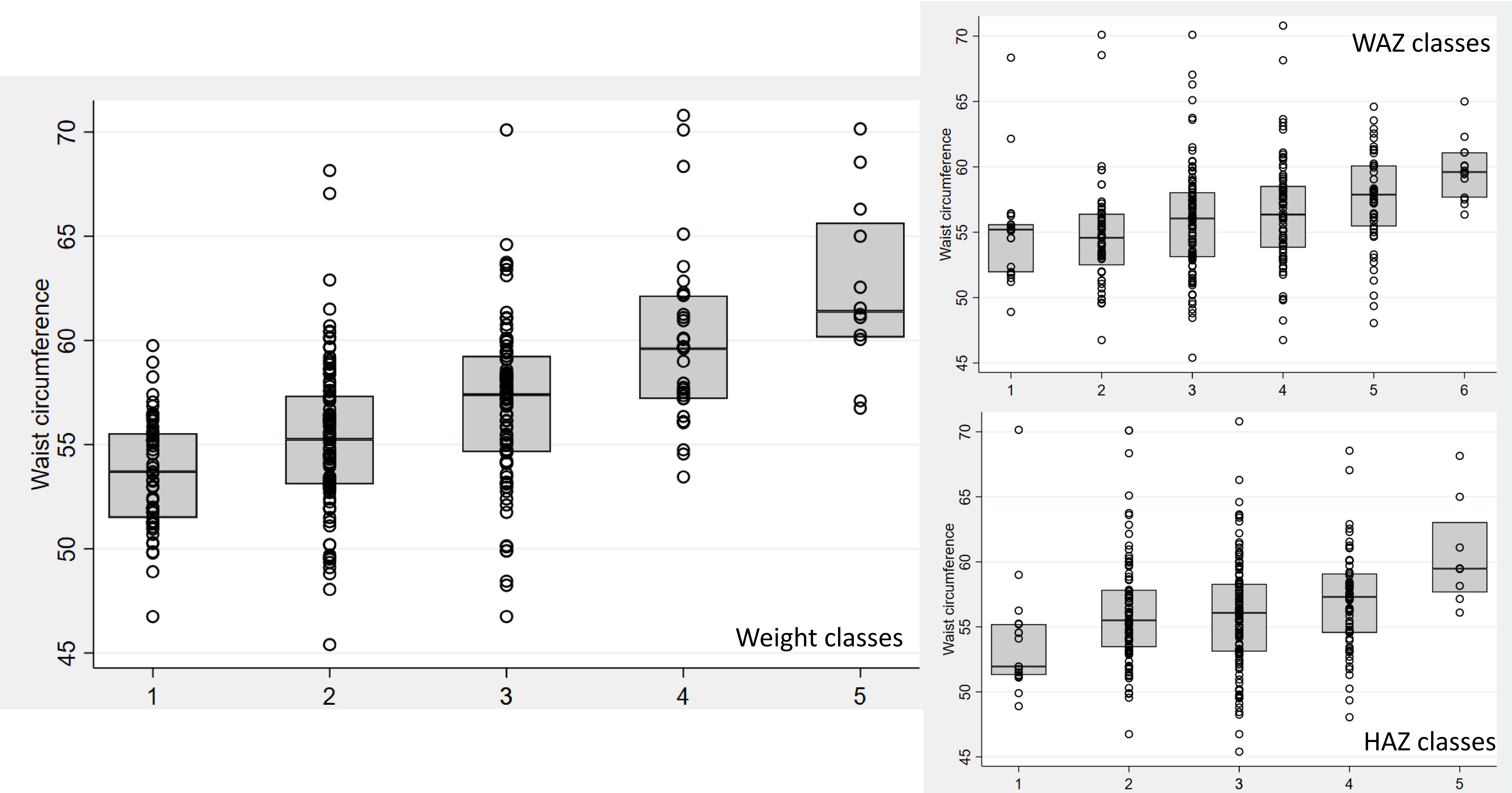
Waist circumference
Weight classes
WAZ classes
HAZ classes

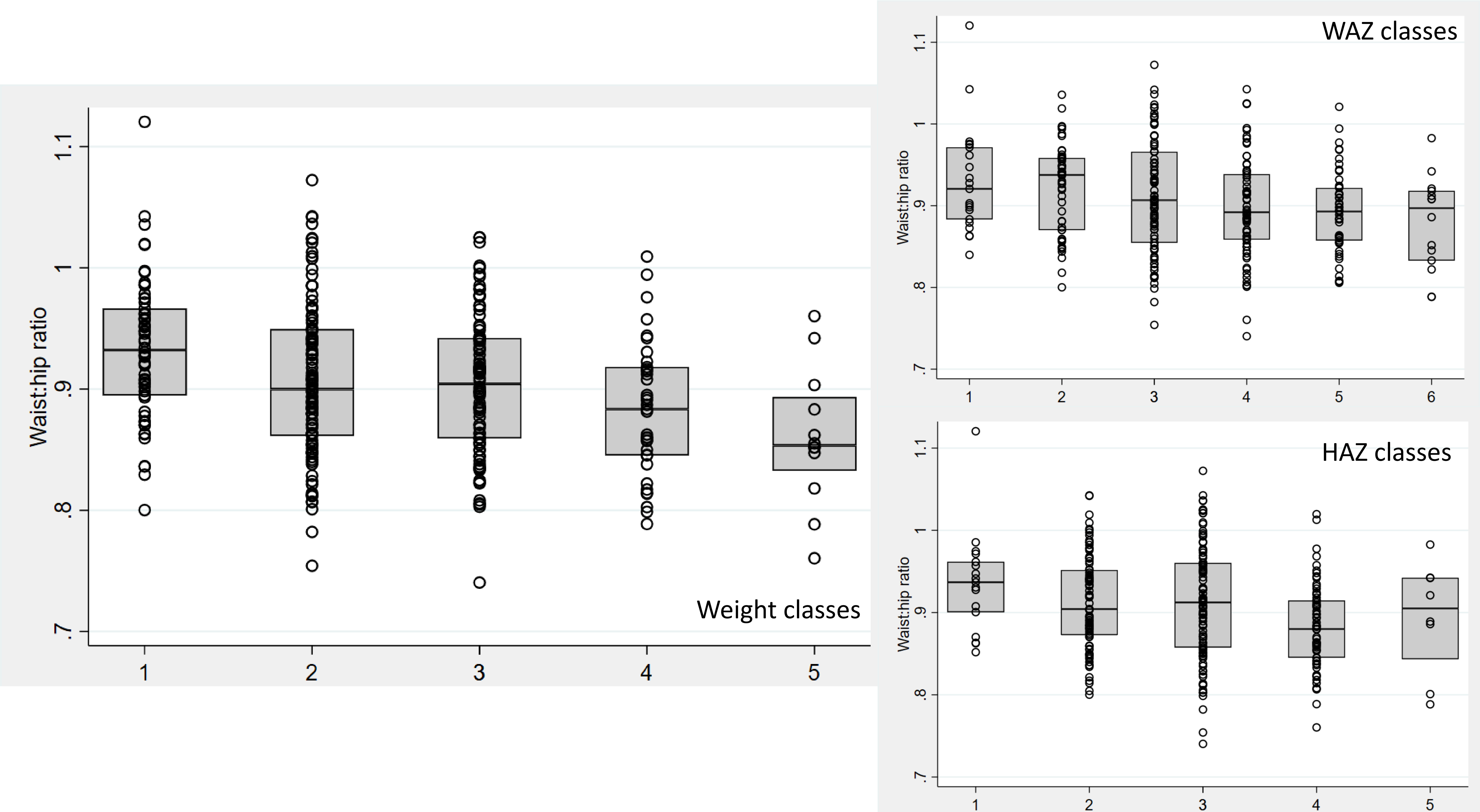
Waist:hip ratio
1.1
1
.9
.8
.7
1
2
3
4
5
Weight classes
WAZ classes
6
HAZ classes

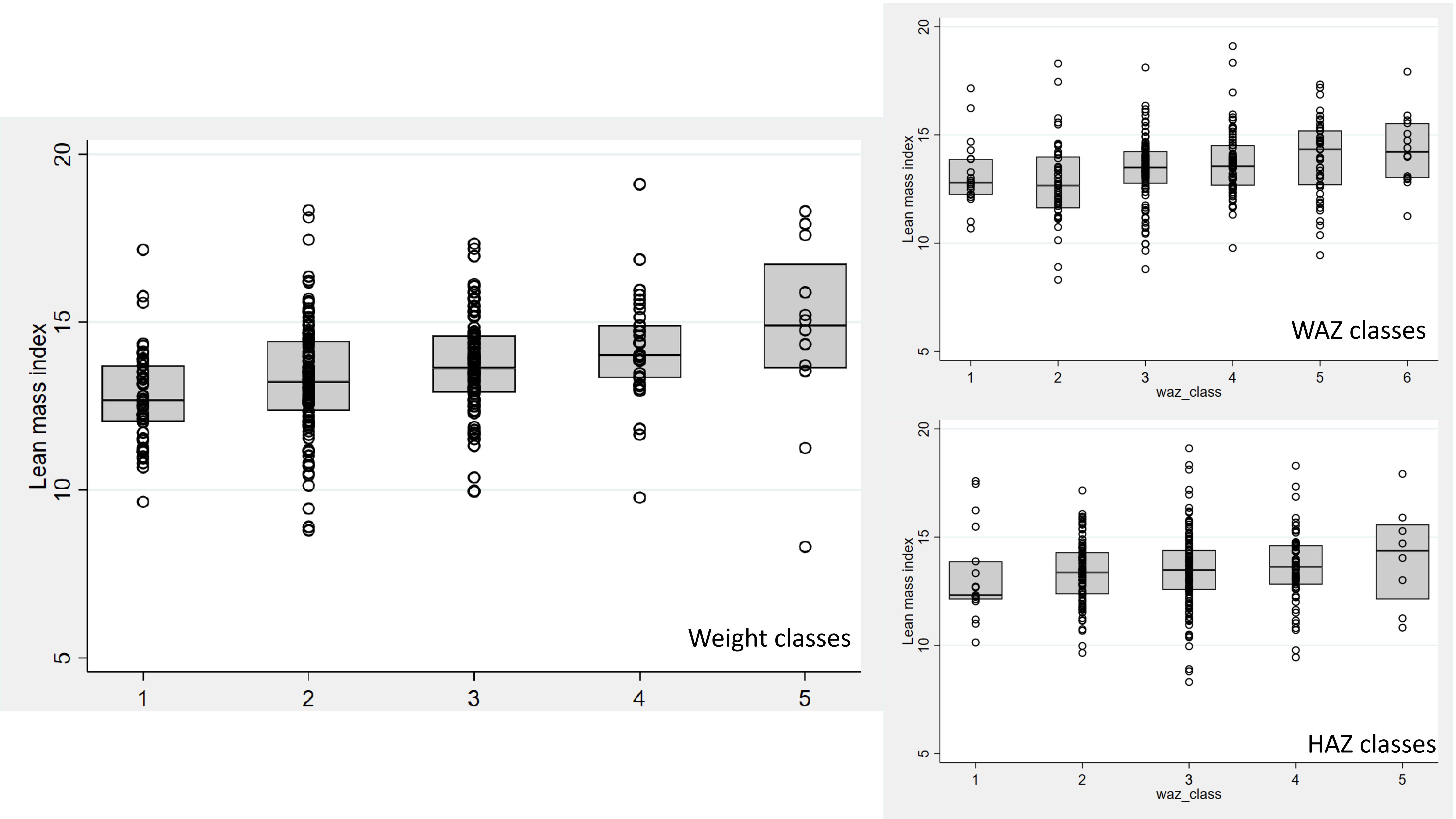

Lean mass index
20
15
10
5
1
2
3
4
5
Weight classes
Lean mass index
20
15
10
5
1
2
3
4
5
6
waz_class
WAZ classes
Lean mass index
20
15
10
5
1
2
3
4
5
waz_class
HAZ classes

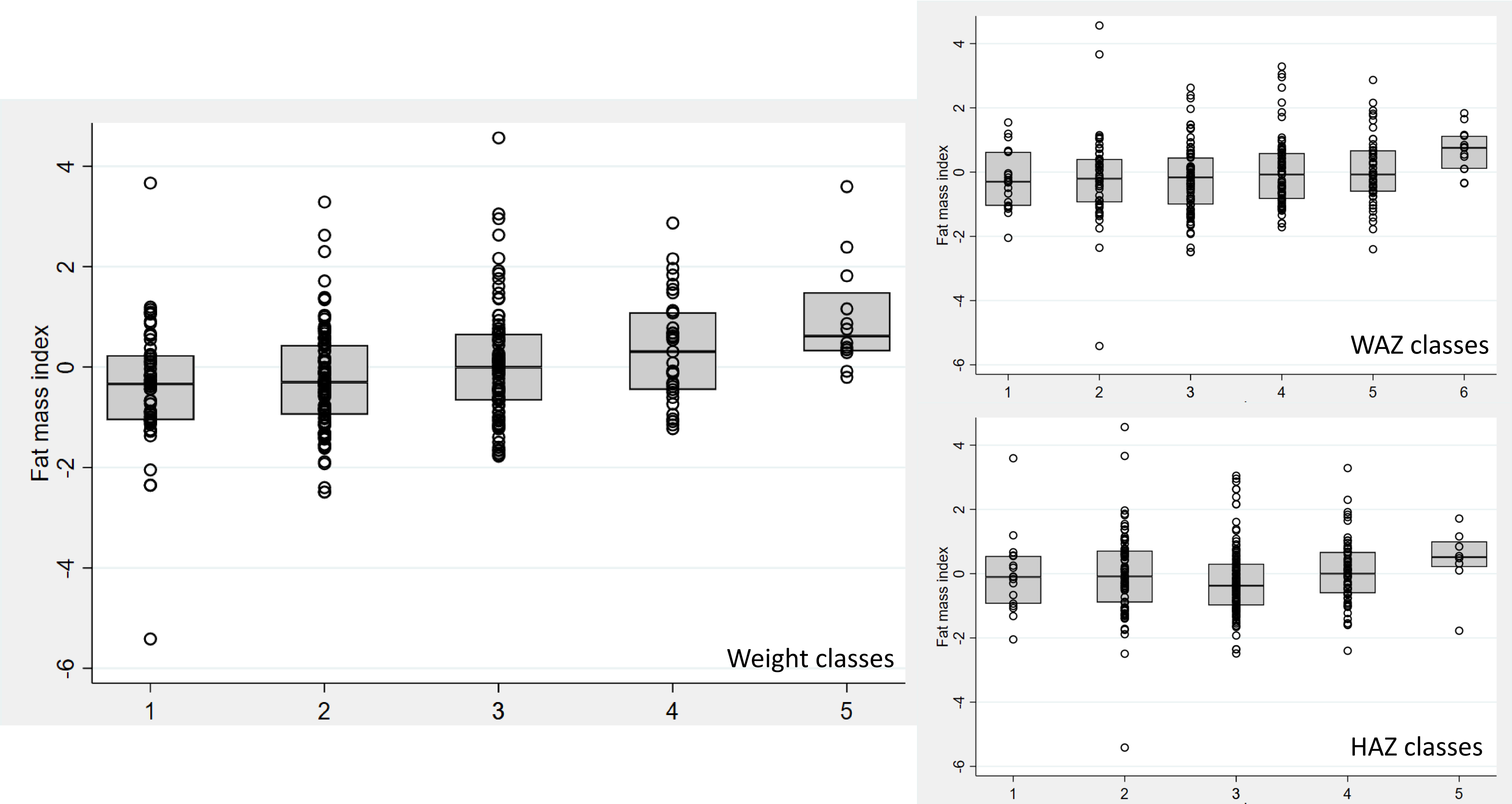
Fat mass index
Weight classes
WAZ classes
HAZ classes
4
2
0
-2
-4
-6
1
2
3
4
5
6

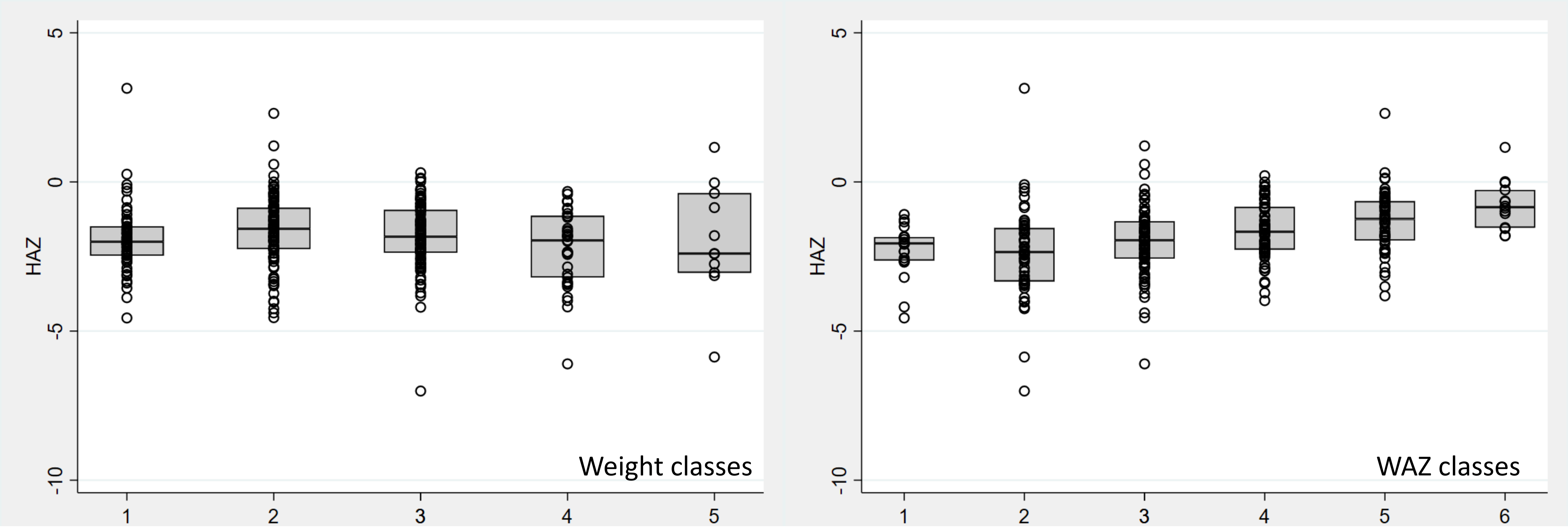

5
0
-5
-10
HAZ
1
2
3
4
5
Weight classes
5
0
-5
-10
HAZ
1
2
3
4
5
6
WAZ classes

### Annex Table 4: Association between Admission anthropometry and NCD outcomes

| NCD indicator | PMGr definition | Unadjusted difference | P-value | 95% CI | Adjusted difference | P-value | 95% CI |
|---|---|---|---|---|---|---|---|
| Systolic BP | Admission WAZ | **0.82** | **0.016** | **(0.15, 1.50)** | 0.01 | 0.986 | (-0.73, 0.74) |
| | Admission weight (kg) | **0.88** | **<0.001** | **(0.50, 1.27)** | -0.14 | 0.624 | (-0.72, 0.43) |
| | Admission HAZ | 0.68 | 0.061 | (-0.03, 1.40) | -0.19 | 0.657 | (-1.02, 0.64) |
| Diastolic BP | Admission WAZ | 0.52 | 0.114 | (-0.13, 1.17) | 0.55 | 0.140 | (-0.18, 1.28) |
| | Admission weight (kg) | 0.25 | 0.159 | (-0.10, 0.61) | 0.07 | 0.797 | (-0.49, 0.65) |
| | Admission HAZ | -0.15 | 0.661 | (-0.84, 0.54) | -0.12 | 0.775 | (-0.95, 0.71) |
| Hand grip | Admission WAZ | **0.63** | **<0.001** | (0.37, 0.89) | **0.49** | **<0.000** | **(0.24, 0.74)** |
| | Admission weight (kg) | **0.71** | **<0.001** | **(0.57, 0.85)** | **0.31** | **0.005** | **(0.09, 0.53)** |
| | Admission HAZ | **0.42** | **0.004** | **(0.14, 0.70)** | **0.53** | **<0.001** | **(0.28, 0.79)** |
| Waist circumference | Admission WAZ | **0.89** | **<0.001** | **(0.62, 1.15)** | 0.87 | **<0.001** | (0.60, 1.13) |
| | Admission weight (kg) | **0.82** | **<0.001** | **(0.68, 0.97)** | **0.52** | **<0.001** | **(0.30, 0.75)** |
| | Admission HAZ | **0.68** | **<0.001** | **(0.39, 0.98)** | 0.85 | **<0.001** | **(0.57, 1.12)** |

| NCD indicator | PMGr definition | Unadjusted difference | P-value | 95% CI | Adjusted difference | P-value | 95% CI |
|---|---|---|---|---|---|---|---|
| Waist:Hip ratio | Admission WAZ | **-0.01** | **<0.001** | **(-0.01, -0.00)** | **-0.01** | **0.004** | **(-0.01, -0.00)** |
| | Admission weight (kg) | -0.01 | **<0.001** | **(-0.01, -0.00)** | **-0.01** | **0.008** | **(-0.01, -0.00)** |
| | Admission HAZ | **-0.01** | **0.001** | **(-0.01, -0.00)** | **-0.08** | **0.001** | (-0.01, -0.00) |
| Lean mass index (BIA) | Admission WAZ | **0.25** | **<0.001** | **(0.12, 0.37)** | **0.24** | **<0.001** | **(0.11, 0.38)** |
| | Admission weight (kg) | **0.18** | **<0.001** | **(0.11, 0.25)** | **0.18** | **0.001** | **(0.07, 0.29)** |
| | Admission HAZ | 0.09 | 0.206 | (-0.05, 0.22) | 0.09 | 0.210 | (-0.05, 0.23) |
| Fat mass index (BIA) | Admission WAZ | **0.13** | **0.003** | **(0.05, 0.21)** | **0.14** | **0.002** | **(0.06, 0.24)** |
| | Admission weight (kg) | **0.12** | **<0.001** | **(0.08, 0.17)** | **0.10** | **0.005** | **(0.03, 0.18)** |
| | Admission HAZ | 0.08 | 0.097 | (-0.01, 0.17) | **0.11** | **0.024** | **(0.01, 0.19)** |
| HAZ | Admission WAZ | **0.22** | **<0.001** | **(0.13, 0.29)** | **0.25** | **<0.001** | **(0.16, 0.33)** |
| | Admission weight (kg) | -0.03 | 0.313 | (-0.08, 0.03) | **0.15** | **<0.001** | **(0.08, 0.22** |
| | Admission HAZ | **0.40** | **<0.001** | **(0.32, 0.48)** | **0.39** | **<0.001** | **(0.31, 0.47)** |

### Annex Figure 6

Growth patterns for WAZ, weight and HAZ described based on LCA classes which have been standardised by admission anthropometry

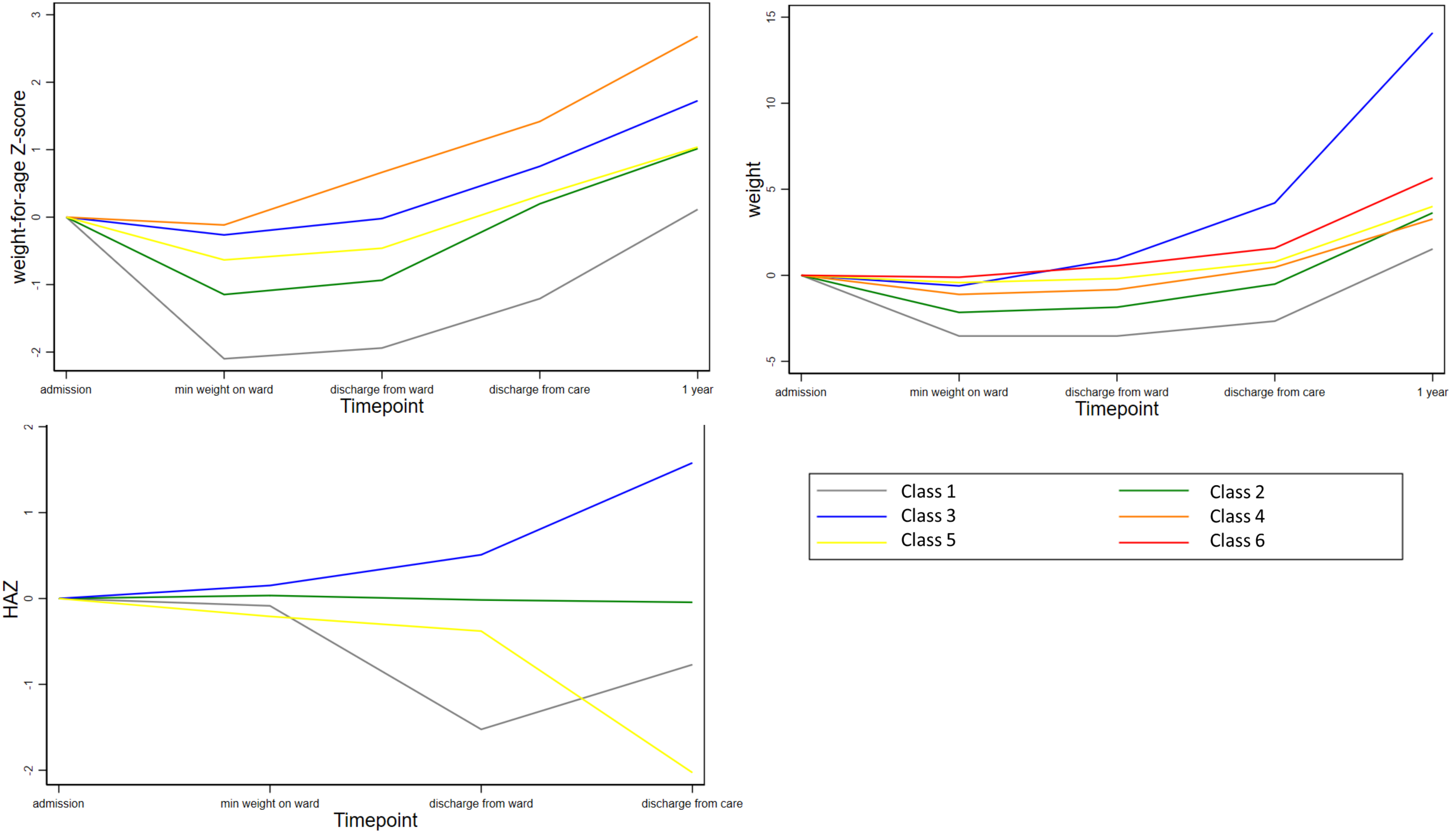

weight-for-age Z-score
3
2
1
0
-1
-2
admission
min weight on ward
discharge from ward
discharge from care
1 year
Timepoint
weight
15
10
5
0
-5
admission
min weight on ward
discharge from ward
discharge from care
1 year
Timepoint
HAZ
2
1
0
-1
-2
admission
min weight on ward
discharge from ward
discharge from care
Timepoint
Class 1
Class 2
Class 3
Class 4
Class 5
Class 6

### Annex Figures 7a-5g

Boxplots showing associations between LCA classes for weight, WAZ and HAZ (standardised for admission anthropometry), and NCD risk outcomes

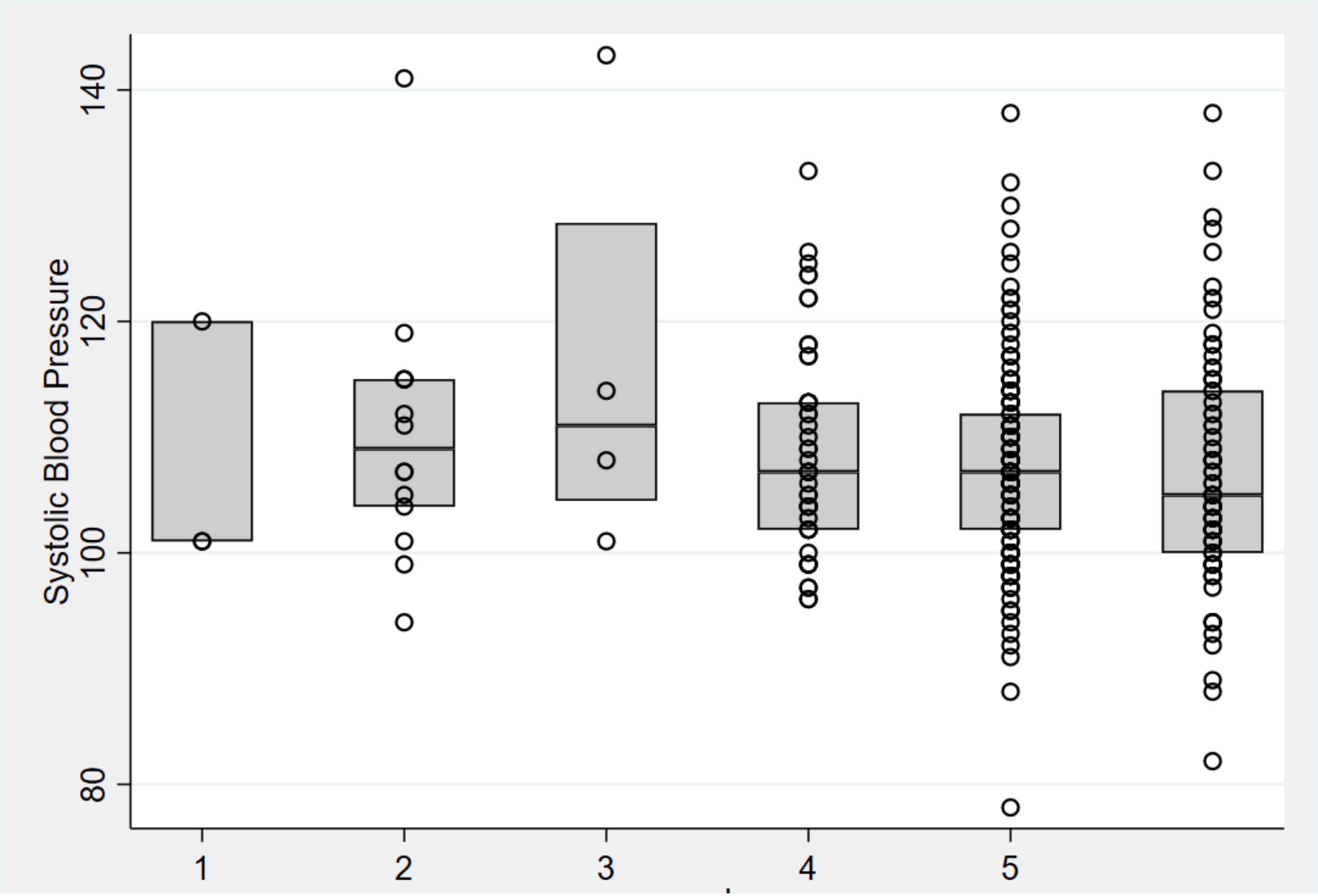
Systolic Blood Pressure
140
120
100
80
1
2
3
4
5

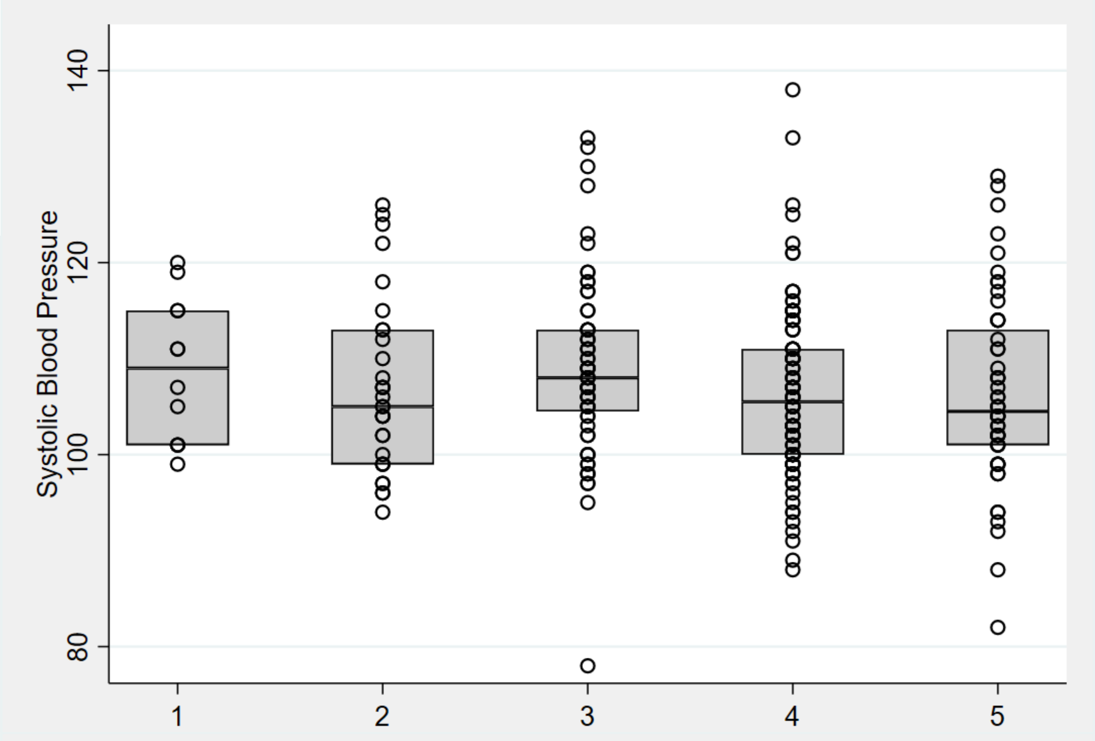
Systolic Blood Pressure
140
120
100
80
1
2
3
4
5

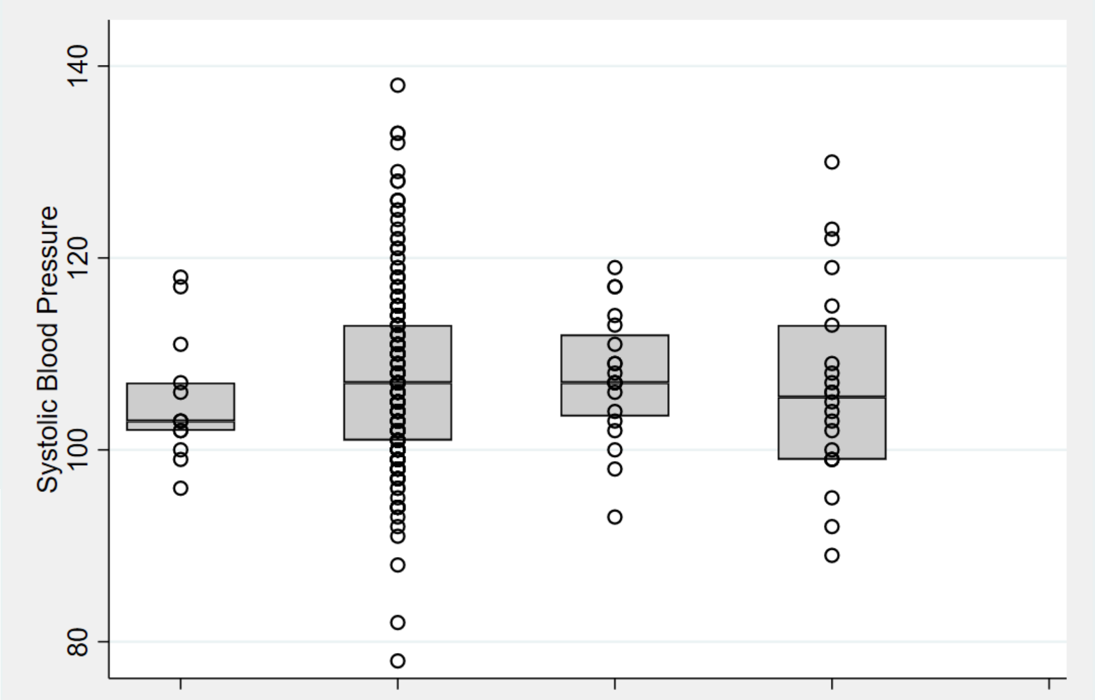
Systolic Blood Pressure
140
120
100
80

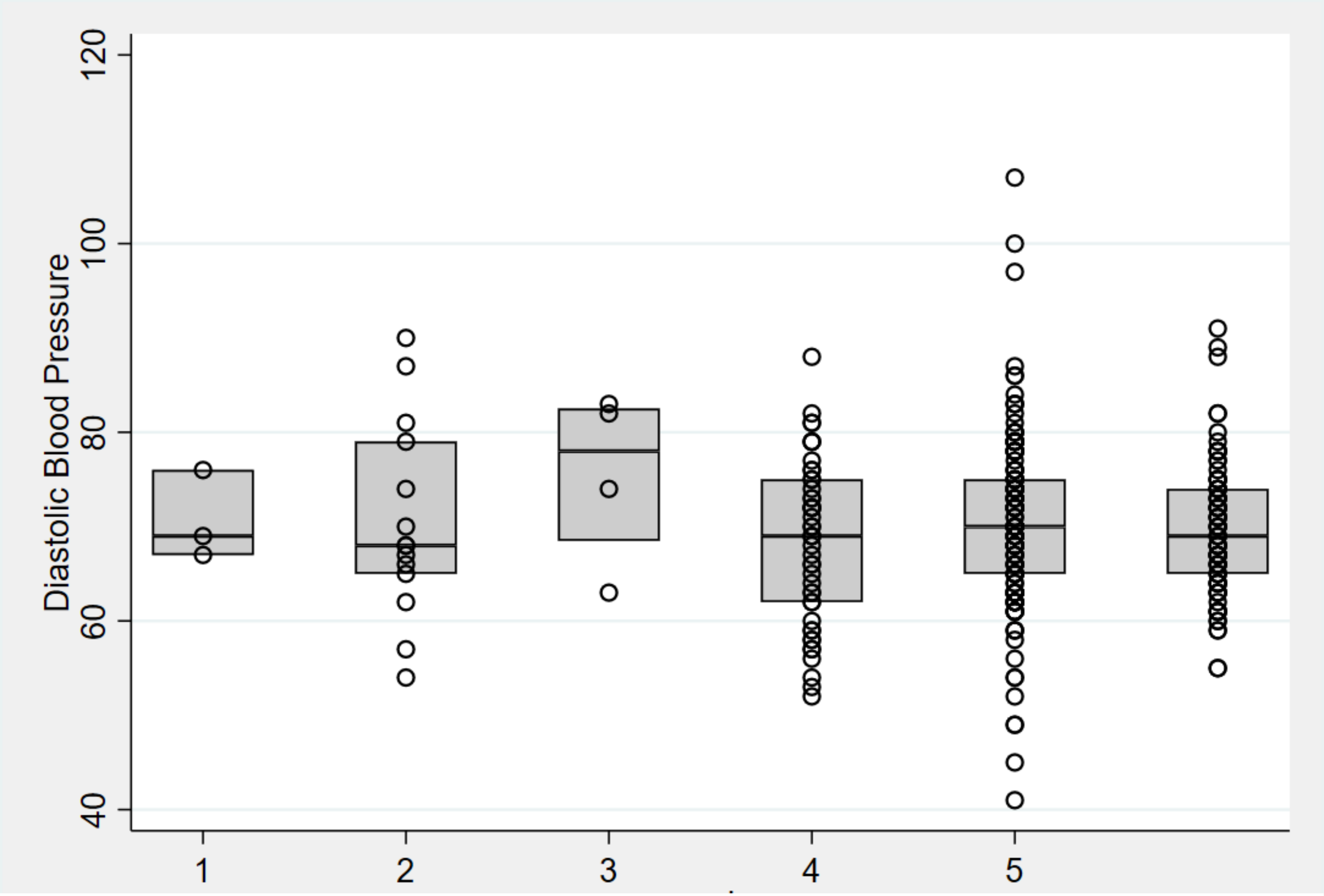
Diastolic Blood Pressure
40
60
80
100
120
1
2
3
4
5

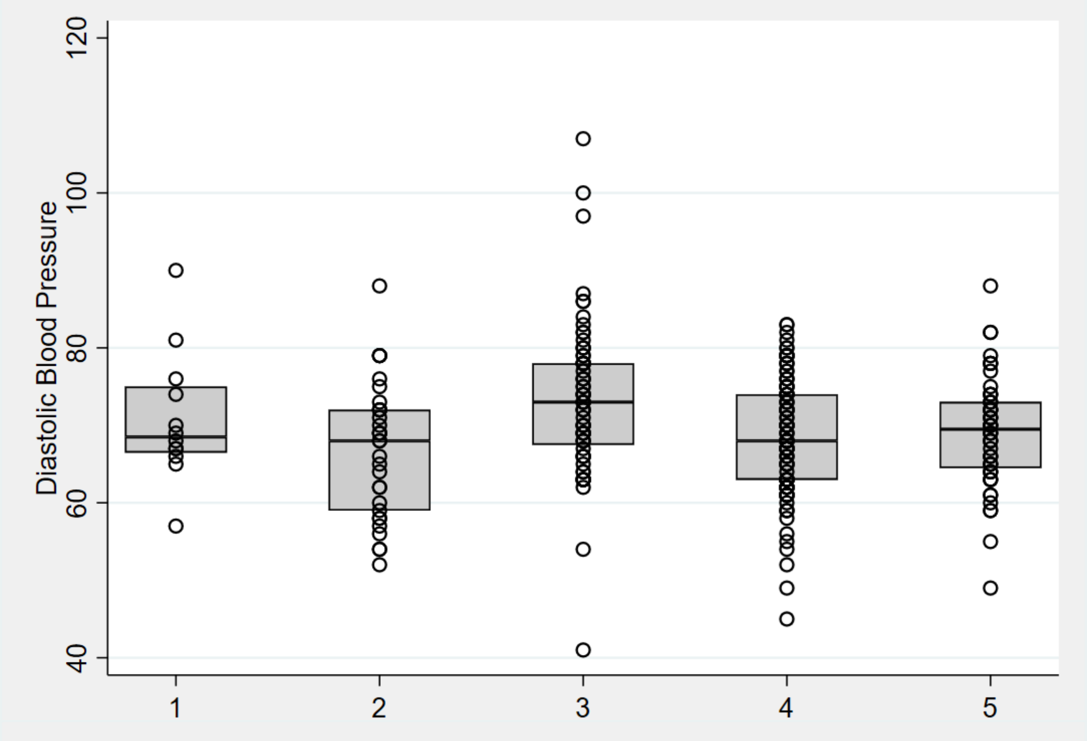
Diastolic Blood Pressure
40
60
80
100
120
1
2
3
4
5

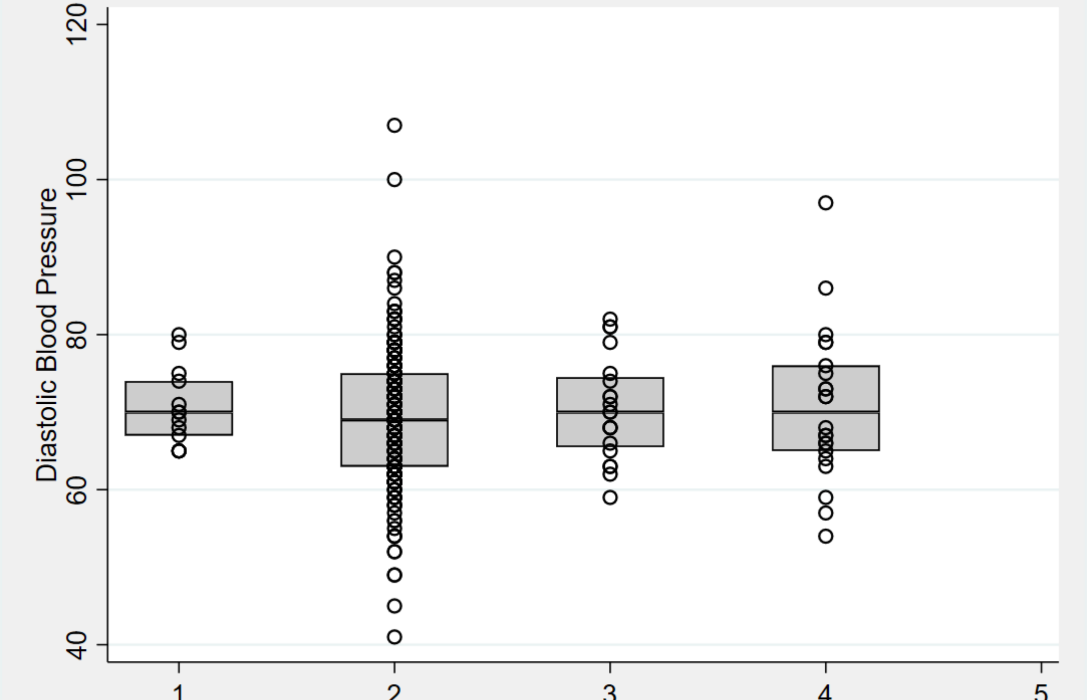
Diastolic Blood Pressure
40
60
80
100
120
1
2
3
4
5

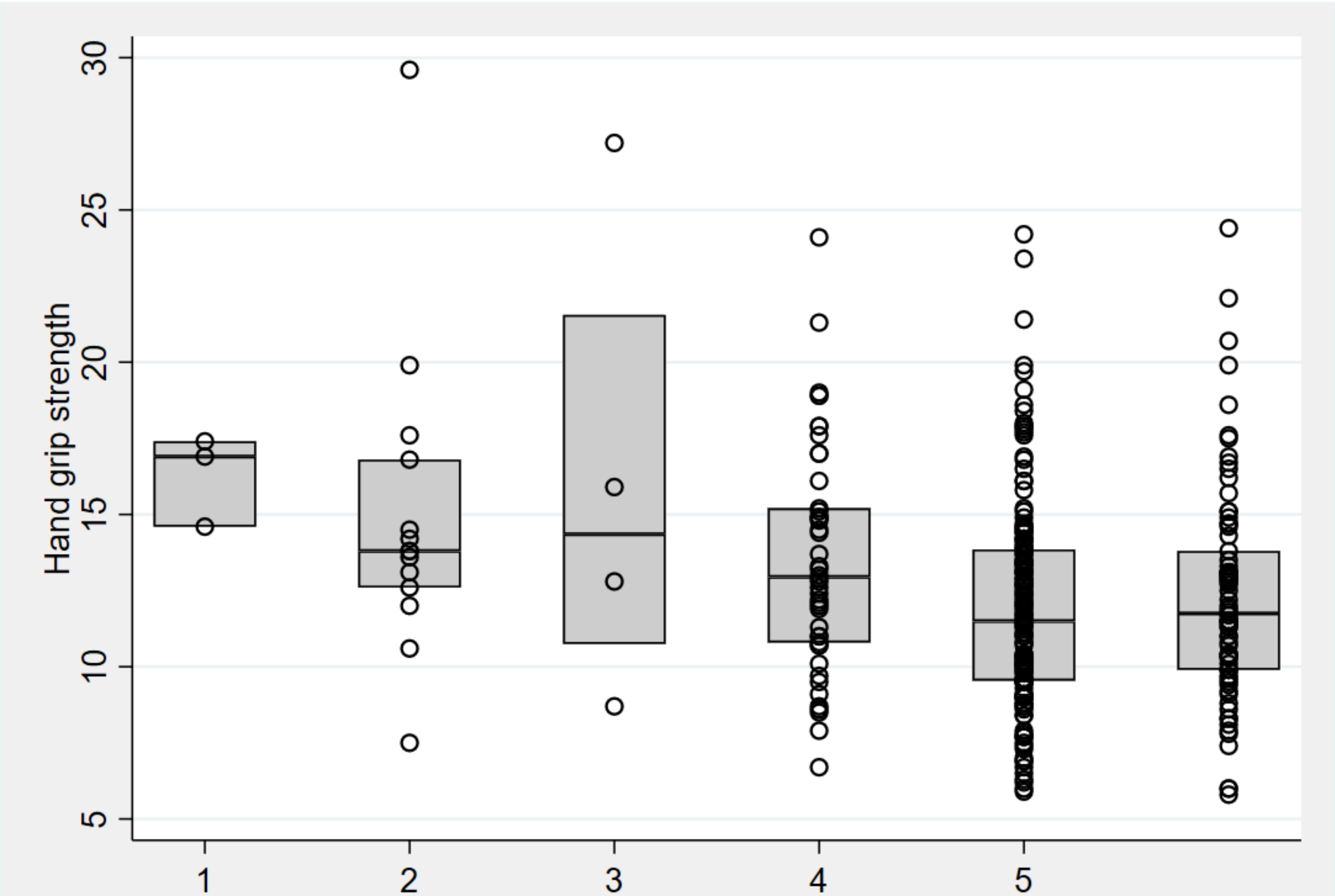
Hand grip strength
5
10
15
20
25
30
1
2
3
4
5

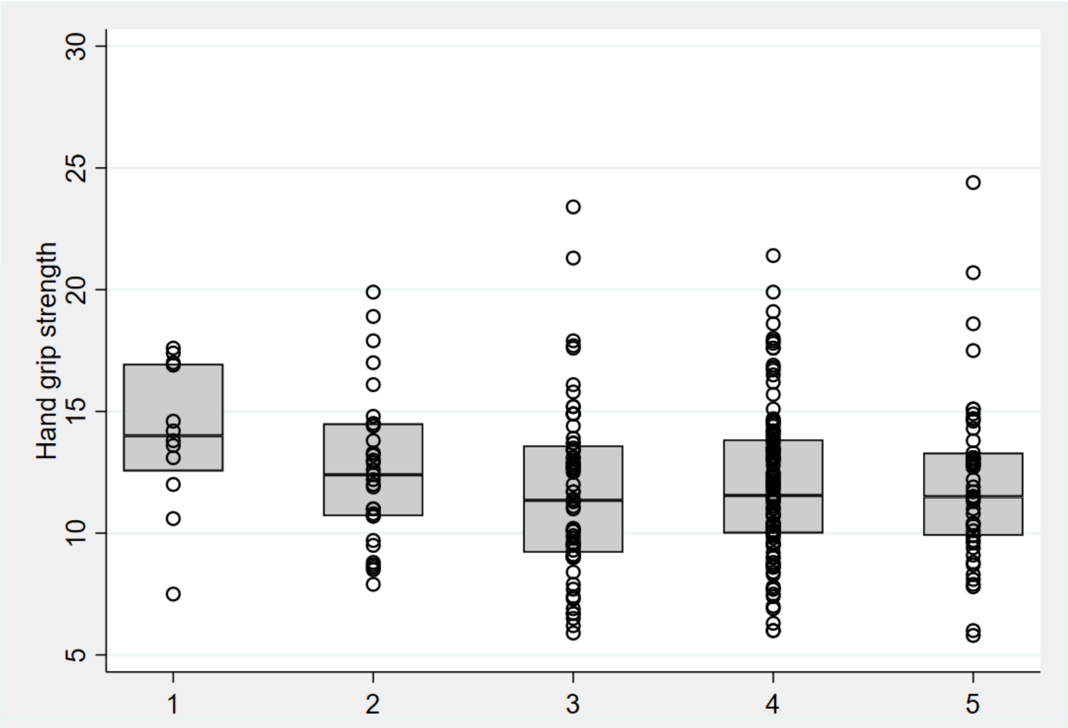
Hand grip strength
5
10
15
20
25
30
1
2
3
4
5

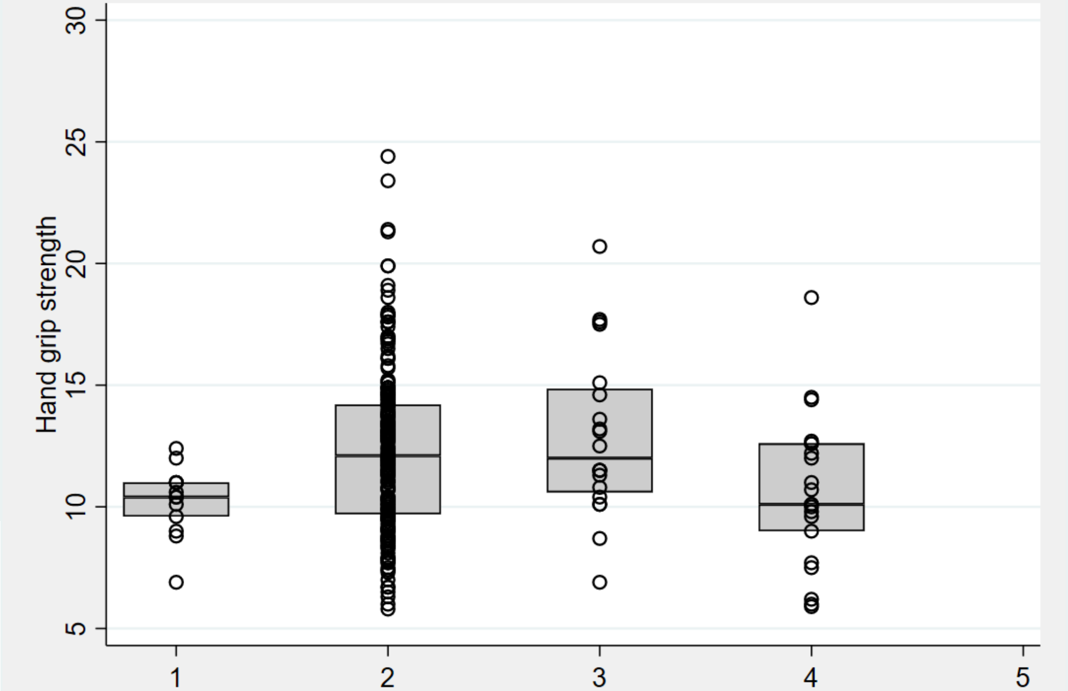
Hand grip strength
5
10
15
20
25
30
1
2
3
4
5

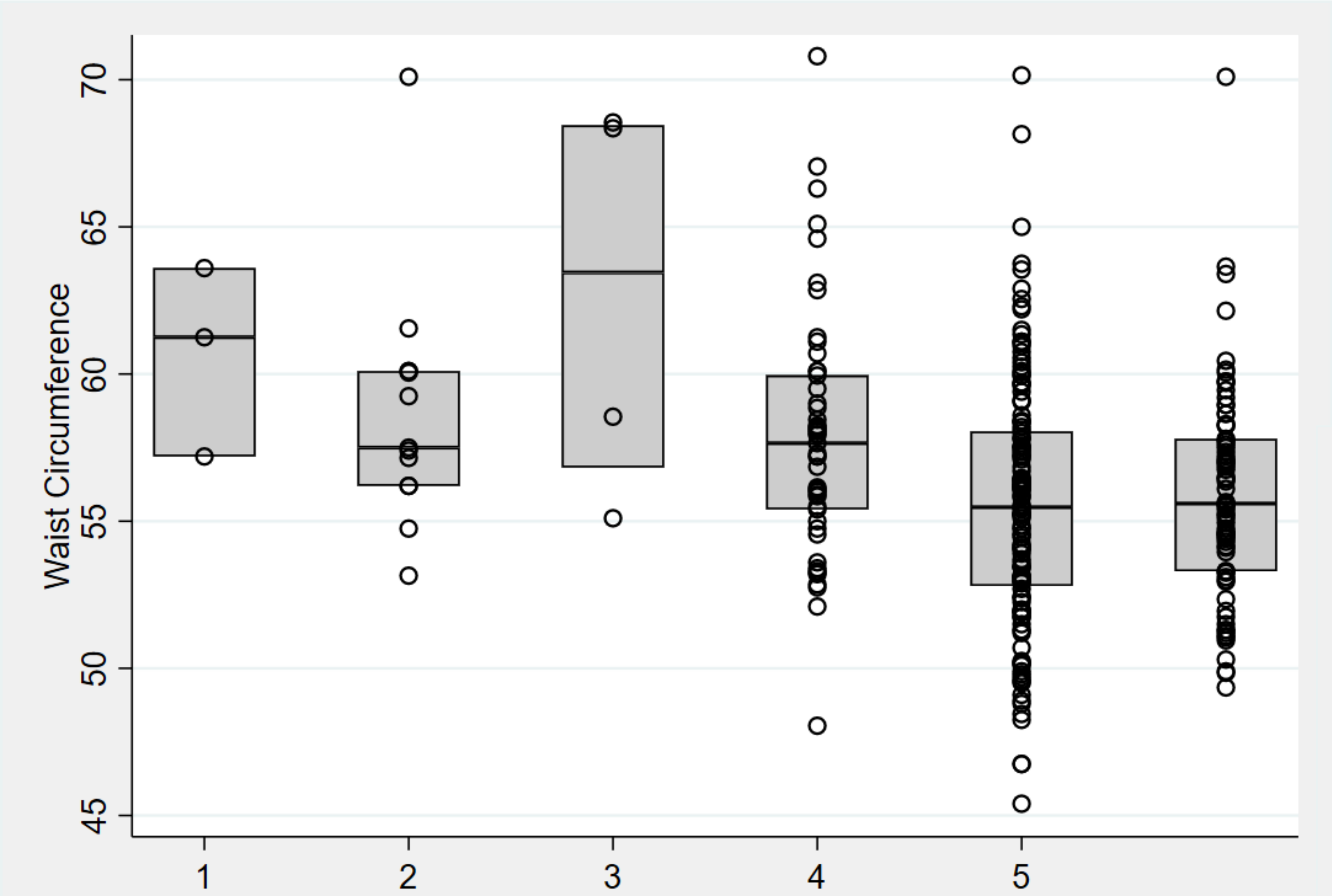
Waist Circumference
45
50
55
60
65
70
1
2
3
4
5

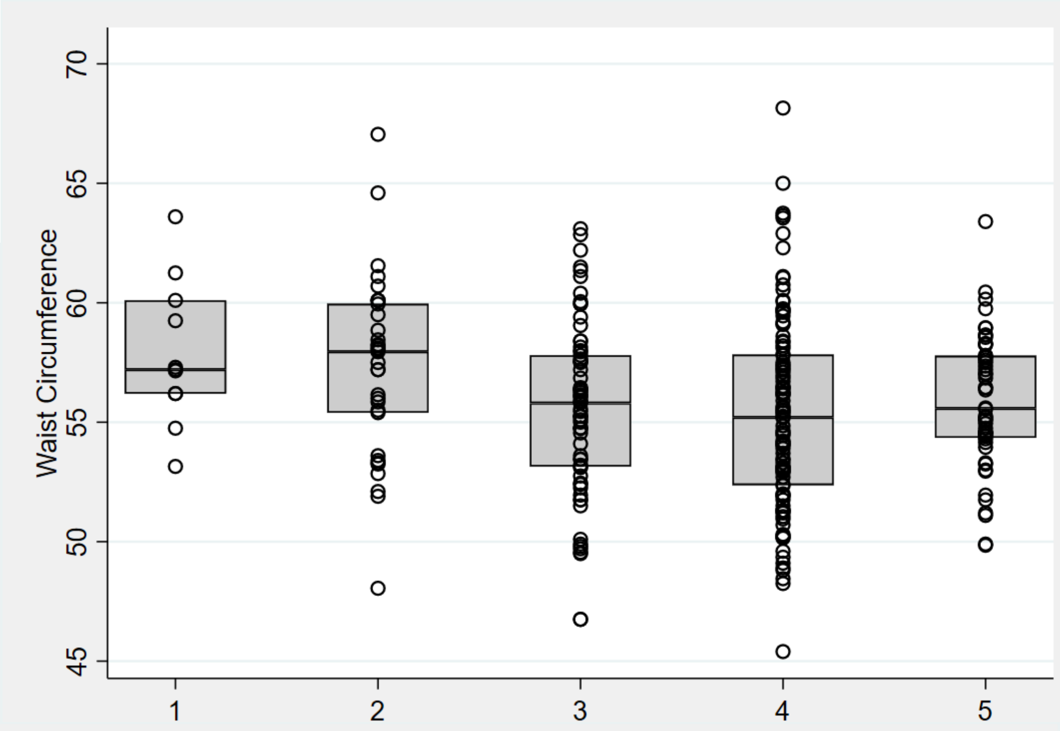
Waist Circumference
45
50
55
60
65
70
1
2
3
4
5

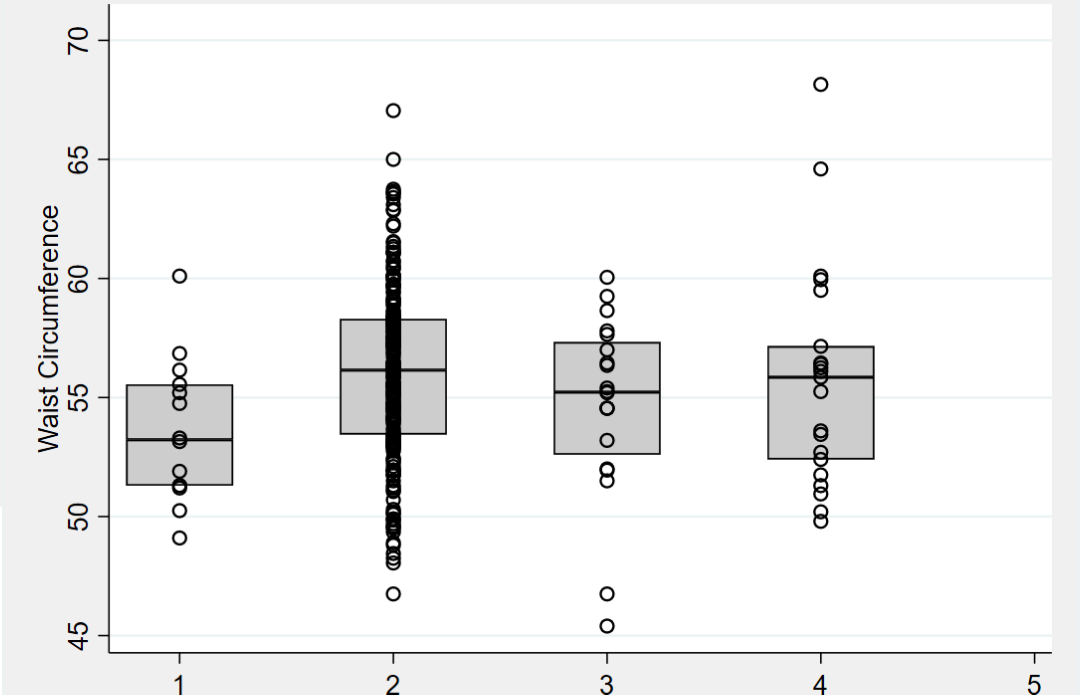
Waist Circumference
45
50
55
60
65
70
1
2
3
4
5

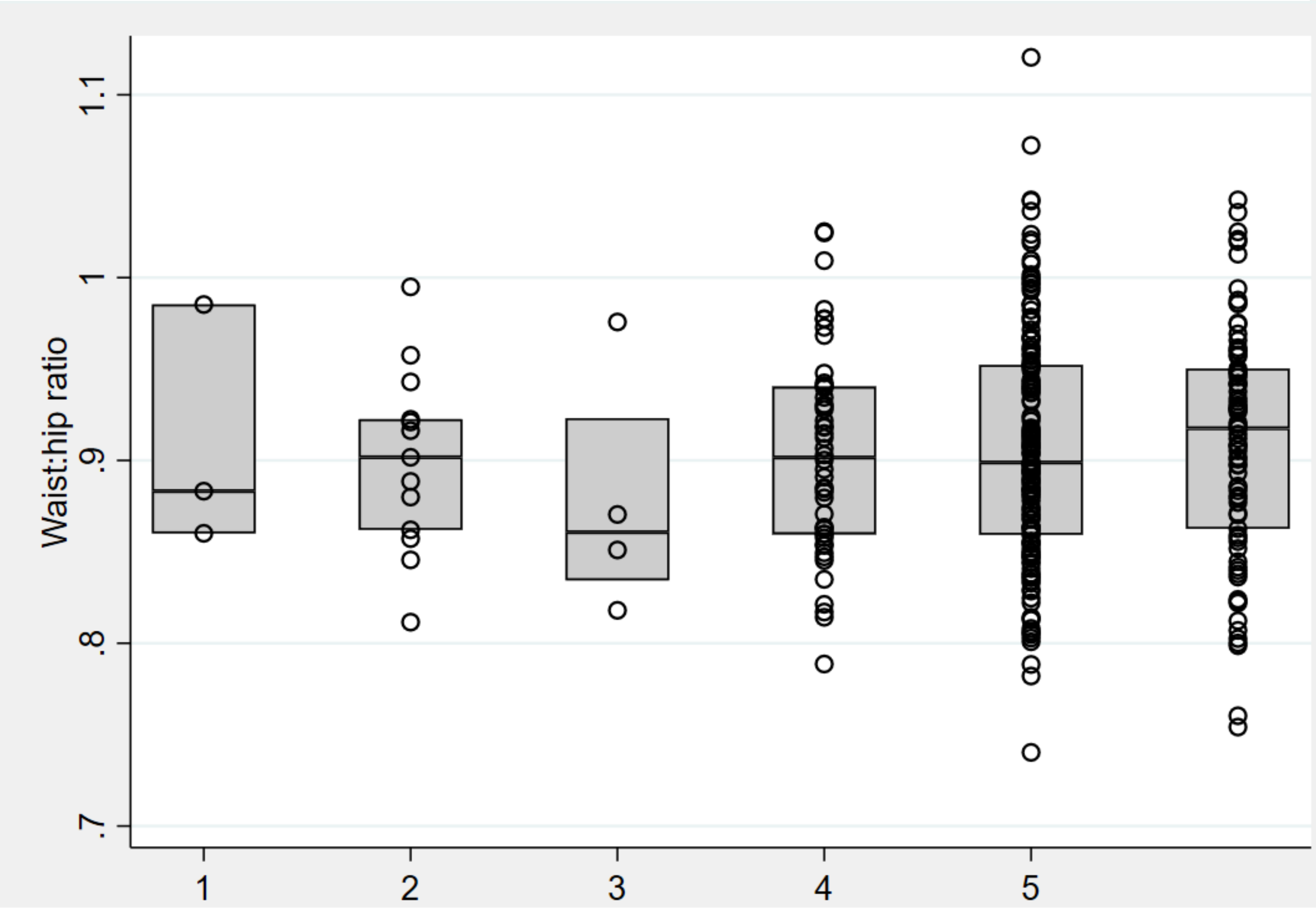
Waist:hip ratio
.7
.8
.9
1
1.1
1
2
3
4
5

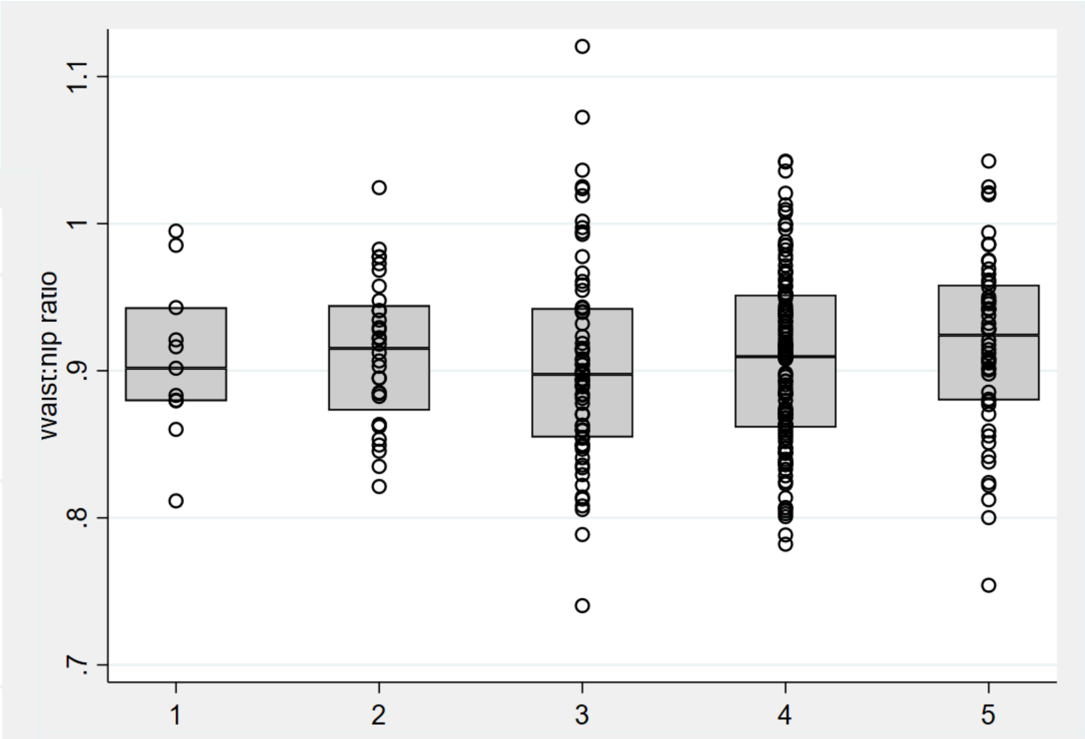
Waist:hip ratio
.7
.8
.9
1
1.1
1
2
3
4
5

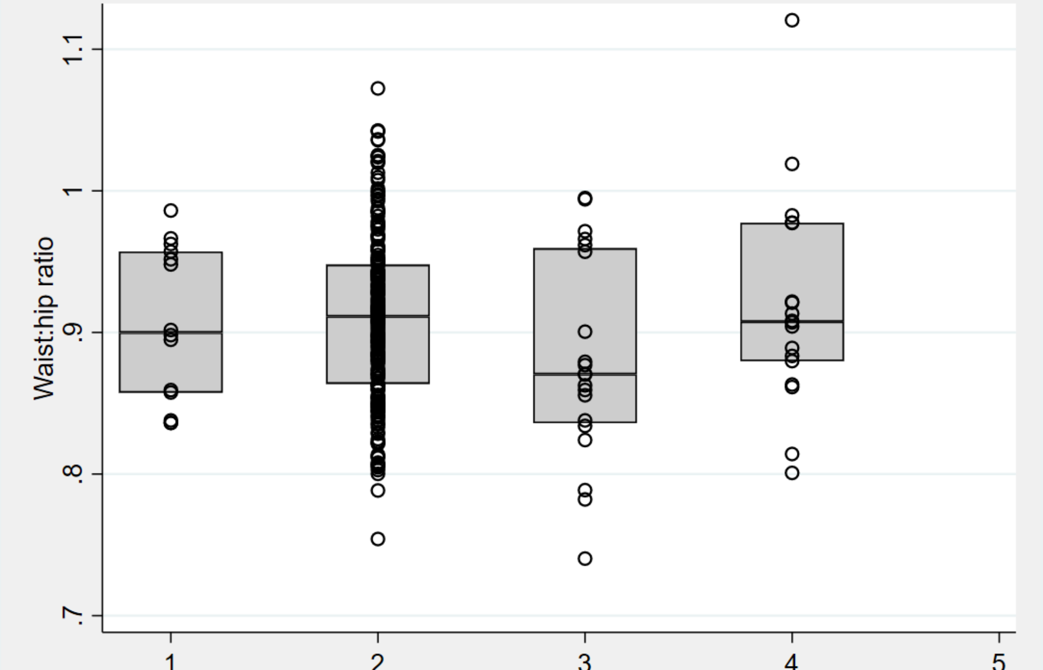
Waist:hip ratio
.7
.8
.9
1
1.1
1
2
3
4
5

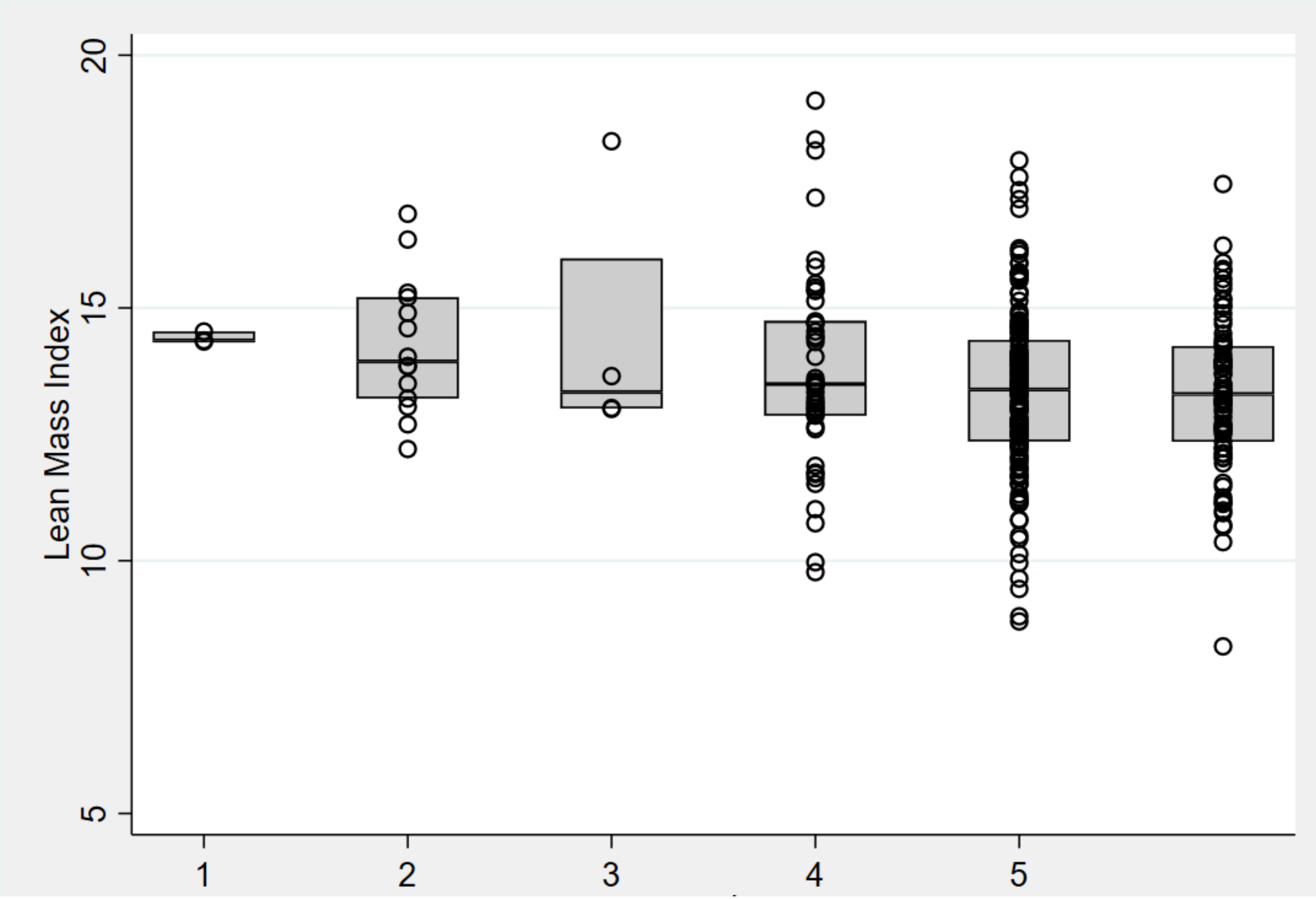
Lean Mass Index
5
10
15
20
1
2
3
4
5

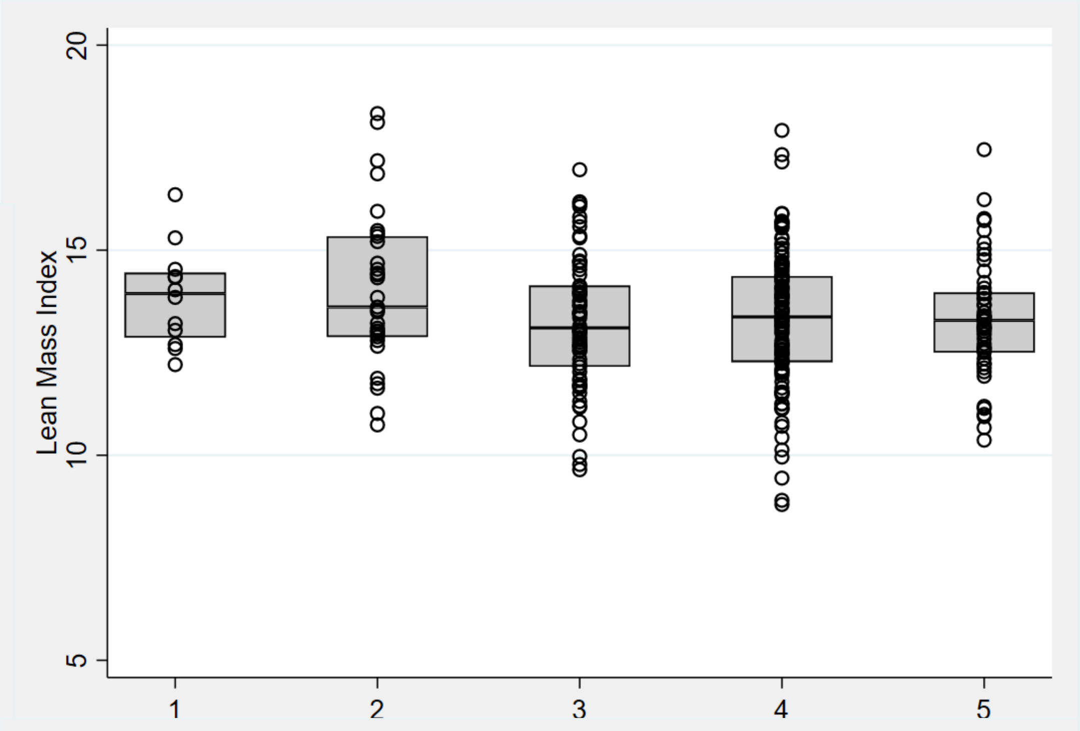
Lean Mass Index
5
10
15
20
1
2
3
4
5

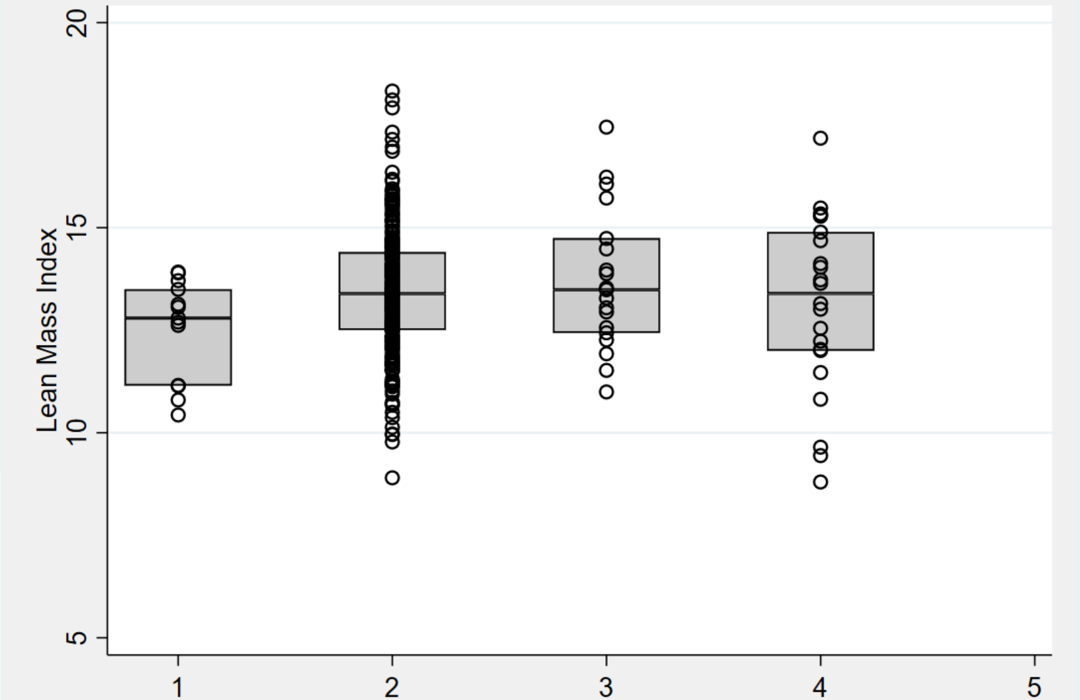
Lean Mass Index
5
10
15
20
1
2
3
4
5

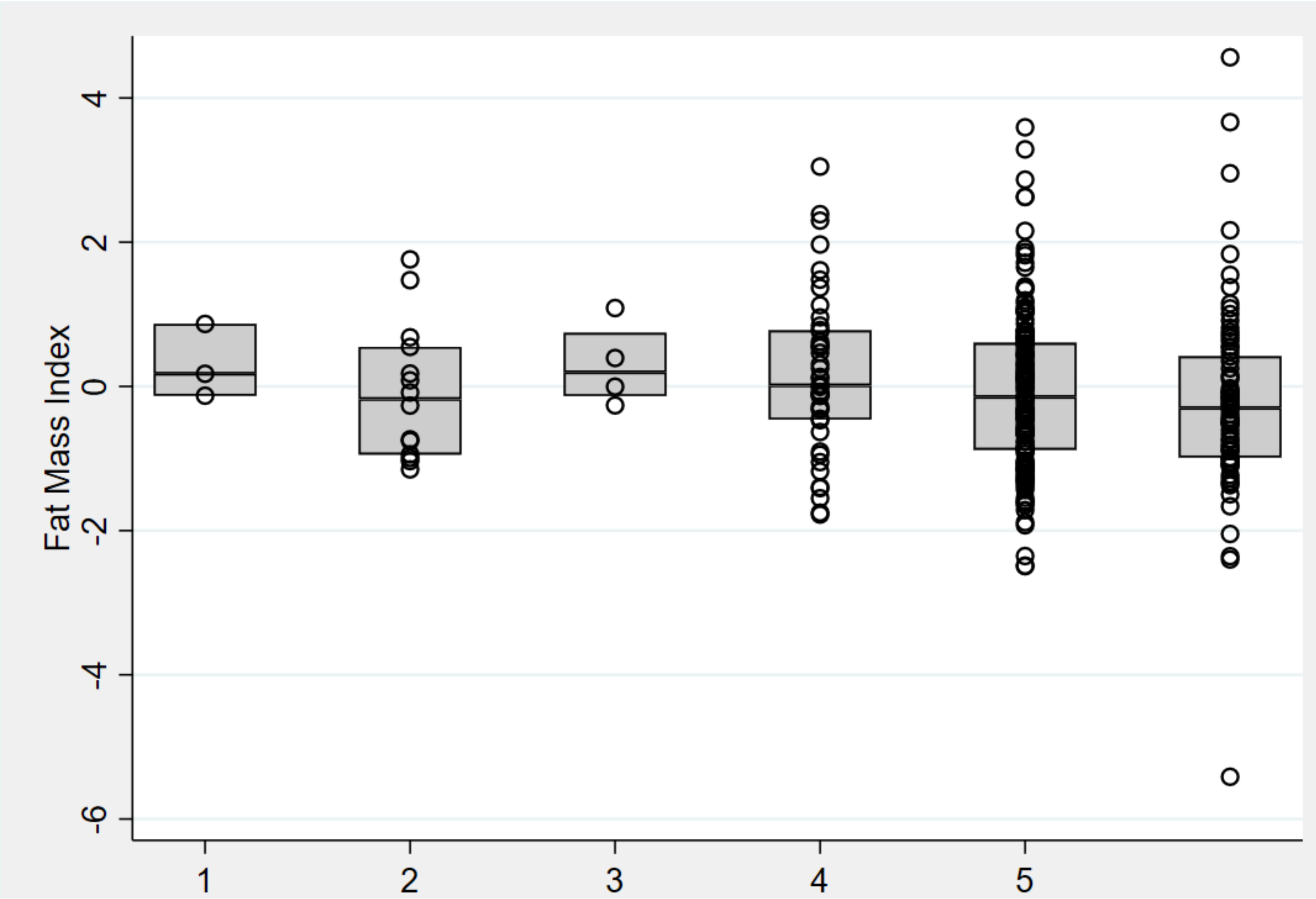
Fat Mass Index
4
2
0
-2
-4
-6
1
2
3
4
5

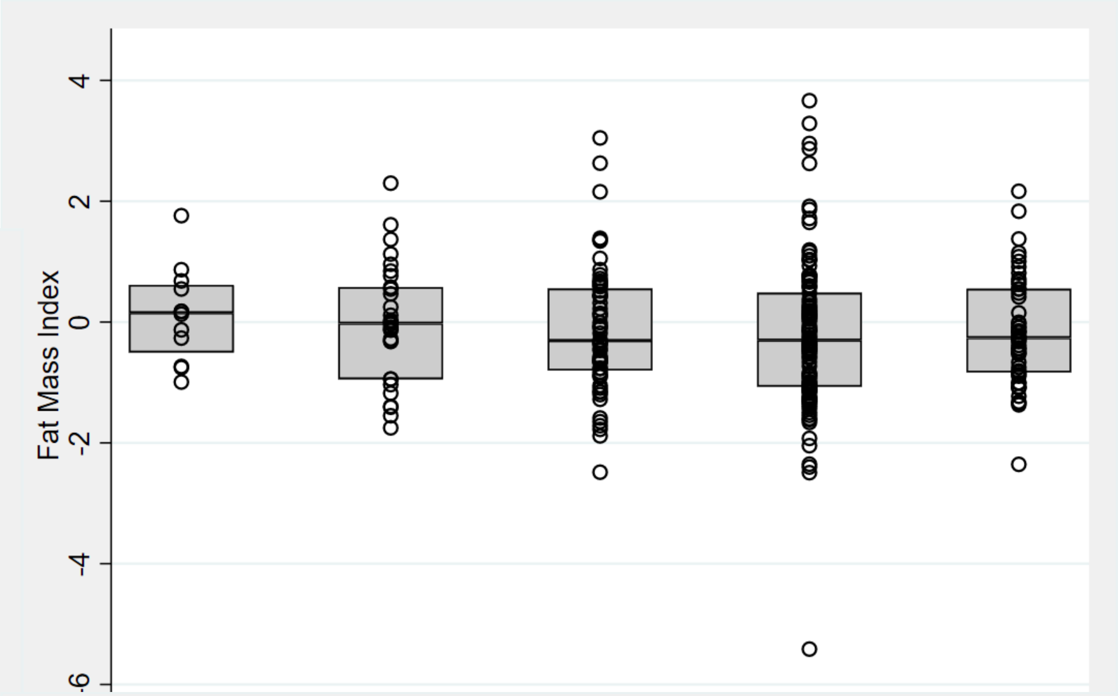
Fat Mass Index
4
2
0
-2
-4
-6

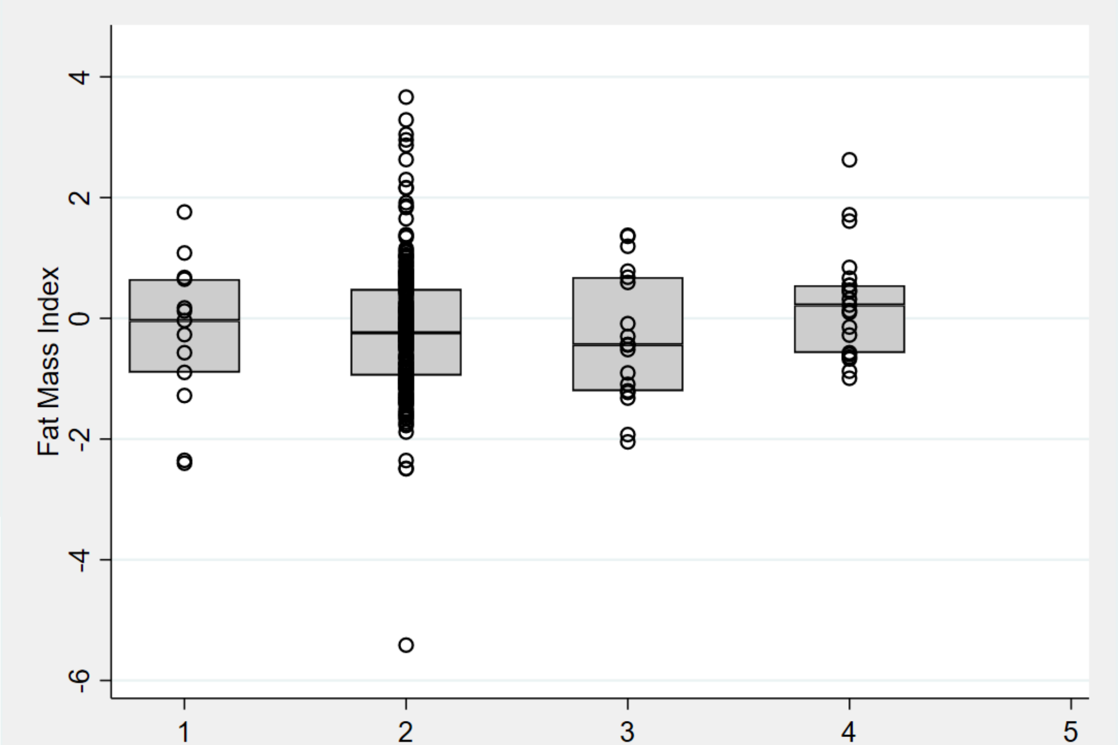
Fat Mass Index
4
2
0
-2
-4
-6
1
2
3
4
5
